# A prototype for decision support tool to help decision-makers with the strategy of handling the COVID-19 UK epidemic

**DOI:** 10.1101/2020.04.24.20077818

**Authors:** Anatoly Zhigljavsky, Ivan Fesenko, Henry Wynn, Roger Whitaker, Kobi Kremnizer, Jack Noonan, Jonathan Gillard

## Abstract

The primary objective of this work is to model and compare different exit scenarios from the lock-down for the COVID-19 UK epidemic. In doing so we provide an additional modelling basis for laying out the strategy options for the decision-makers. The main results are illustrated and discussed in Part I. In Part II, we describe the stochastic model that we have developed for modelling this epidemic. As argued in Part II, the developed model is more flexible than the SEIR/SEIRS models and can be used for modelling the scenarios which may be difficult or impossible to model with the SEIR/SEIRS models. To compare different scenarios for exiting from the lock-down, in Part III we provide our previous report on the same topic where similar (although not as detailed) scenarios were considered. As the possible exit dates, we have chosen May 4, May 11, May 18 and May 25.

We model differently the regions with high initial reproductive number chosen to be *R*_0_ = 2.5, medium *R*_0_ = 2.3 and low *R*_0_ = 2. The numbers for the whole of the UK can be obtained by appropriate averaging of the numbers given in the report. Typical figures are given in Section 4. For each scenario considered, we plot the expected proportion of infected at time *t* and the expected number of deaths at time *t*. To compute the expected numbers of deaths we used the total mortality rate 0.66%. Many recent studies suggest lower values and therefore the numbers in our projections should be considered as rather pessimistic. Our analysis suggests a value around 0.5% for the mortality rate.

In the model, we assume that the isolation of older and vulnerable people continues and the public carries on certain level of isolation until the end of 2020; also we assume that immunity is kept for at least a year and there is no international travel influence. Our main conclusions are:

- In regions with higher initial reproductive number 2.5 the proportion of susceptible at the start of the lock-down should be not smaller than 0.95, the epidemic curve in such regions is in the fast monotonic decline irrespectively of the date of the lock-down lift;
- In regions with lower initial reproductive number 2.0 the second mild wave can be expected, the difference between the expected mortality rates is very small for all May 2020 lifting lock-down dates;
- In regions with initial reproductive number 2.3, a mild second wave can be expected in the case of large proportion of susceptible at the start of the lock-down, but its severity and resulting mortality depend very little on the date of lifting the lock-down;
- For the overall UK epidemic, even for rather pessimistic scenarios considered, the second wave is much less pronounced (in terms of the expected mortality rate) than the first one, and the total numbers of expected deaths are within 2% for all May 2020 dates of lifting the lock-down. Moreover, by keeping *R*_0_-value after lifting the lock-down below 1.75 is likely to lead to the avoidance of a UK-wide second wave, see Section 4.

We believe that the model build in this work can be considered as an important decision support tool to help decision-makers with the strategy of handling the epidemic. We invite other scholars to participate in an open discussion of the strategy options. We feel that this kind of models should be used in the short and long term management of the disease. We recommend the development of a permanent and modularised modelling suite for COVID-19 management to which additional modules can be added as anti-viral drugs and vaccination are introduced, extending the options. We trust that this work makes a start in that direction and demonstrates the advantages of a heterogeneous demographic refinement, which can only improve targeting role out of treatments.

## Part I. Two-stage exit scenarios starting at different dates in May 2020

### Introduction

We show results describing further development of the COVID-19 epidemic in the UK given the current data and assuming different dates for lifting the lock-down, which we have chosen to be *t*_2_ = May 4, *t*_2_ = May 11, *t*_2_ = May 18, *t*_2_ = May 25. We use the model of the COVID-19 epidemic in the UK suggested and investigated in [1, 2]; see Part II for the description of the model.

We model differently the regions with high initial reproductive number chosen to be *R*_0_ = 2.5, medium *R*_0_ = 2.3 and low *R*_0_ = 2. The expected numbers of deaths are computed for a city or region with population 3 million people. The numbers for the whole of the UK can be obtained by appropriate averaging of the numbers given in the report. Typical figures are given in Section 4. We assume that each person in the UK is susceptible to COVID-19. If this is not the case, then the predictions will be more optimistic.

For each scenario considered, we plot the expected proportion of infected at time *t* and the expected number of deaths at time *t*. We also give values of the following related quantities: *x* – the proportion of susceptible on March 23, *I*_0_ – the proportion of infected on May 1, and *I*_1_ – the proportion of infected from the start of the epidemic up to May 1.

The cities with large *R*_0_ = 2.5 are considered in Section 1. The main findings of Section 1 are:

- *The scenarios of Section 1.1 with large R*_0_ *and large x* = 0.98 *(and hence small I*_0_ *and I*_1_*) seem to be inadequate for London* as in these scenarios the peak in expected deaths should be arriving at latter dates and this contradicts to the current situation.
- In case *R*_0_ = 2.5, the range of most suitable values for *x* is around 0.9 but not smaller than 0.95. *For such values of x, the epidemic in the badly hit regions of the UK is in the fast monotonic decline irrespectively of the date of the lock-down lift*.

The regions with *R*_0_ = 2 and *R*_0_ = 2.3 are considered in Sections 2 and 3. The main findings of these sections are:

- In most natural scenarios of Section 2 with *R*_0_ = 2, the second wave should be expected for all dates of lifting the lock-down, but it is going to be mild in all scenarios.
- *For R*_0_ = 2, *the difference between the expected deaths is very small for the lifting lock-down dates* May 4, May 11, May 18 *or* May 25, *for all cases considered in Section 2*.
- In the case *R*_0_ = 2.3, a mild second wave can be expected in the case of large *x* = 0.98 but its severity depends on the date of lifting the lock-down very little.

The overall UK epidemic is briefly considered in Section 4, see Figures 36 and 36.

- The presence of the second wave is explained by the fact that in the case considered, overall only 2.65% have been infected by March 23.
- Even in the worst-case scenario considered, the second wave is much less pronounced (in terms of the expected number of deaths) than the first one.
- The total numbers of expected deaths are within 2% for the four dates of lifting the lock-down.

### Notation, model parameters and description of plots

- *t* - time (in days)
- *t*_1_ - the lock-down time, *t*_1_ = March 23
- *t*_2_ - time when the lock-down finishes, *t*_2_ = May 4, *t*_2_ = May 11, *t*_2_ = May 18, *t*_2_ = May 25,
- *t*_3_ - time when more restrictions are lifted, *t*_3_ = *t*_2_+two weeks
- *I*(*t*) - number of infectious at time *t*
- *S*(*t*) - number of susceptible at time *t*;
- *R*_0_ - reproductive number before lock-down
- *R*_1_ - reproductive number during the lock-down (*t*_1_ ≤ *t < t*_2_)
- *R*_2_ - reproductive number for 2 weeks after the lock-down finishes (for *t*_2_ ≤ *t < t*_3_)
- *R*_3_ - reproductive number when more restrictions are lifted (for *t* ≥ *t*_3_)
- *x* - the proportion of susceptible at the start of the lock-down
- *I*_0_ - the proportion of infected on May 1
- *I*_1_ - the proportion of infected from the start of the epidemic up to May 1

In the model, *x* is a parameter but *I*_0_ and *I*_1_ are computed; their values are uniquely determined from *x* and other parameters of the model.

We consider a city of size 3 million people and use the recently updated number for the UK expected mortality rate *r* = 0.66%. Recent studies show that this rate is lower still but we shall keep this number. We keep the main parameters of the model as set in Section 7.2 in Part II, except we set *k*_*M*_ = *k*_*S*_ = 1 in order to be more consistent with SIR-based models (so that, some of our results could be re-checked using SIR-based models). As in Parts II and III, we assume that after lifting the lock-down, the group of vulnerable people and people of 70+ is going to be rather well isolated with *c* = 0.25. For the value of *R*_1_, we assume the following model. Initially, we suppose that every infected person will still infect all others in the same household, so that *R*_1_ could be larger than 1 for *t* close to March 23 and we assumed *R*_1_ = 1.25 during the week March 23 - March 29. Once the household has been infected, *R*_1_ drops to 0.95, a value slightly lower than 1 and what is recommended by a study [3] made for Germany (that is, we set *R*_1_ = 0.95 for *t* ≥ March 30) as there will be fewer new infections during the lock-down period.

In all plots colours relate to the date of lifting the lock-down:

- Purple: *t*_2_ = May 4,
- Green: *t*_2_ = May 11,
- Red: *t*_2_ = May 18,
- Black: *t*_2_ = May 25

In all figures with odd numbers, we use dashed lines and plot *I*(*t*)*/N*, the proportion of infected at time *t* (here *N* is the size of population considered). In all figures with even numbers, we use solid lines and plot expected deaths numbers at time *t*.

### 1 Regions with high initial reproductive number *R*_0_ = 2.5

#### 1.1 *x* = 0.98

Here we have *I*_0_ = 0.011, *I*_1_ = 0.143.

##### 1.1.1 *R*_2_ = 1.75, *R*_3_ = 2

**Figure 1:**
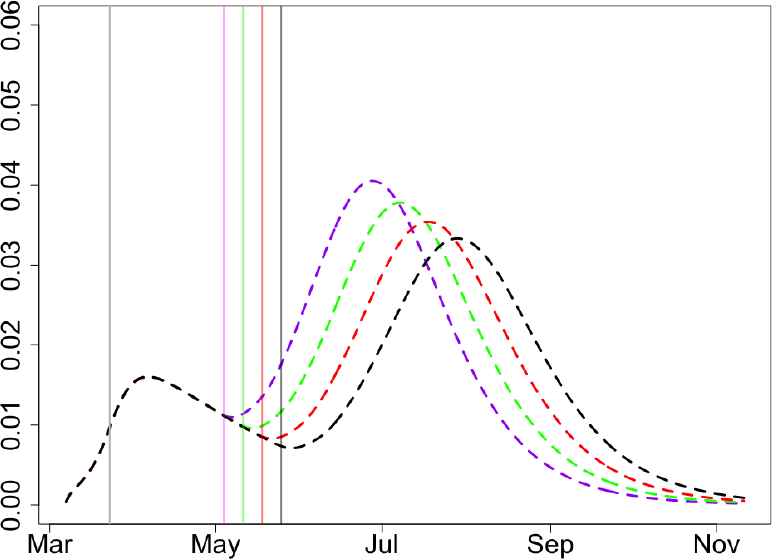
Proportion of infectious, *x* = 0.98, *R*_2_ = 1.75, *R*_3_ = 2.

**Figure 2:**
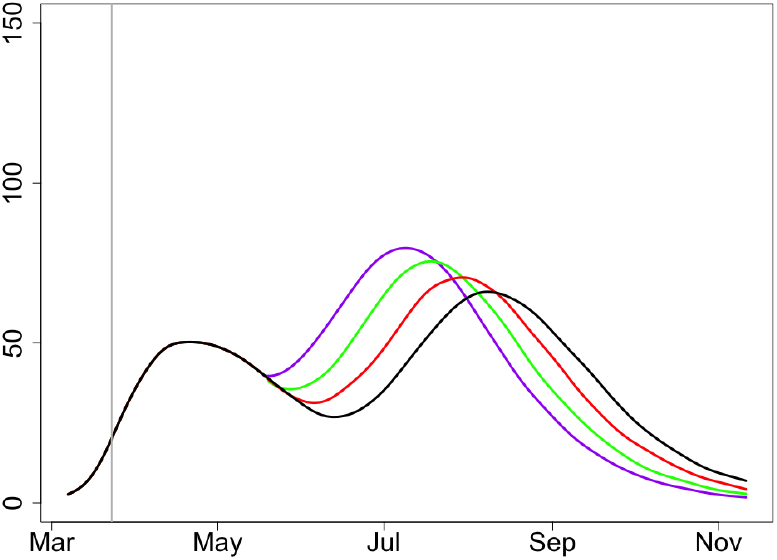
Expected deaths, *x* = 0.98, *R*_2_ = 1.75, *R*_3_ = 2.

##### 1.1.2 *R*_2_ = 1.5, *R*_3_ = 1.75

**Figure 3:**
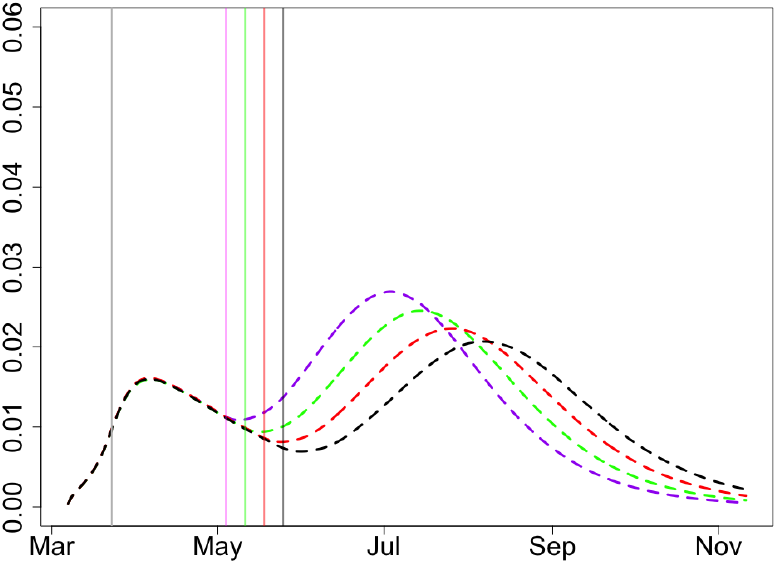
Proportion of infectious, *x* = 0.98, *R*_2_ = 1.5, *R*_3_ = 1.75.

**Figure 4:**
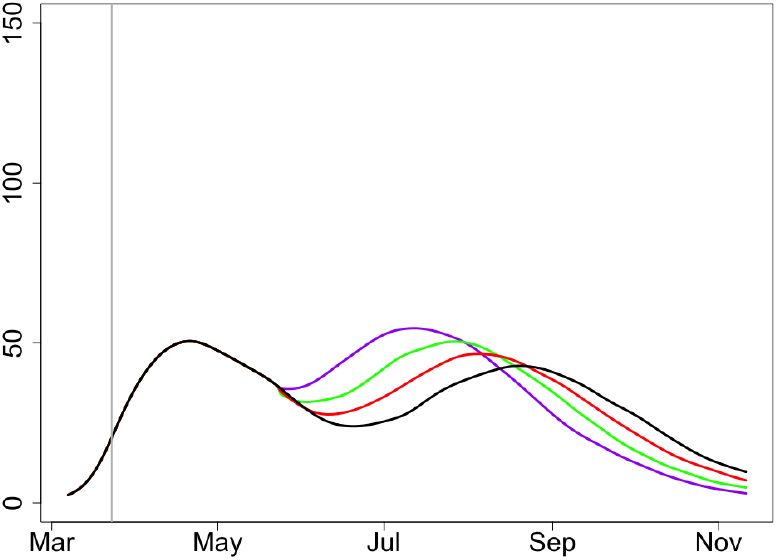
Expected deaths, *x* = 0.98, *R*_2_ = 1.5, *R*_3_ = 1.75.

This scenario, with large *R*_0_ and large *x*, seems to be inadequate for a city like London; in case of large *x* and hence small *I*_0_ and *I*_1_, the peak in expected deaths is shifted too far to the later dates. Indeed, in London the mortality seems to be steadily going down from about April 20.

#### 1.2 *x* = 0.95

Here we have *I*_0_ = 0.0179, *I*_1_ = 0.277.

##### 1.2.1 *R*_2_ = 1.75, *R*_3_ = 2

**Figure 5:**
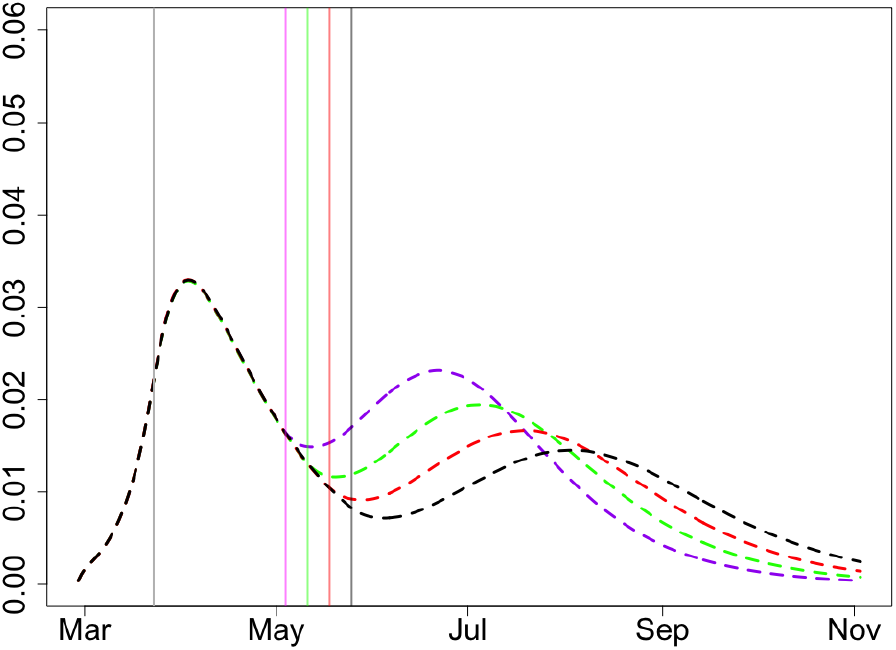
Proportion of infectious, *x* = 0.95, *R*_2_ = 1.75, *R*_3_ = 2.

**Figure 6:**
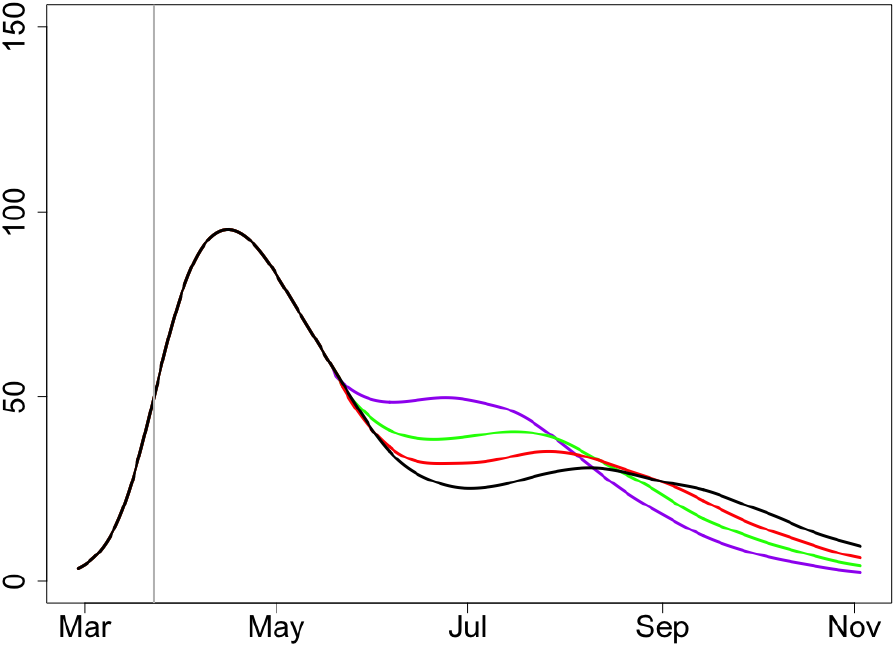
Expected deaths, *x* = 0.95, *R*_2_ = 1.75, *R*_3_ = 2.

##### 1.2.2 *R*_2_ = 1.5, *R*_3_ = 1.75

**Figure 7:**
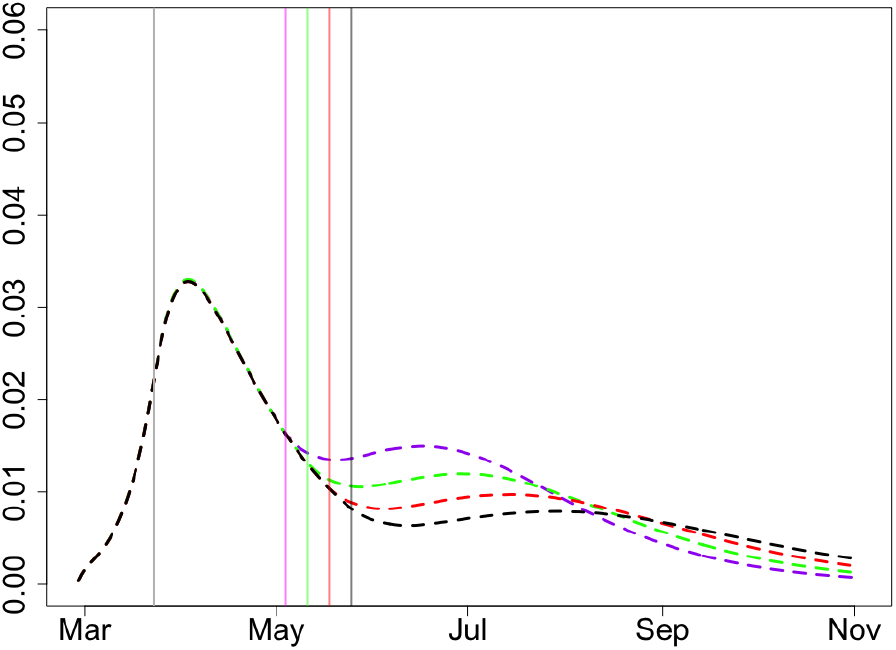
Proportion of infectious, *x* = 0.95, *R*_2_ = 1.5, *R*_3_ = 1.75.

**Figure 8:**
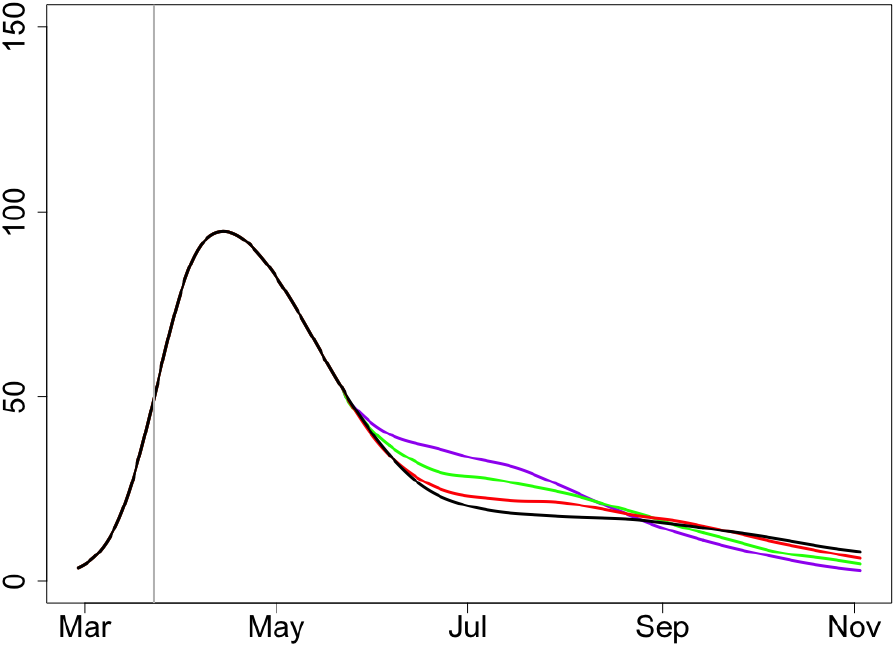
Expected deaths, *x* = 0.95, *R*_2_ = 1.5, *R*_3_ = 1.75.

In view of the location of the peaks in the curve of the expected deaths, in the case *R*_0_ = 2.5, the scenarios with *x* = 0.95 seem to be much more likely than with *x* = 0.98 but less likely than with *x* = 0.9, for London and other areas with high *R*_0_. For *x* = 0.95, a visible second wave in the number of infections and deaths is not expected irrespectively of the date of lifting the lock-down.

#### 1.3 *x* = 0.93

Here we have *I*_0_ = 0.019, *I*_1_ = 0.342.

##### 1.3.1 *R*_2_ = 1.75, *R*_3_ = 2

**Figure 9:**
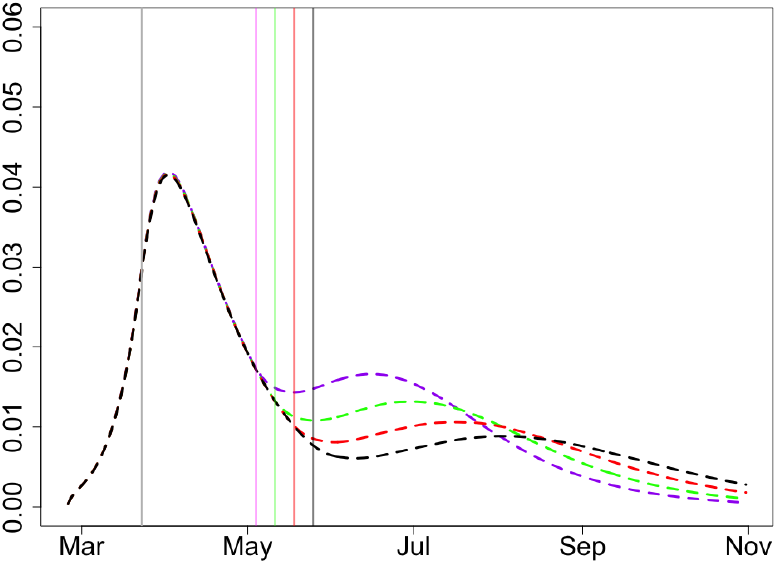
Proportion of infectious, *x* = 0.93, *R*_2_ = 1.75, *R*_3_ = 2.

**Figure 10:**
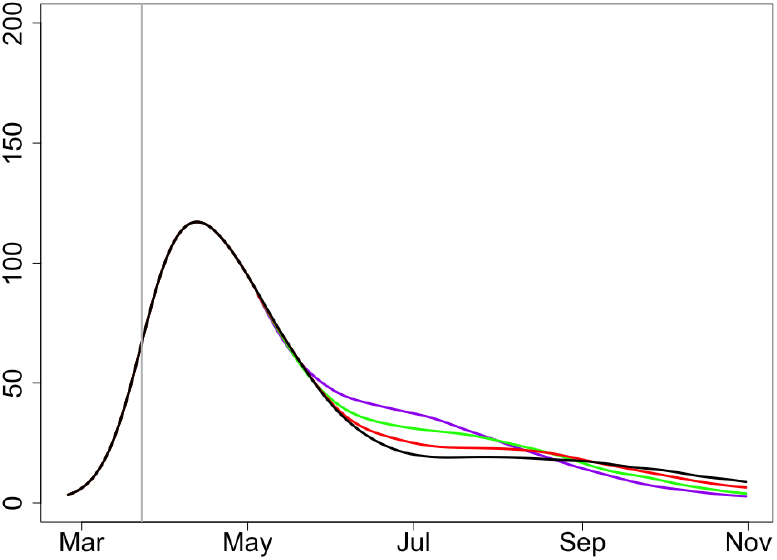
Expected deaths, *x* = 0.93, *R*_2_ = 1.75, *R*_3_ = 2.

##### 1.3.2 *R*_2_ = 1.5, *R*_3_ = 1.75

**Figure 11:**
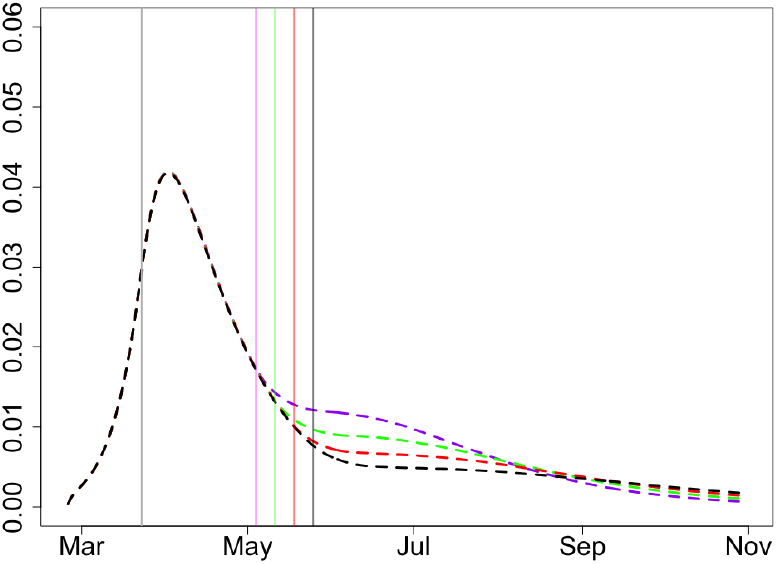
Proportion of infectious, *x* = 0.93, *R*_2_ = 1.5, *R*_3_ = 1.75.

**Figure 12:**
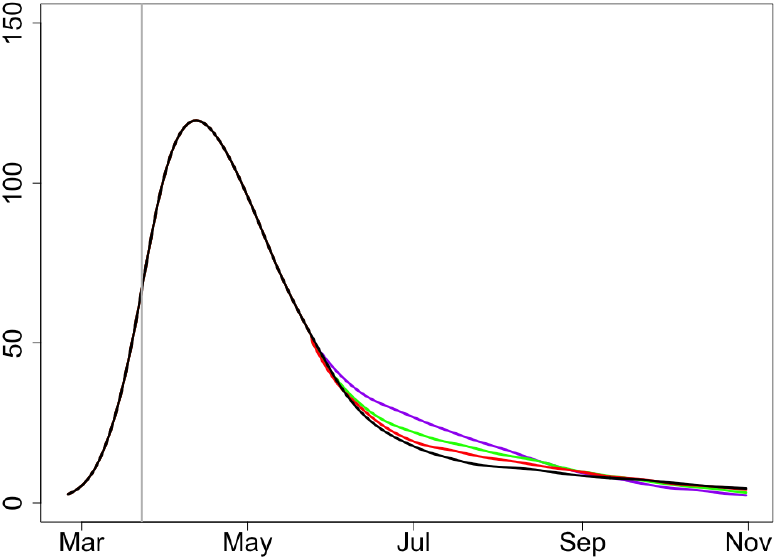
Expected deaths, *x* = 0.93, *R*_2_ = 1.5, *R*_3_ = 1.75.

In view of the location of the peaks in the curve of the expected deaths, in the case *R*_0_ = 2.5, the scenarios with *x* = 0.93 seem to be very likely for London and other areas with high *R*_0_. For *x* = 0.93, the second wave in the number of infections and deaths is not expected irrespectively of the date of lifting the lock-down.

#### 1.4 *x* = 0.9

Here we have *I*_0_ = 0.0194, *I*_1_ = 0.429.

##### 1.4.1 *R*_2_ = 1.75, *R*_3_ = 2

**Figure 13:**
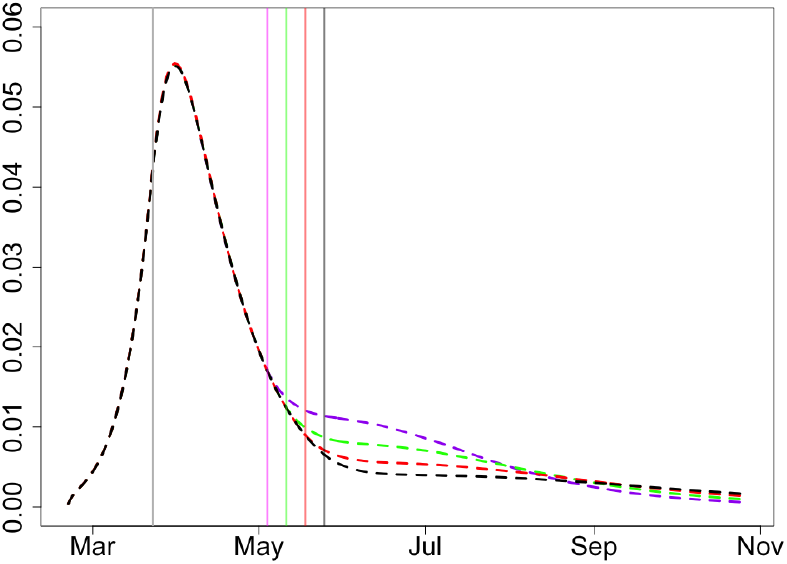
Proportion of infectious, *x* = 0.9, *R*_2_ = 1.75, *R*_3_ = 2.

**Figure 14:**
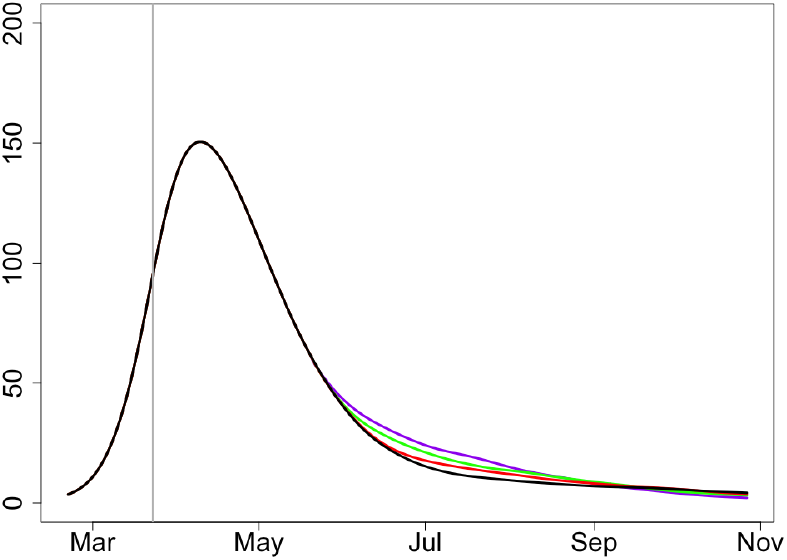
Expected deaths, *x* = 0.9, *R*_2_ = 1.75, *R*_3_ = 2.

##### 1.4.2 *R*_2_ = 1.5, *R*_3_ = 1.75

**Figure 15:**
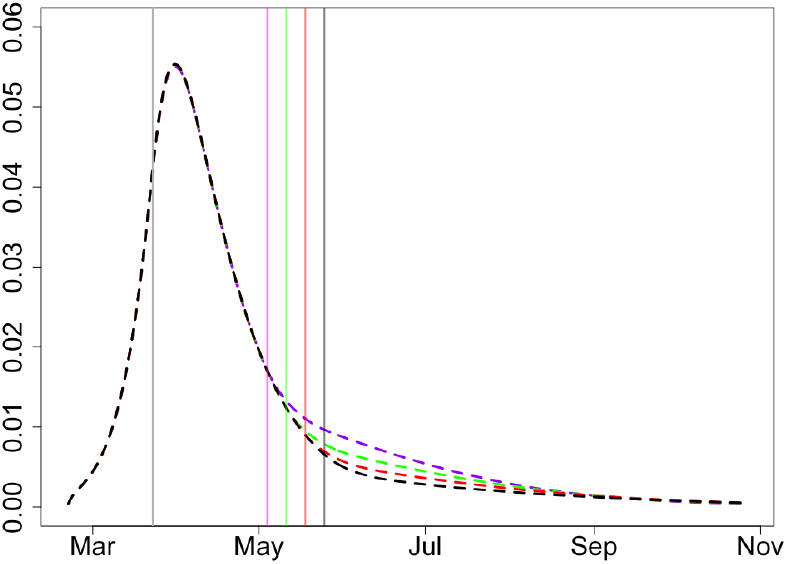
Proportion of infectious, *x* = 0.9, *R*_2_ = 1.5, *R*_3_ = 1.75.

**Figure 16:**
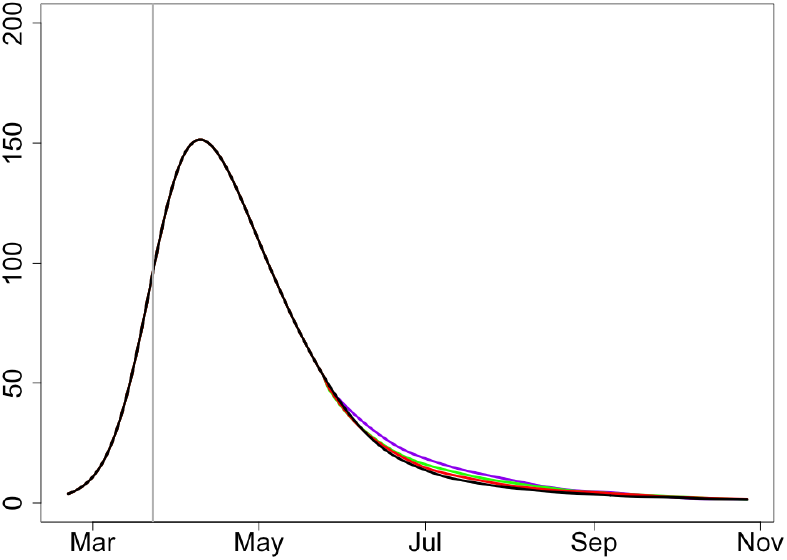
Expected deaths, *x* = 0.9, *R*_2_ = 1.5, *R*_3_ = 1.75.

The value *x* ≃ 0.9 matches the current deaths data in London quite well (assuming large reproductive number *R*_0_). *If indeed x* ≃ 0.9 *or larger, then the epidemic in London is finishing very soon*. It is then guaranteed that there will be no second wave. The curves describing expected infections and deaths do not depend on the date of lifting the lock-down.

### 2 Regions with low initial reproductive number *R*_0_ = 2

#### 2.1 *x* = 0.98

Here we have *I*_0_ = 0.009, *I*_1_ = 0.115.

##### 2.1.1 *R*_2_ = 1.5, *R*_3_ = 1.75

**Figure 17:**
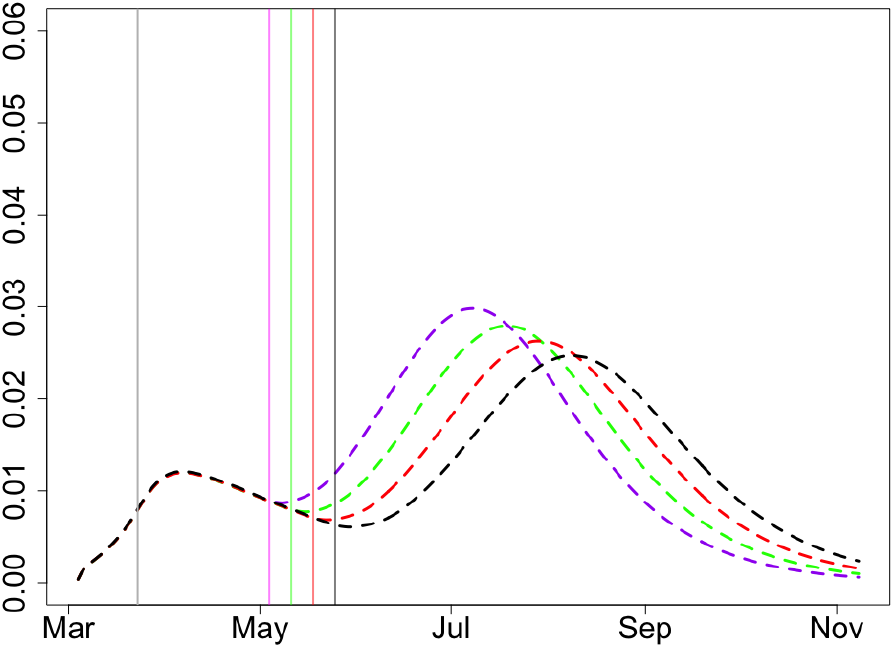
Proportion of infectious, *x* = 0.98, *R*_2_ = 1.5, *R*_3_ = 1.75.

**Figure 18:**
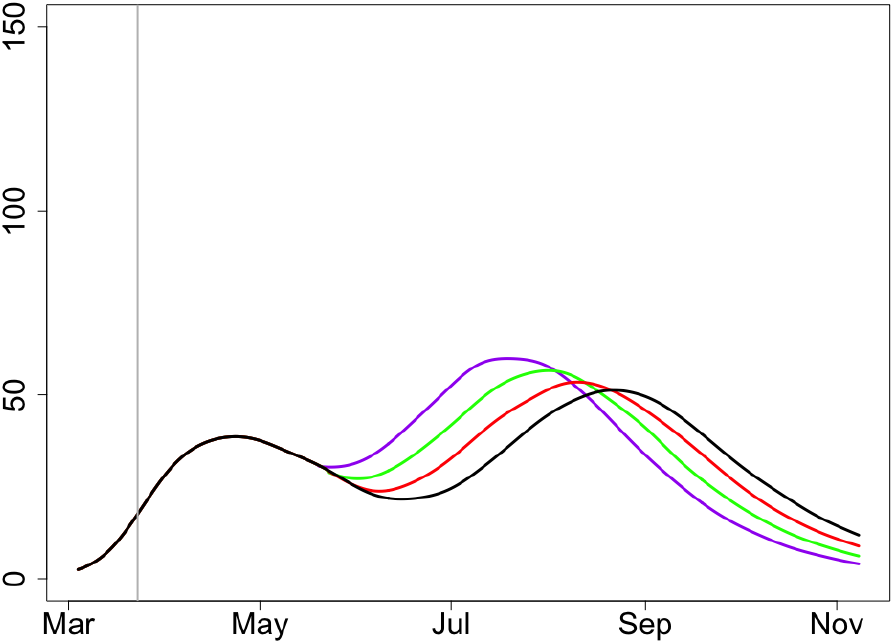
Expected deaths, *x* = 0.98, *R*_2_ = 1.5, *R*_3_ = 1.75.

##### 2.1.2 *R*_2_ = 1.3, *R*_3_ = 1.5

**Figure 19:**
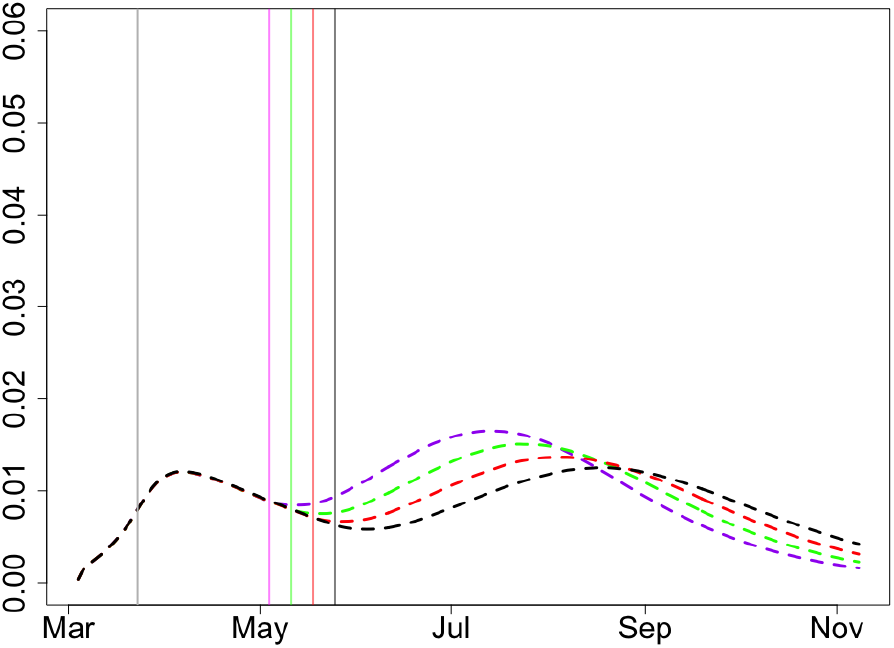
Proportion of infectious, *x* = 0.98, *R*_2_ = 1.3, *R*_3_ = 1.5.

**Figure 20:**
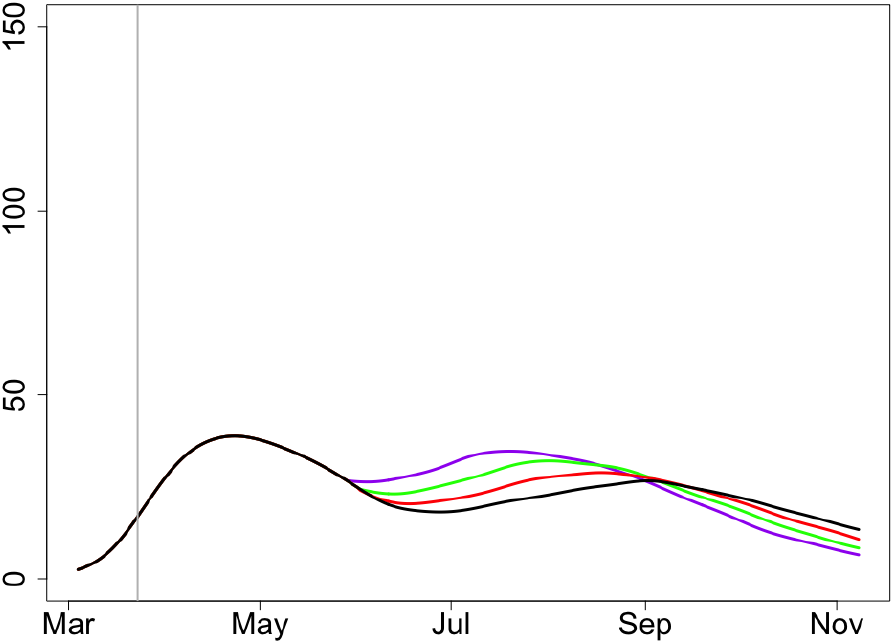
Expected deaths, *x* = 0.98, *R*_2_ = 1.3, *R*_3_ = 1.5.

The scenario with *R*_0_ = 2 and very large *x* seems very reasonable. The second wave should be expected in all cases but please note the scales for the proportions of infected and expected deaths; the numbers are much smaller than in the case *R*_0_ = 2.5. The second wave is expected but (a) it is going to be mild in all cases, and (b) the dependence of the severity of the second wave is not much different for all dates of lifting the lock-down.

#### 2.2 *x* = 0.95

Here we have *I*_0_ = 0.0143, *I*_1_ = 0.216.

##### 2.2.1 *R*_2_ = 1.5, *R*_3_ = 1.75

**Figure 21:**
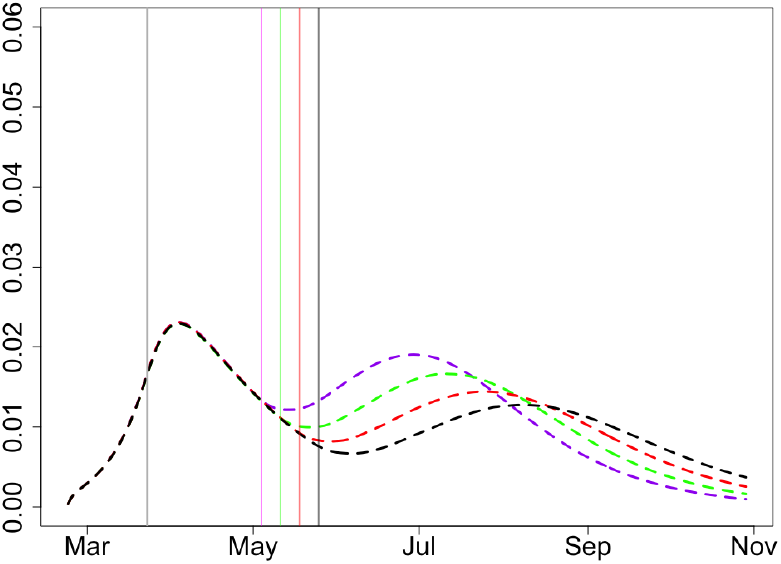
Proportion of infectious, *x* = 0.95, *R*_2_ = 1.5, *R*_3_ = 1.75.

**Figure 22:**
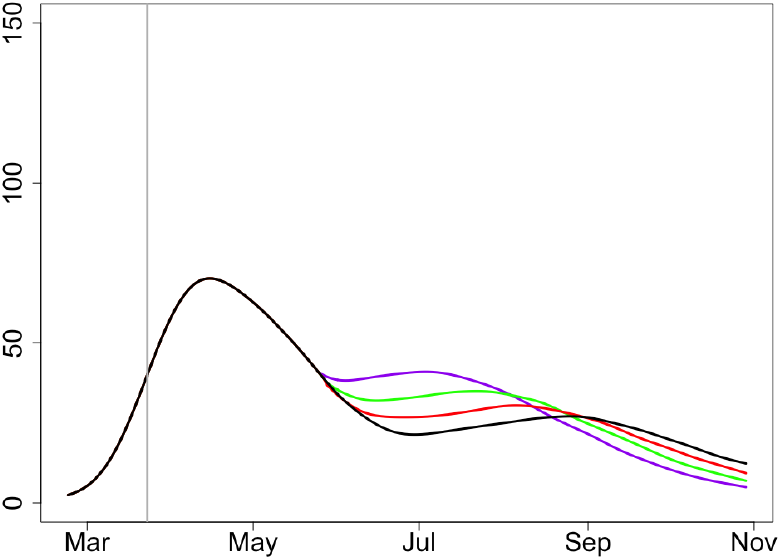
Expected deaths, *x* = 0.95, *R*_2_ = 1.5, *R*_3_ = 1.75.

##### 2.2.2 *R*_2_ = 1.3, *R*_3_ = 1.5

**Figure 23:**
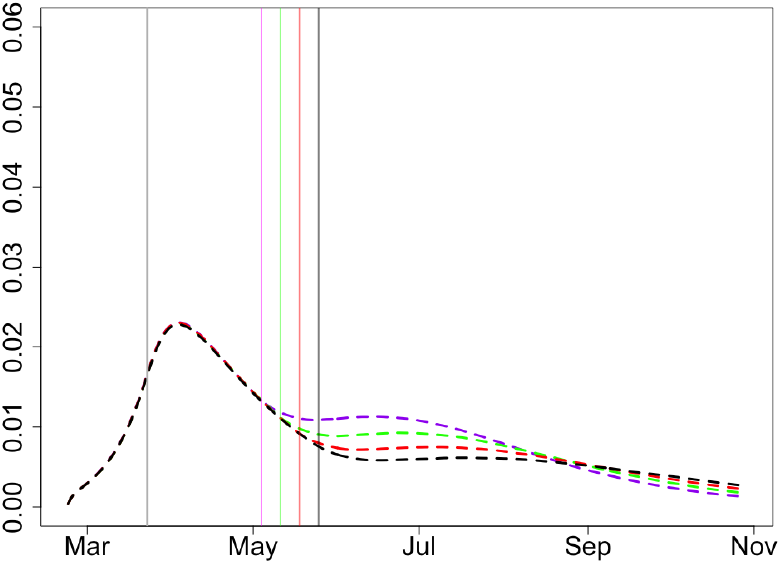
Proportion of infectious, *x* = 0.95, *R*_2_ = 1.3, *R*_3_ = 1.5.

**Figure 24:**
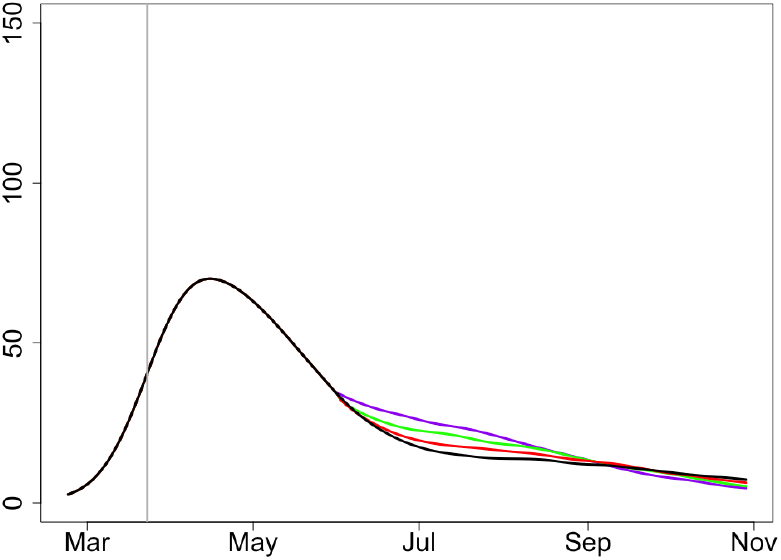
Expected deaths, *x* = 0.95, *R*_2_ = 1.3, *R*_3_ = 1.5.

The scenario with *R*_0_ = 2 and very large *x* = 0.95 seems quite reasonable. No second wave is expected for all dates of lifting the lock-down.

#### 2.3 *x* = 0.9

Here we have *I*_0_ = 0.170, *I*_1_ = 0.3357.

##### 2.3.1 *R*_2_ = 1.5, *R*_3_ = 1.75

**Figure 25:**
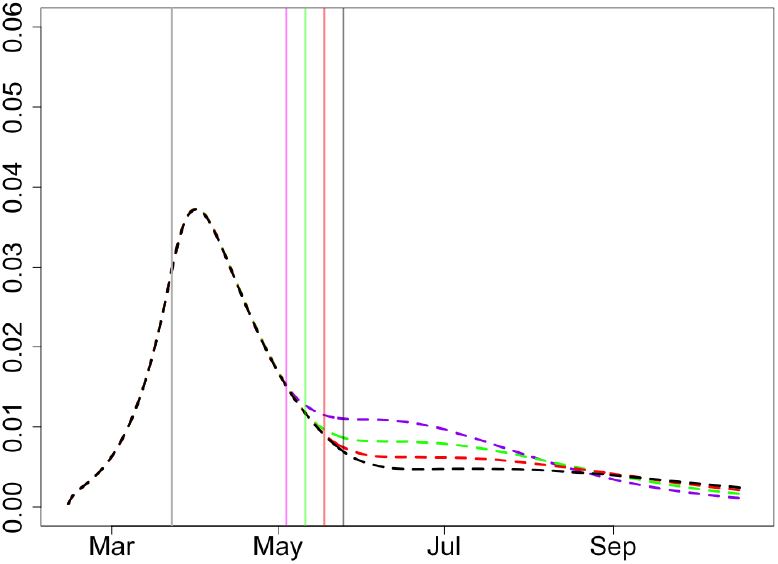
Proportion of infectious, *x* = 0.9, *R*_2_ = 1.5, *R*_3_ = 1.75.

**Figure 26:**
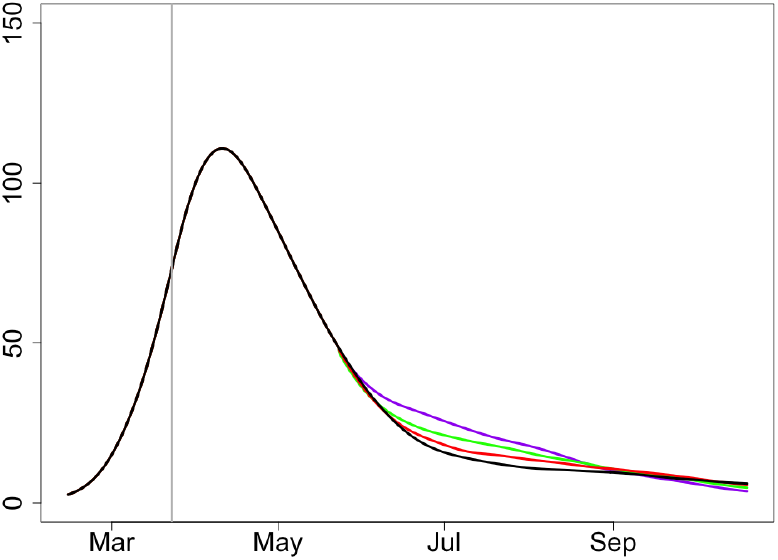
Expected deaths, *x* = 0.9, *R*_2_ = 1.5, *R*_3_ = 1.75.

##### 2.3.2 *R*_2_ = 1.3, *R*_3_ = 1.5

**Figure 27:**
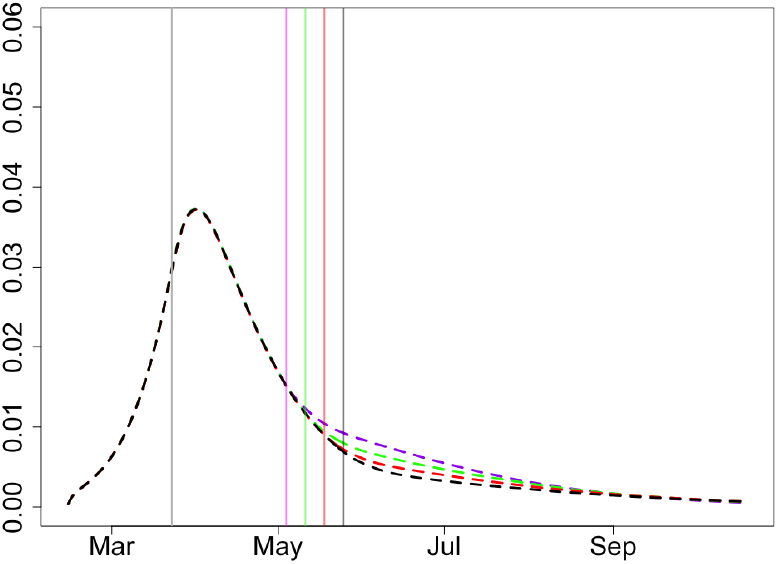
Proportion of infectious, *x* = 0.9, *R*_2_ = 1.3, *R*_3_ = 1.5.

**Figure 28:**
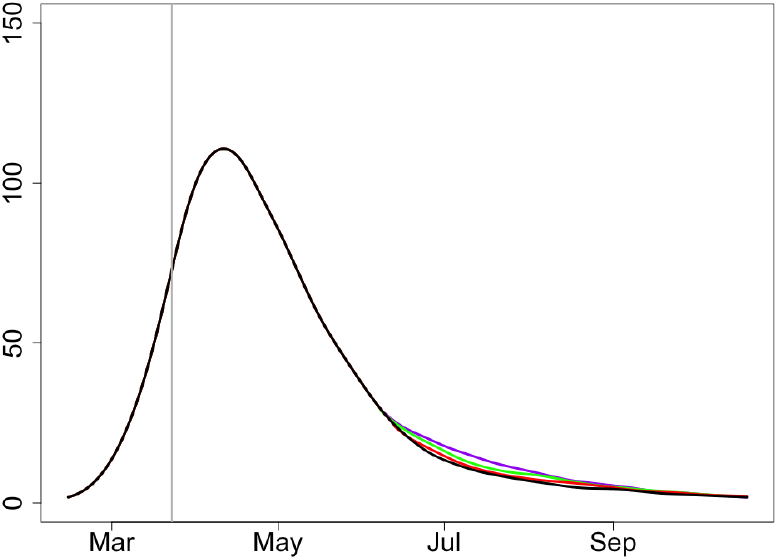
Expected deaths, *x* = 0.9, *R*_2_ = 1.3, *R*_3_ = 1.5.

The value *x ∼* 0.9 seems to be slightly too high assuming small reproductive number *R*_0_. No second wave is expected for any date of lifting the lock-down.

### 3 Regions with medium initial reproductive number *R*_0_ = 2.3

#### 3.1 *x* = 0.98

Here we have *I*_0_ = 0.0109, *I*_1_ = 0.133.

#### 3.2 *x* = 0.97

Here we have *I*_0_ = 0.0136, *I*_1_ = 0.1827.

**Figure 29:**
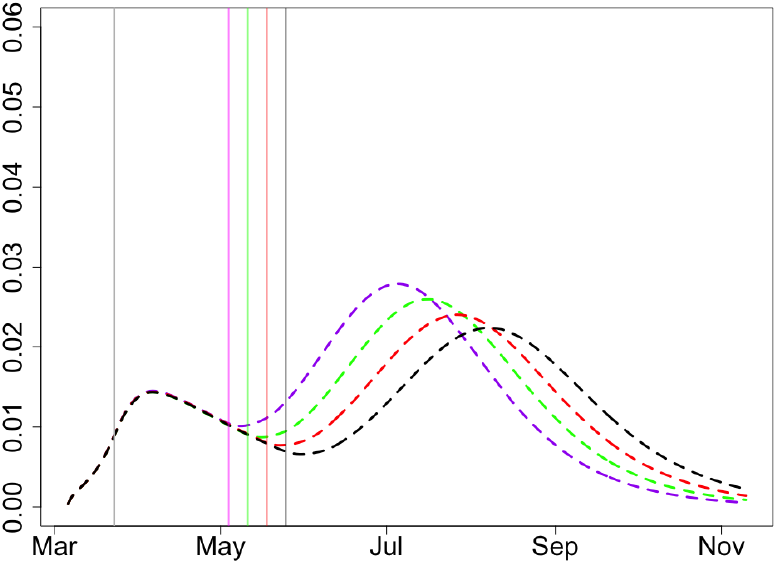
Proportion of infectious, *x* = 0.98, *R*_2_ = 1.5, *R*_3_ = 1.75.

**Figure 30:**
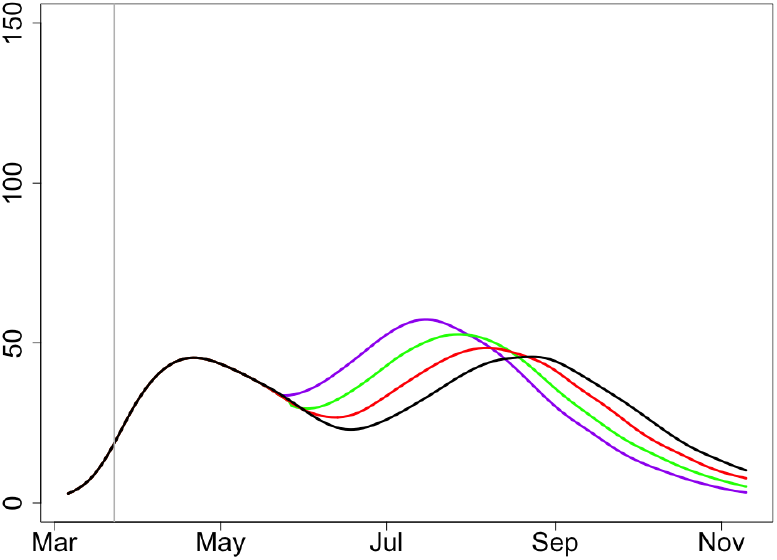
Expected deaths, *x* = 0.98, *R*_2_ = 1.5, *R*_3_ = 1.75.

**Figure 31:**
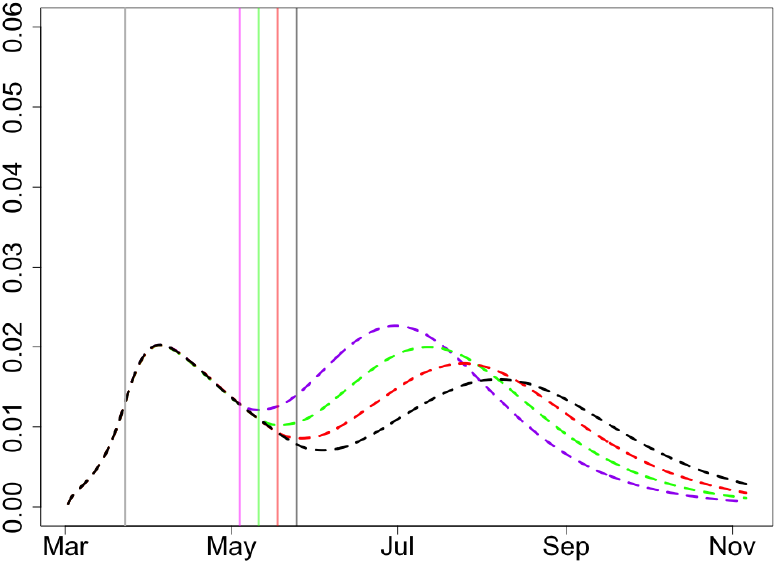
Proportion of infectious, *x* = 0.97, *R*_2_ = 1.5, *R*_3_ = 1.75.

**Figure 32:**
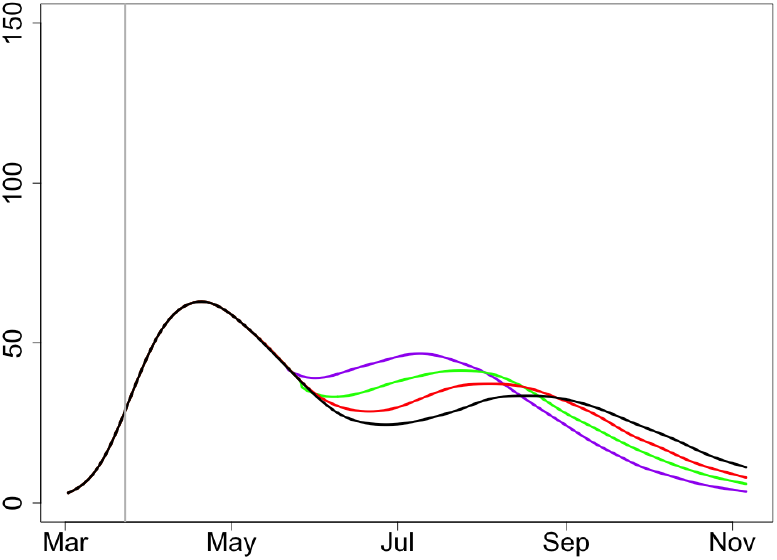
Expected deaths, *x* = 0.97, *R*_2_ = 1.5, *R*_3_ = 1.75.

**Figure 33:**
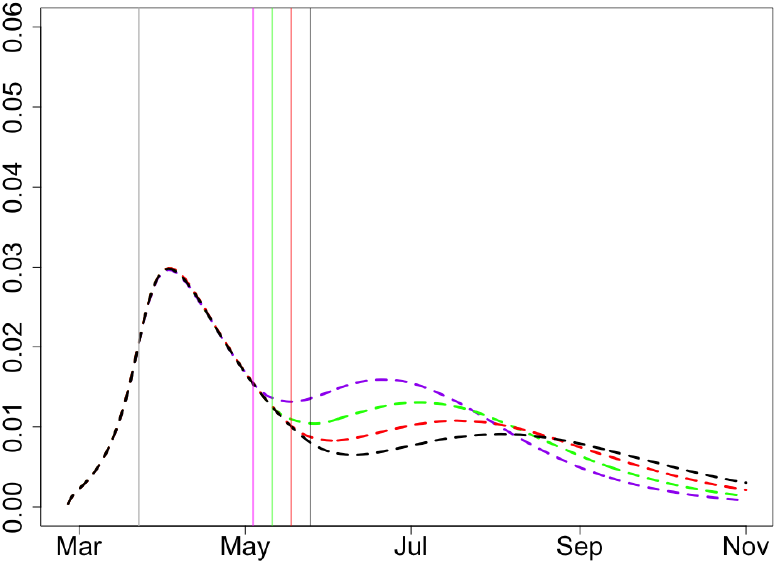
Proportion of infectious, *x* = 0.95, *R*_2_ = 1.5, *R*_3_ = 1.75.

**Figure 34:**
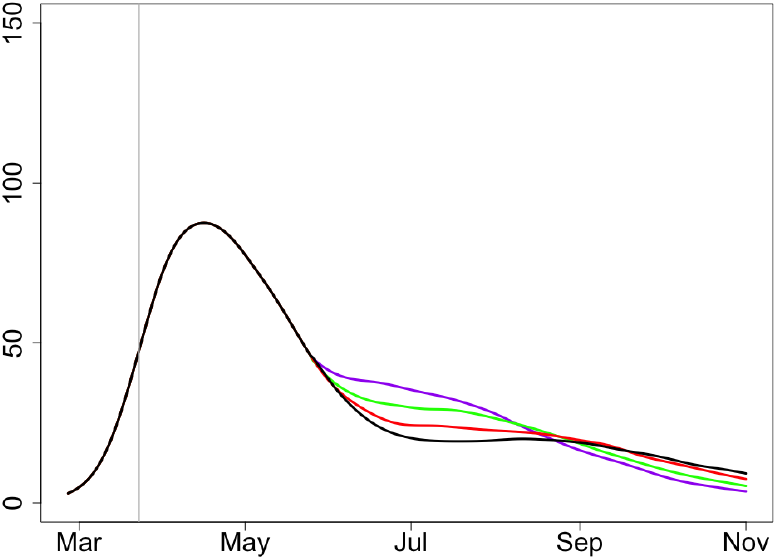
Expected deaths, *x* = 0.95, *R*_2_ = 1.5, *R*_3_ = 1.75.

#### 3.3 *x* = 0.95

Here we have *I*_0_ = 0.0168, *I*_1_ = 0.259.

All three scenarios, with *x* = 0.95, 0.97, 0.98 look very possible in the case *R*_0_ = 2.3. A very mild second wave can be expected in the case of large *x* = 0.98 but its severity does very little depends on the date of lifting the lock-down.

### 4 Typical scenarios for the entire UK

In this section, we provide two typical scenarios for the entire country. They are based on appropriate averaging of the scenarios discussed above. We have averaged the following three very likely scenarios for regions with different *R*_0_:

a. *R*_0_ = 2.5, *x* = 0.9, *R*_2_ = 1.5, *R*_3_ = 1.75, see Section 1.2.2;
b. *R*_0_ = 2.3, *x* = 0.95, *R*_2_ = 1.5, *R*_3_ = 1.75, see Section 3.3;
c. *R*_0_ = 2.0, *x* = 0.99, *R*_2_ = 1.3, *R*_3_ = 1.5, similar to Section 3.1 where *x* = 0.98.

In the case *R*_0_ = 2.0, we have chosen the worst possible case in terms of *x*; that is, we have assumed that only 1% of people in the corresponding regions have been infected before March 23. This seems to be too low but we wanted to show the worst-case scenario.

We have considered two different mixing schemes for the regions satisfying (a)-(c). Assuming that the total population of the UK is 66 million, we averaged the populations in the following proportions.

**Scheme 1**. (a) - 8m, (b) - 8m, (c) 50m; see Figures 35 and 36;

**Figure 35:**
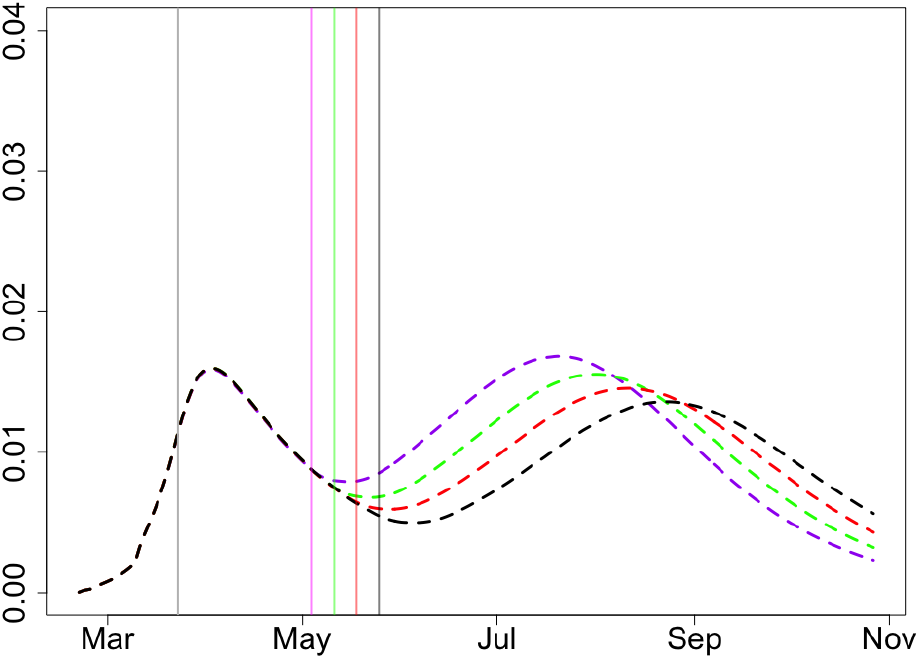
Proportion of infectious, UK, Scheme 1.

**Figure 36:**
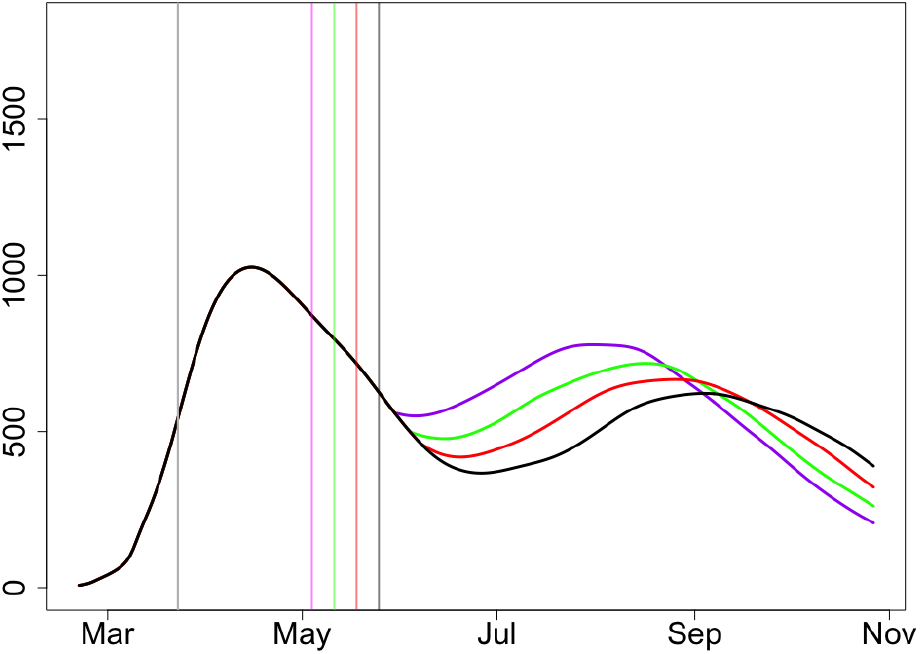
Expected deaths, UK, Scheme 1.

**Scheme 2**. (a) - 6m, (b) - 30m, (c) 30m, see Figures 37 and 38.

**Figure 37:**
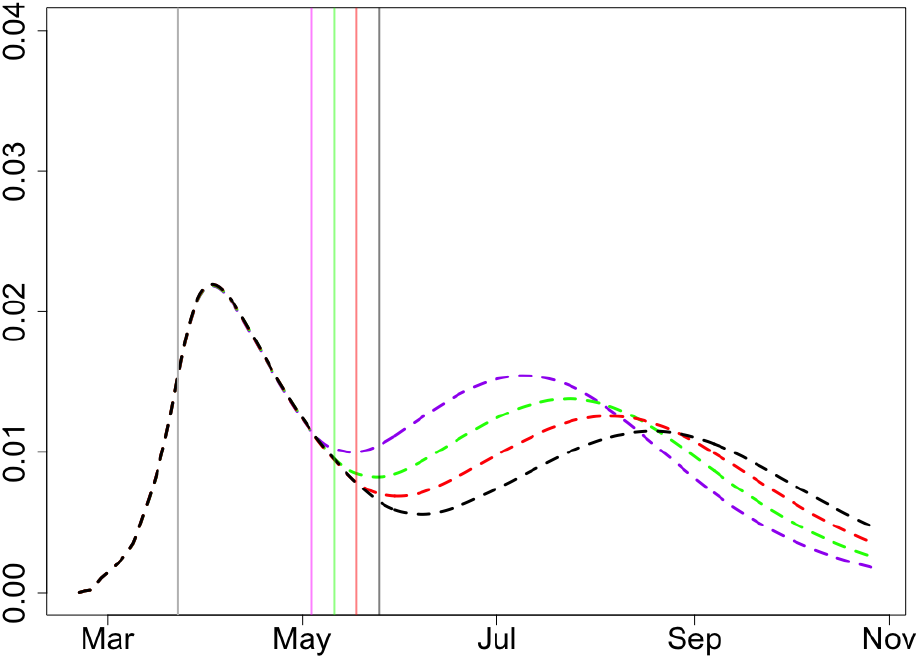
Proportion of infectious, UK, Scheme 2.

**Figure 38:**
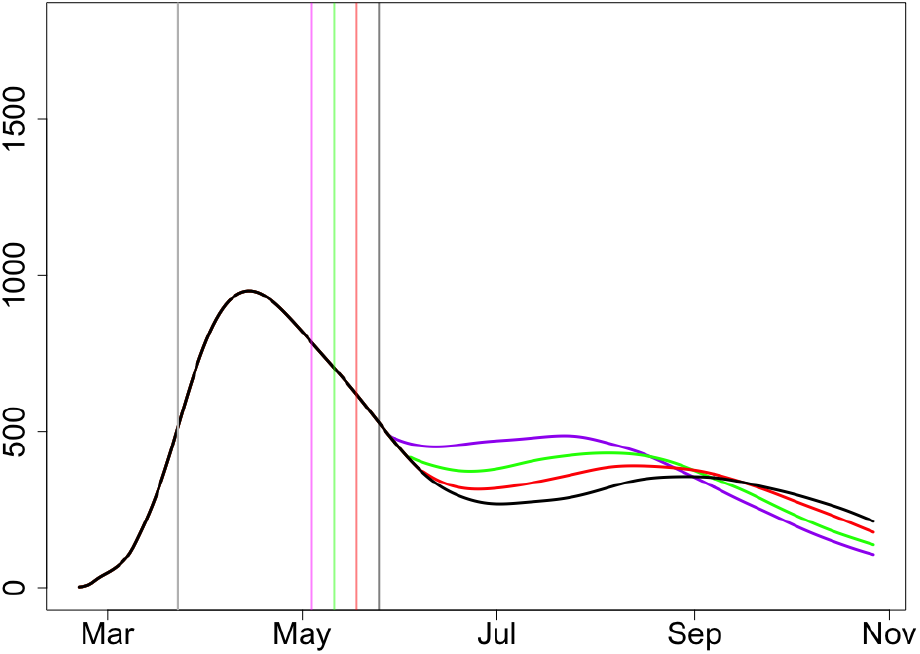
Expected deaths, UK, Scheme 2.

**Scheme 3**. (a) - 6m, (b) - 45m, (c) 15m, see Figures 39 and 40.

**Figure 39:**
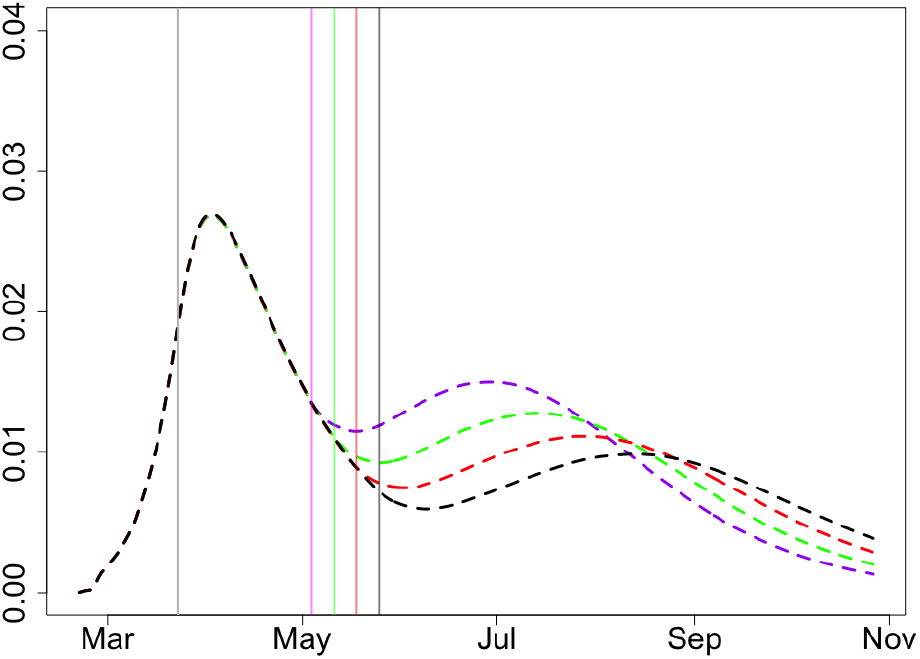
Proportion of infectious, UK, Scheme 3.

**Figure 40:**
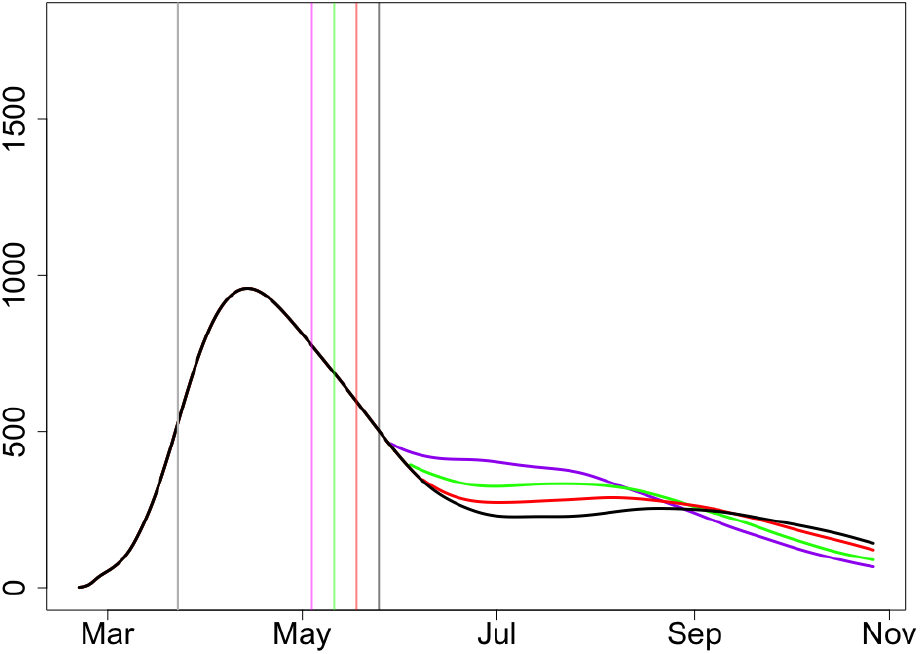
Expected deaths, UK, Scheme 3.

In Scheme 1 we used the same overall mortality rate as in all other figures, 0.66%. However, for Scheme 2 (which seems to be more realistic from the view-point of the demographics of the UK), we had to adjust the mortality rate to 0.44% = 2*/*3*×*0.66% to be consistent with the current mortality data. For the Scheme 3, we adjust the mortality rate to 0.363% = 0.55*×*0.66%

In Scheme 1, the percentage of infected by March 23 is only 2.65%, where 2.65 ≃ (10 *·* 8 + 5 *·* 8 + 1 *·* 60)*/*66 is uniquely determined by the scenario considered. In Scheme 2, this percentage is slightly higher: 3.63 ≃ (10 *·* 6 + 5 *·* 30 + 1 *·* 30)*/*66. In Scheme 3, it is 4.5 ≃ (10 *·* 6 + 5 *·* 45 + 1 *·* 15)*/*66.

In Scheme 1, despite considering a very pessimistic scenario, the second wave is much less pronounced (in terms of the expected number of deaths) than the first one. Note also that we assume that there is no vaccine and no progress with the way of treatment of the disease. In Schemes 2 and 3 there is no second wave. In all three schemes and all four lock-down lift dates, the number of new ICU patients does not exceed 5,000.

*For all three schemes considered, the total numbers of expected deaths are within 2% of each other, for the four dates of lifting the lock-down*. This is not very easy to deduce from Figures 36, 38 and 40 but please note that the epidemic will continue about 6 weeks longer for the lock-down lift date May 25 than for May 4; in the case of May 25, the epidemic is running much slower and 3 week difference in *t*_2_ becomes about 6 weeks at the end.

One of the strategies for managing the epidemic is based on the assumption that there will be a vaccine ready for a common use some time in October. It is therefore worthwhile to compare the scenarios in terms of the expected numbers of deaths occurred before November 1. For Schemes 2 and 3, which we consider rather most realistic, the total numbers of deaths occurred before November 1 for the four considered dates of lifting the lock-down differ by less than 5%. For Scheme 1, these numbers differ by less than 8%.

## Part II. Description of the main principles of the model for modeling the COVID-19 epidemic in the UK

The primary objective of our work is construction of a reliable, robust and interpretable model that describes the COVID-19 epidemic under different control regimes. We calibrate this model against currently available data about the epidemic and main recommendations of experts concerning the choice of parameters characterising virus behaviour. We discuss main scenarios of development of the epidemic and give recommendations about the exit strategy. We provide results of sensitivity studies and argue that currently acceptable SIR-based epidemiological models have serious limitations in such studies.

### Introduction

Coronavirus COVID-19 spreads through the population mostly based on social contact. To gauge the potential for widespread contagion, to cope with associated uncertainty and to inform its mitigation, more accurate and robust modelling is centrally important for policy making.

We provide a flexible modelling approach that increases the accuracy with which insights can be made. We use this to analyse different scenarios relevant to the COVID-19 situation in the UK. We present a stochastic model that captures the inherently probabilistic nature of contagion between population members. The computational nature of our model means that spatial constraints (e.g., communities and regions), the susceptibility of different age groups and other factors such as medical pre-histories can be incorporated with ease. We analyse different possible scenarios of the COVID-19 situation in the UK. Our model is robust to small changes in the parameters and is flexible in being able to deal with different scenarios.

Decision-makers get serious benefits from using better and more flexible models as they can avoid nuanced lock-downs, better plan the exit strategy based on local population data, different stages of the epidemic in different areas, making specific recommendations to specific groups of people; all this resulting in a lesser impact on economy, improved forecasts of regional demand upon hospitals allowing for intelligent resource allocation.

Our approach goes beyond the convention of representing the spread of an epidemic through a fixed cycle of susceptibility, infection and recovery (SIR), see e.g. [4, 5]. These SIR-based models, unlike our model, are not flexible enough and are also not stochastic and hence should be used with caution. Our model allows both heterogeneity and inherent uncertainty to be incorporated. Due to the scarcity of verified data, we draw insights by calibrating our model using parameters from other relevant sources, including agreement on average (mean field) with parameters in SIR-based models.

We use the model to assess parameter sensitivity for a number of key variables that characterise the COVID-19 epidemic. We also test several control parameters with respect to their influence on the severity of the outbreak. Our analysis shows that due to inclusion of spatial heterogeneity in the population and the asynchronous timing of the epidemic across different areas, the severity of the epidemic might be lower than expected from other models.

We model the development of the COVID-19 epidemic in the UK under different scenarios of handling the epidemic. There are many standard epidemiological models for modelling epidemics, see e.g. [4, 5]. In this work, we develop a generic stochastic model which has the following features:

- it can take into account spatial heterogeneity of the population and heterogeneity of development of epidemic in different areas;
- it allows the use of time-dependent strategies for analysing the epidemics;
- it allows taking into account special characteristics of particular groups of people, especially people with specific medical pre-histories and elderly.

Standard epidemiological models such as SIR/SEIR/SEIRS have some limitations and are not easily applicable for any study that requires the use of the features above. In particular, for influenza the mortality changes less significantly with age in comparison to coronavirus; hence common influenza models do not give much insight in modelling the COVID-19 epidemic. The report [6], which is widely considered as the main document specifying the current epidemic in the UK, is almost entirely based on the use of standard epidemiological models and hence the conclusions of [6] seem to be lacking specifics related to important issues of the study such as heterogeneity of development of epidemic at different locations and even flexible use of different death rates across different ages. Unreliability of COVID-19 data, including the numbers of COVID cases and COVID deaths, is a serious problem; it is widely discussed in media.

Despite the SIR models including the model of [6] have been heavily criticised [7], in view of the absence of reliable data, when choosing the main parameters of our model we calibrate it so that its mean field version approximately reproduces the same output as SIR-based models of [6, 8] and we use the parameter values consistent with [6, 8]. Also, the main notation (which sometimes does not look natural from a statistician’s point of view) is taken from [8].

We model further development of the COVID-19 epidemic in the example of the UK given the current data and assuming different scenarios of handling the epidemic. We incorporate all available to us (as to April 10, 2020) knowledge about parameters characterising the behaviour of the virus and the illness induced by it. It will take a long time before we know more precise data and facts about this pandemic.

We include assessments of the sensitivity of our model, using Julia programming language [9]; most of the previous epidemiological models do not allow this kind of sensitivity assessment as they have far too many parameters and very rigid conditions about the probability distributions involved. We believe the sensitivity studies are very important and should become the standard in this area.

The main practical findings from this research, with respect to the UK COVID-19 epidemic, are the following:

- very little gain, in terms of the projected hospital bed occupancy and expected numbers of death, of continuing the lock-down beyond April 13, provided the isolation of older and vulnerable people continues and the public carries on some level of isolation in the next 2-3 months, see Section 8.1;
- isolation of the group of vulnerable people during the next 2-3 months should be one of the main priorities, see Section 8.2;
- it is of high importance that the whole population carries on some level of isolation in the next 2-3 months, see Section 8.6;
- the timing of the current lock-down seems to be very sensible in areas like London where the epidemic has started to pick up by March 23; in such areas the second wave of epidemic is not expected, see figures in Sections 7.2 and 8;
- in the UK, the epidemic is expected to almost finish in July but a second wave may be expected in the areas where the first wave is almost absent, see Section 8.4.

For more findings, see conclusions at the end of subSections of Sections 8 and 9.

### 5 The model

#### 5.1 Variables and parameters

- *t* - time (in days)
- *t*_1_ - the lock-down time (in the case of the UK, *t*_1_ = March 23)
- *t*_2_ - time when the lock-down finishes
- *N* - population size
- *G* - sub-population (e.g. a group of people aged 70+)
- *n* - size of group *G*
- *α* = *n/N* (in the case of the UK and *G* consisting of people aged 70+, *α* = 0.132)
- *r*_*G*_ - average mortality rate in the group *G*
- *r*_*other*_ - average mortality rate for the rest of population
- *I*(*t*) - number of infectious at time *t*
- *S*(*t*) - number of susceptible at time *t*;
- 1*/σ*_*M*_ - average time for recovery in mild cases
- 1*/σ*_*S*_ - average time until recovery (or death) in severe cases
- *λ*_*I*_ - mean of the incubation period during which an infected person cannot spread the virus
- *δ* - the probability of death in severe cases
- *R*_0_ - reproductive number (average number of people who will capture the disease from one contagious person)
- *R*_1_ - reproductive number during the lock-down (*t*_1_ ≤ *t < t*_2_)
- *R*_2_ - reproductive number after the lock-down finishes (*t* ≥ *t*_2_)
- *c* - the degree of separation of vulnerable people
- *x* - the proportion of infected at the start of the lock-down

#### 5.2 Values of parameters and generic model

The reproductive number *R*_0_ is the main parameter defining the speed of development of an epidemic. There is no true value for *R*_0_ as it varies in different parts of the UK (and the world). In particular, in rural areas one would expect a considerably lower value of *R*_0_ than in London. Authors of [6] suggest *R*_0_ = 2.2 and *R*_0_ = 2.4 as typical; the authors of [8] use values for *R*_0_ in the range [2.25, 2.75]. We shall use the value *R*_0_ = 2.5 as typical, which, in view of the recent data, looks to be a rather high (pessimistic) choice overall. However, *R*_0_ = 2.5 seems to be an adequate choice for the mega-cities where the epidemics develop faster and may lead to more causalities. In rural areas, in small towns, and everywhere else where social contacts are less intense, the epidemic is milder.

The flow-chart in Figure 41 describes the process of illness. We assume that the person becomes infected *τ*_*I*_ days after catching the virus, where *τ*_*I*_ has Poisson distribution with mean of *λ*_*I*_ days. Parameter *λ*_*I*_ defines the mean of the incubation period during which the person cannot spread the virus. Recommended value for *λ*_*I*_ is *λ*_*I*_ = 5.

**Figure 41:**
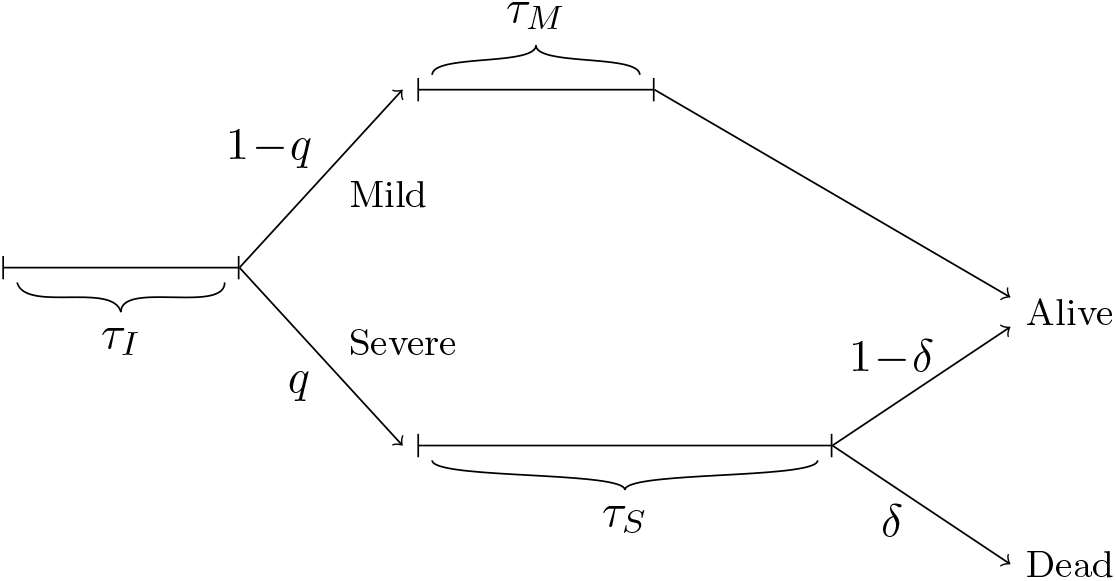
Flow-chart for the process of illness.

We then assign probabilities *q* and 1 − *q* for a person to get a severe or mild case correspondingly. The value of *q* depends on whether the person belongs to the group *G* (then *q* = *q*_*G*_) or the rest of population (in this case, *q* = *q*_*other*_). In Section 5.3 below we will relate the probabilities *q*_*G*_ and *q*_*other*_ to the mortality rates in the two groups.

In a mild case, the person stays infectious for *τ*_*M*_ days and then discharges alive (that is, stops being infectious). We assume that the continuous version of *τ*_*M*_ has Erlang distribution with shape parameter *k*_*M*_ and rate parameter *λ*_*M*_, the mean of this distribution is *k/λ*_*M*_ = 1*/σ*_*M*_ (in simulations, we discretise the numbers to their nearest integers). We use values *k*_*M*_ = 3 and *λ*_*M*_ = 0.6 so that *Eτ*_*M*_ = *k*_*M*_ */λ*_*M*_ = 1*/σ*_*M*_ = 5. The variance of *τ*_*M*_ is 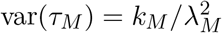; for *k*_*M*_ = 3 and *λ*_*M*_ = 0.6 we have var(*τ*_*M*_) ≃ 8 which seems to be a reasonable value.

In a severe case, the person stays infectious for *τ*_*S*_ days. The continuous version of *τ*_*S*_ has Erlang distribution with shape parameter *k*_*S*_ and rate parameter *λ*_*S*_. The mean of this distribution is *k*_*S*_*/λ*_*S*_ = 1*/σ*_*S*_. We use values *k*_*S*_ = 3 and *λ*_*S*_ = 1*/*7 so that *Eτ*_*S*_ = *k*_*S*_*/λ*_*S*_ = 21, in line with the current knowledge, see e.g. [10, 11]. The variance of *τ*_*S*_ is 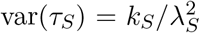; for *k*_*M*_ = 3 and *λ*_*S*_ = 1*/*7 the standard deviation of the chosen Erlang distribution is approximately 12, which is rather large and reflects the uncertainty we currently have about the period of time a person needs to recover (or die) from COVID-19.

In a severe case, the person dies with probability *δ >* 0 on discharge. The relation between *δ*, the average mortality rates, the probabilities *q*_*S*_ for the group *G* and the rest of population is discussed in Section 5.3.

Note that the use of Erlang distribution is standard for modelling similar events in reliability and queuing theories, which have much in common with epidemiology. SIR-based models do not allow the use of such distributions and this is one of the reasons why SIR models cannot be considered as flexible and reliable.

#### 5.3 Probability of getting a severe case, probability of death in case of severe case *δ* and mortality rates

In this Section, we relate the probability of getting severe case from group *G* and the rest of population to the average mortality rates.

We define *G* as the group of vulnerable people. In the computations below, we assume that *G* consists of people aged 70+. We would like to emphasise, however, that there is still a lot of uncertainty on who is vulnerable. Moreover, it is very possible that the long term effect of the virus might cause many extra morbidities in the coming years coming from severe cases who recover.

As we use UK epidemic as our main example, we use the common split of the UK population into following age groups:

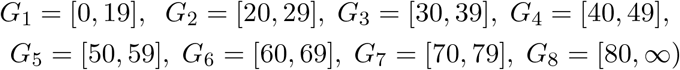

and corresponding numbers *N*_*m*_ (*m* = 1, …, 8; in millions) taken from [12]

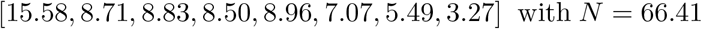

The probabilities of death for group *G*_*m*_, denoted by *p*_*m*_, are given from Table 1 in [6] and replicated many times by the BBC and other news agencies are:

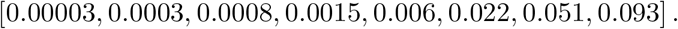

Unfortunately, these numbers do not match the other key number given in [6]: the UK average mortality rate which is estimated to be about 0.9%. As we consider the value of the UK average mortality rate as more important, we have multiplied all probabilities above by 0.732 to get the average mortality rate to be 0.9%.

Moreover, recent COVID-19 mortality data shows that the average mortality rates, especially for younger age groups, are significantly lower than the ones given above. As we do not have reliable sources for the average mortality rates, we shall use the data from [6] adjusted to the average mortality rate to be 0.9%. As we shall be getting better estimates of the average mortality rates, the model can be easily adjusted to them.

It is important to note that different authors disagree on the values of mortality *r* for the COVID-19; see, for example, [13]. UK’s experts believe *r* ≃ 0.009 [6], WHO sets the world-wide mortality rate at 0.034, the authors of [14] believe *r* is very small and could be close to 0.001, an Israeli expert D. Yamin sets *r* = 0.003, see [15]. Current development of the epidemic suggest that the overall (world-wide) value of *r* is not larger than *r* = 0.005 but it is too early to say. Note that in a particular country or even region the average mortality is different as it depends on the age structure of the county (region) and almost certainly on climatic conditions influencing virus activity.

If consider population data for the whole UK and define the group *G* as a union of groups *G*_7_ and *G*_8_, we have *n* = 5.49 + 3.27 = 8.76m and *α* = 8.76*/*66.41 *∼* 0.132. The mortality rate in the group *G* and for the rest of population are therefore

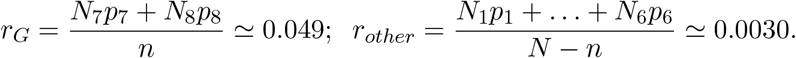

Consider now a random person from the group *G* who has got an infection. The probability he dies is *r*_*G*_. On the other hand, this is also the product of the probability that this person has a severe case (which is *q*_*G*_) and the probability (which is *δ*) that the person dies conditionally he/she has severe case. The same is true for the rest of population. Therefore,

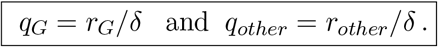

The COVID-19 epidemic in the whole of the UK is subject to different heterogeneities discussed in detail in [1], see also Section 6. The model above assumes homogeneity and hence cannot be applied to the whole of the UK and we feel it would be irresponsible to estimate the total death toll for the UK using such a model. Instead, we shall assume that we apply this model to the population of inner London with population size rounded to 3 million. It may be tempting to multiply our expected death tolls by 22=66/3 but this would give very elevated forecasts. Indeed, epidemic in inner London can be considered as the worst-case scenario and, in view of [1], we would recommend to down-estimate the UK overall death toll forecasts by a factor of 2 or even more.

Note that in this work we only discuss mortality directly associated with COVID-19. Recent reports point to many additional deaths associated with failure of patients, such as cancer patients, to get proper treatment.

#### 5.4 The number of infected at time *t*

To start with, in Figure 42 we consider an uninterrupted run of an epidemic with *R*_0_ = 2.5 in a homogeneous (one-group) population with the parameters set in Section 7.2. The starting time of an epidemic is unknown and is even hard to define as the first transmissions of the virus take random and perhaps long times. We start plotting the curves after 0.5% of the population are infected.

**Figure 42:**
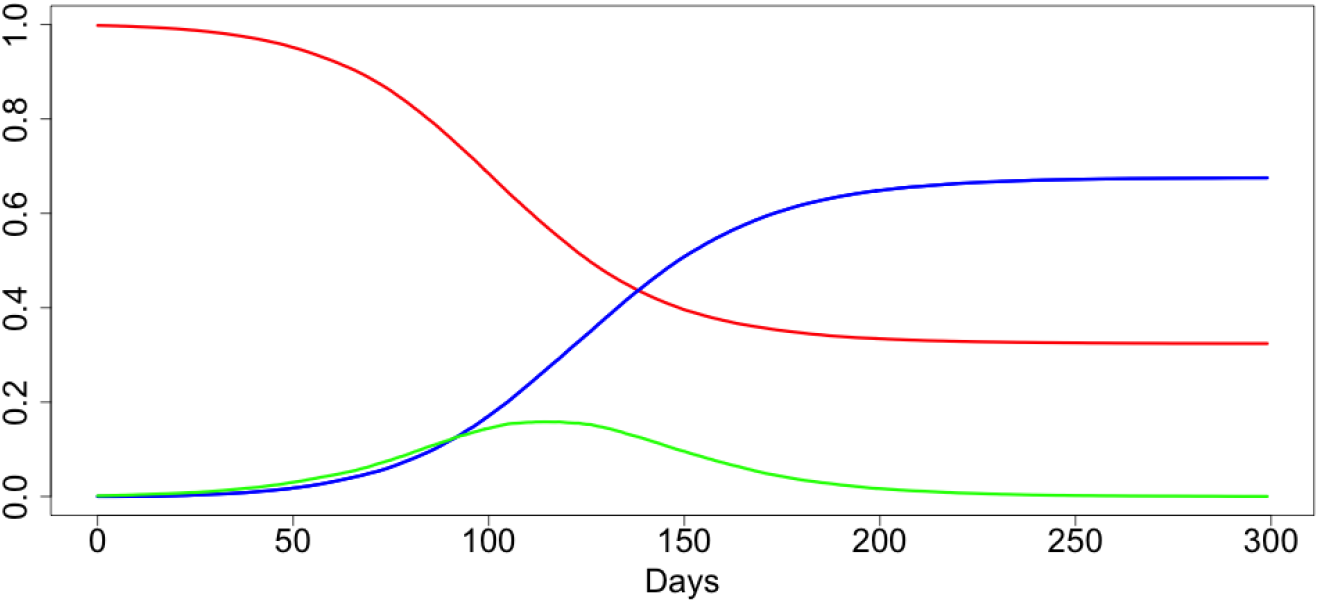
An uninterrupted run of a COVID-19 epidemic in homogeneous conditions.

In red colour, in Figure 42 and all plots below we plot values of *S*(*t*)*/N*, the proportion of people non-infected by time *t*. In blue, we plot *R*(*t*)*/N*, the proportion of people recovered from the disease (or dead) and in green we plot our main quantity of interest which is *I*(*t*)*/N*, the proportion of infected people at time *t*. The values of *R*(*t*) do not play any part in modelling and are plotted for information only. Plots similar to Figure 42 are usually provided to illustrate SIR-based model outputs.

One of the main targets of decision-makers for dealing with epidemics like COVID-19 can be referred to as ‘flattening the curve’, where ‘the curve’ is *I*(*t*)*/N* or any of its equivalents and ‘flattening’ roughly means ‘suppressing the maximum’. We shall consider this in the next Sections. From the value *I*(*t*), we can roughly estimate the required number of hospital beds and even the expected number of deaths at time *t*. In Section 7.2 we discuss more accurate estimates for these quantities which use the splitting of the population on the two groups.

### 6 Spatial heterogeneity of the population and heterogeneity of epidemic development in different areas

The purpose of this Section is to demonstrate that heterogeneity of epidemic developments for different sub-populations has significant effect on ‘flattening the curve’.

Already when this paper was completed, the authors learned about a preprint [16] which uses SIR modelling to produce somewhat similar conclusions with more specifics for COVID-19 in the UK. However, SIR modelling using numerous age-classes and many equations for each age-class different across different parts of the country resulting in an astronomical number of parameters; this makes any sensitivity analysis to various parameters uncheckable. We believe that a combination of stochastic models like the present one with standard SIR-based models can help in significantly reducing the number of parameters while also adding more flexibility to a hybrid model.

Consider the following situation. Assume that we have a population consisting of *M* sub-populations (groups) *G*_*m*_ with similar demographic and social characteristics and these sub-populations are subject to the same epidemic which has started at slightly different times. Let the sizes of sub-population *G*_*m*_ be *N*_*m*_ with *N*_1_ + … + *N*_*m*_ = *N*. In all cases, the curves *I*_*m*_(*t*)*/N*_*m*_ are shifted in time versions of the curve *I*(*t*)*/N* of Figure 42.

In Figures 43 and 44, *M* = 2 and the second epidemic started 50 days after the first one (incubation period is set to be 0). In Figures 45 and 46, *M* = 10 and each next epidemic cycle has started 7 days after the previous one. In all cases, we evidence the significant ‘flattening the curve’ phenomenon. In Figures 43– 46, the green line is used for the resulting curves *I*(*t*)*/N*. The maximal values of *I*(*t*)*/N* in the examples depicted in Figures 43–46 are 0.124, 0.136, 0.134 and 0.132, respectively. This is significantly lower than the value 0.163 for the original curve. In the assumptions of Example 1, these would respectively lead to the maximum expected number of deaths 57.7, 63.3, 62.9 and 62.7.

**Figure 43:**
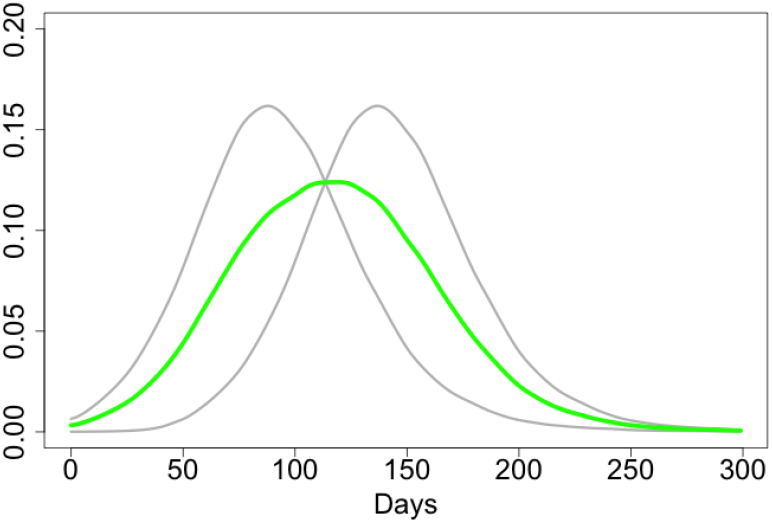
*M* = 2, *N*_1_ = *N*_2_.

**Figure 44:**
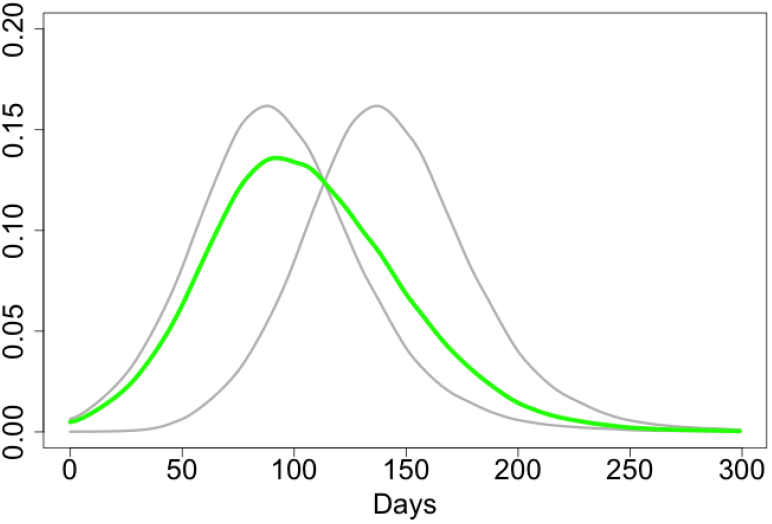
*M* = 2, *N*_1_ =3*N*_2_.

**Figure 45:**
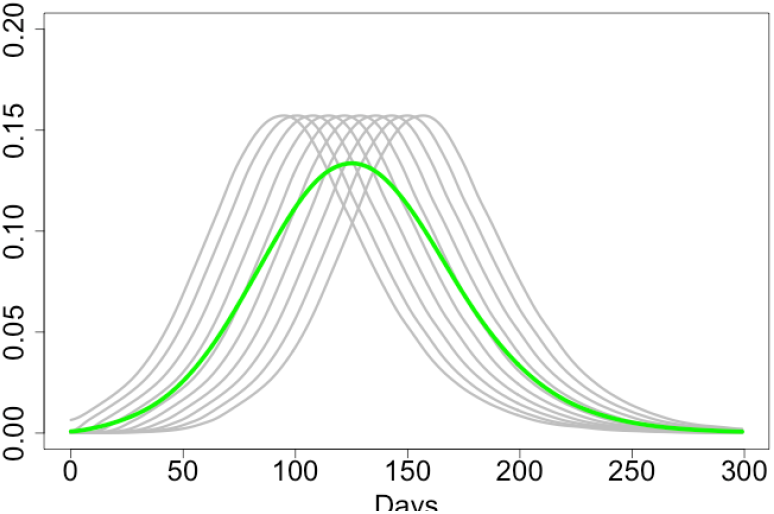
*M =* 10, 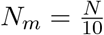 (*m* = 1; …, 10)

**Figure 46:**
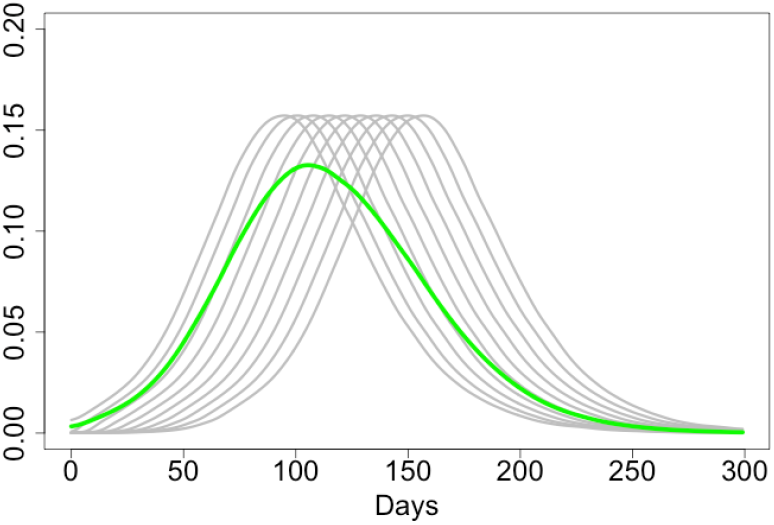
*M* = 10,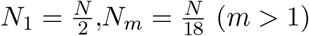.

Extra heterogeneity of different epidemics caused by the social and demographic heterogeneity of the sub-populations would further ‘flatten the curve’. The following conclusions are in line with what the Israeli expert D. Yamin states [15] and agree with the main conclusions of the paper [17] which is rather critical towards standard epidemiological models.

**Conclusions**. *(a) Isolation of sub-populations at initial stages of an epidemic is very important for preserving heterogeneity of the epidemic and ‘flattenning the curve’; this flattening can be very significant. (b) Epidemiological models based on the assumption of a homogeneous population but applied for populations consisting of heterogeneous sub-populations may give completely misleading results*.

### 7 Main scenarios for modelling the epidemic with lock-down

In this Section we formulate two main scenarios and explain the main figures. In Section 8 we study sensitivity with respect to all model parameters.

#### 7.1 Two plots for the main set of parameters, no intervention

As the main set of parameters for the main model we use the following:

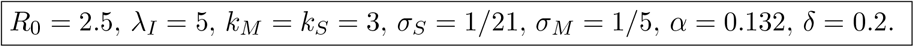

Note that in Section 7.2 we shall introduce four further parameters which will describe the lock-down of March 23 and the situation at the exit from this lock-down.

In Figure 47 we plot, in the case with no intervention, proportions of infected at time *t* (either overall or in the corresponding group) and use the following colour scheme:

- Solid green: *I*(*t*), the proportion of infected in overall population at time *t*.
- Purple: *I*_*G*_(*t*), the proportion of infected in group *G* at time *t*
- Dashed green: the proportion of infected in the rest of population.

**Figure 47:**
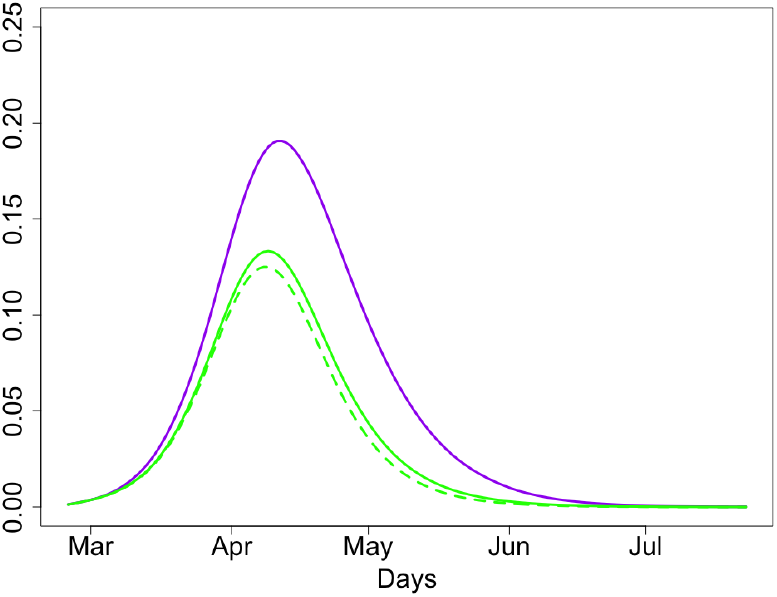
Proportions of people infected at time *t* (overall, group *G* and the rest of population).

Knowing proportion of infected at time *t* is important for knowing the danger of being infected for non-infected and non-immune people.

In Figure 48 we plot, in the case of no intervention, proportions of people with severe case of disease at time *t* and proportions of such people discharged at time *t*; there are separate plots for the group *G* and the rest of population.

- Solid purple: proportion of people from group *G* with severe case at time *t*.
- Solid green: proportion of people from the rest of population with severe case at time *t*.
- Dashed purple: proportion of people from group *G* died at time *t*, multiplied by 100.
- Dashed green: proportion of people from the rest of population died at time *t*, multiplied by 1000 (so that the curve can be seen in the figure).

**Figure 48:**
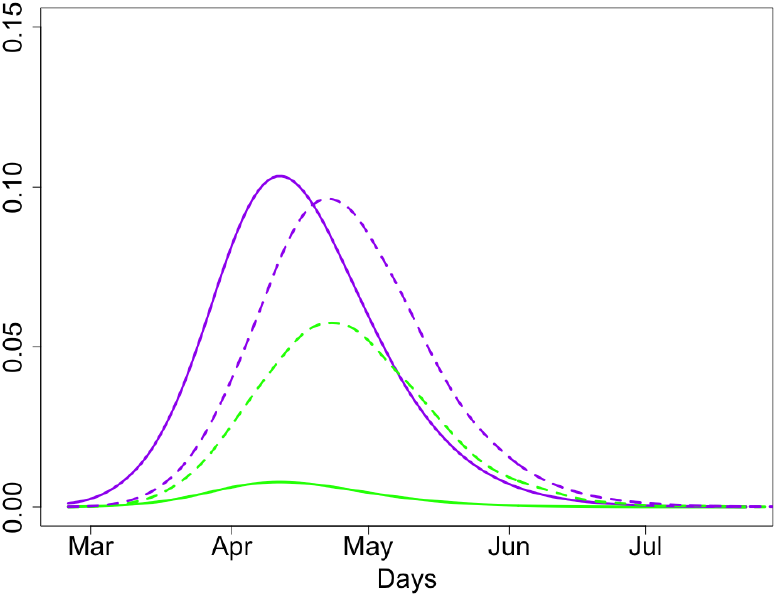
Proportions of people with severe case of disease at time *t* (solid lines) and died at time *t* (dashed lines).

Proportions of people with severe case at time *t* is the main characteristic needed for planning NHS workload. The proportion of people with severe case from group *G* discharged at time *t* is proportional to the expected number of deaths after multiplying this number by *δ* and the size of the group *G*; similar calculations can be done for the rest of population. Note that in these calculations we do not take into consideration an extremely important factor of hospital bed availability are at time *t*. Currently, we do not have a model for this.

We provide the values of the overall expected death toll. In the case of no lock-down, this toll would be 24(7+17)K, where the first number in the bracket correspond to the rest of population and the second one to the group *G*.

#### 7.2 Key plots for the main set of parameters, the case with lock-down

In this Section, we study the scenario where we have made an intervention on *t*_1_ = March 23 by reducing *R*_0_ to *R*_1_ *< R*_0_ and then terminating the lock-down on *t*_2_ = April 22.

As we do not know at which stage of the epidemic the lock-down has started, we define it through the parameter *x* ∈ (0, 1) such that *S*(*t*_1_)*/N* = *x*.

After lifting the lock-down, we assume that people from the group *G* are still isolated but the rest of population returns back to normal life. However, it is natural to assume that the reproduction number *R*_2_ for *t* ≥ *t*_2_ is smaller that the initial value of *R*_0_. To denote the degree of isolation of people from *G* for *t* ≥ *t*_2_, we assume that for *t* ≥ *t*_2_ the virus is transmitted to people in such a way that, conditionally a virus is transmitted, the probability that it reaches a person from *G* is *p* = *cα* with 0 *< c* ≤ 1. Our main value for *c* is 1*/*4. This means that for *t* ≥ *t*_2_, under the condition that a virus is infecting a new person, the probabilities that this new person belongs to *G* is 1*/*5.

Measuring the level of compliance in the population and converting this to simple epidemiological measures *c, R*_1_ and *R*_2_ and is hugely complex problem which is beyond the scope of this paper.

Summarizing, there are five parameters modelling the lock-down:

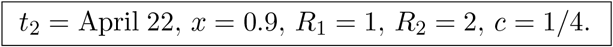

Figures 49 and 50 similar to Figures 47 and 48 but computed for the case of intervention. The colour schemes in Figures 49 and 50 are exactly the same as in Figures 47 and 48, respectively. The two grey lines in Figures 49 and 50 mark the intervention times. Note different scales in the *y*-axis in Figures 49 and 50 in comparison to Figures 47 and 48

**Figure 49:**
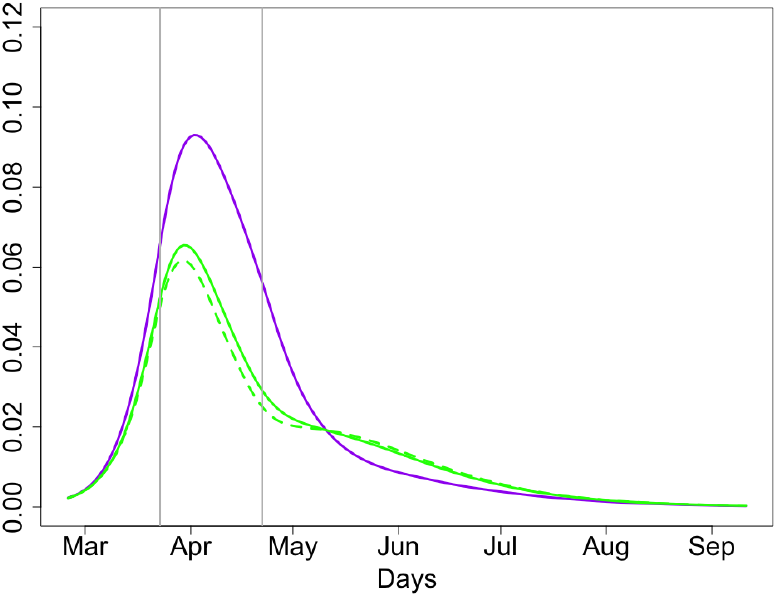
Proportions of people infected at time *t* (overall, group *G* and the rest of population).

**Figure 50:**
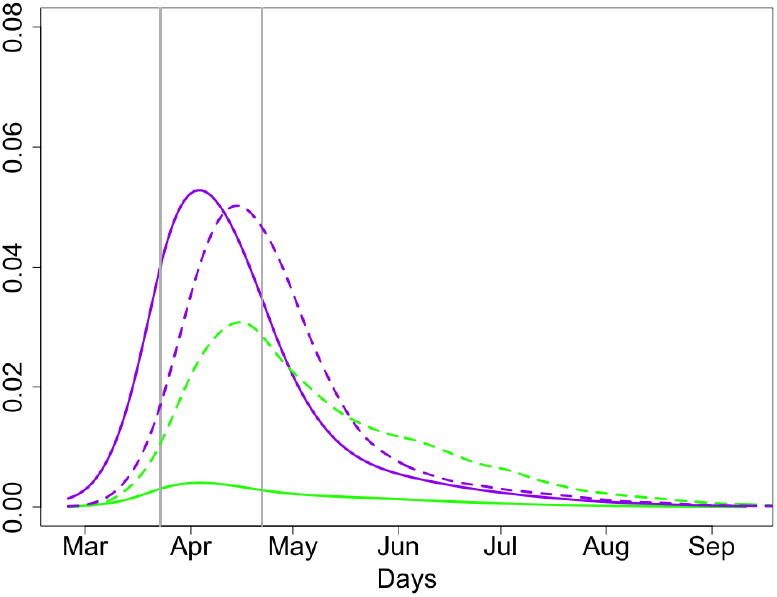
Proportions of people with severe case of disease at time *t* (solid lines) and discharged at time *t* (dashed). Expected death toll: 15K.

We will study sensitivity of the curves *I*(*t*) and *I*_*G*_(*t*), plotted in Figures 49 and again in Figure 51 for the same parameters, with respect to all parameters of the model. In Figure 52 we plot average death numbers (purple for group *G* and green for the rest of the population) in the case of the inner London as explained at the end of Section 5.3. Note that these numbers do not take into account the important factor of hospital bed availability and various heterogeneities, as explained in [1].

**Figure 51:**
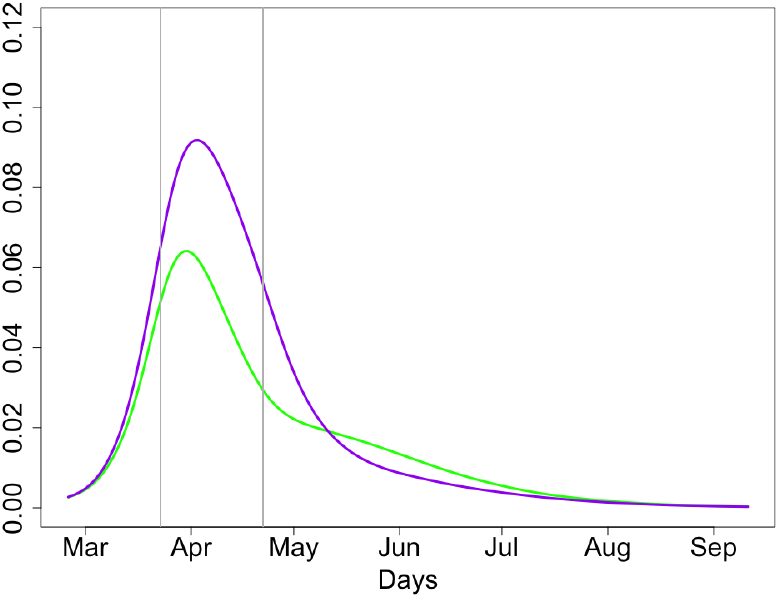
Proportions of people infected at time *t* (group *G* and overall).

**Figure 52:**
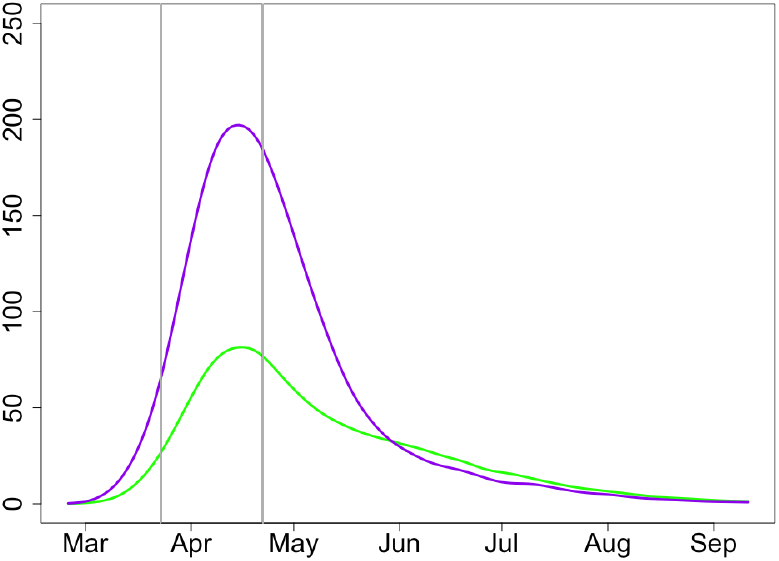
Expected deaths at time *t* at group *G* and the rest of population.

### 8 Sensitivity to main parameters of the model and exit strategy

We have selected Figures 51 and 52 as the main figures. The next pairs of figures show sensitivity of the model with respect to different parameters in the model. All these pairs of figures are similar Figures 51 and 52 except one of the parameters is changing. Moreover, all the curves from Figures 51 and 52 are reproduced in the pairs of figures below in the same colours.

The colour scheme in all figures below with even numbers is:

- Solid purple: *I*_*G*_(*t*)*/n*, the proportion of infected at time *t* from group *G*; main set of parameters;
- Solid green: *I*(*t*)*/N*, the proportion of infected at time *t* from the population; main set of parameters;
- Solid (and perhaps, dashed) red: *I*_*G*_(*t*)*/n* for the alternative value (values) of the chosen parameter;
- Solid (and perhaps, dashed) blue: *I*(*t*)*/N* for the alternative value (values) of the chosen parameter. The colour scheme in all figures below with odd numbers is:
- Solid purple: *D*_*G*_(*t*), the expected number of death at time *t* at group *G*; main set of parameters;
- Solid green: *D*_*other*_(*t*), the expected number of death at time *t* for the rest of population; main set of parameters;
- Solid (and perhaps, dashed) red: *D*_*G*_(*t*) for the alternative value (values) of the chosen parameter;
- Solid (and perhaps, dashed) blue: *D*_*other*_(*t*) for the alternative value (values) of the chosen parameter.

### 8.1 Sensitivity to *t*_2_, the time of lifting the lock-down

In Figures 53 and 54, we have moved the time *t*_2_, the time of lifting the lock-down restrictions, 9 days forward so that the lock-down period is 21 days rather than 30 days as in the main scenario. The solid red and blue colours in Figures 53 and 54 correspond to *t*_2_ = April 13 and the middle grey vertical line marks April 13.

**Figure 53:**
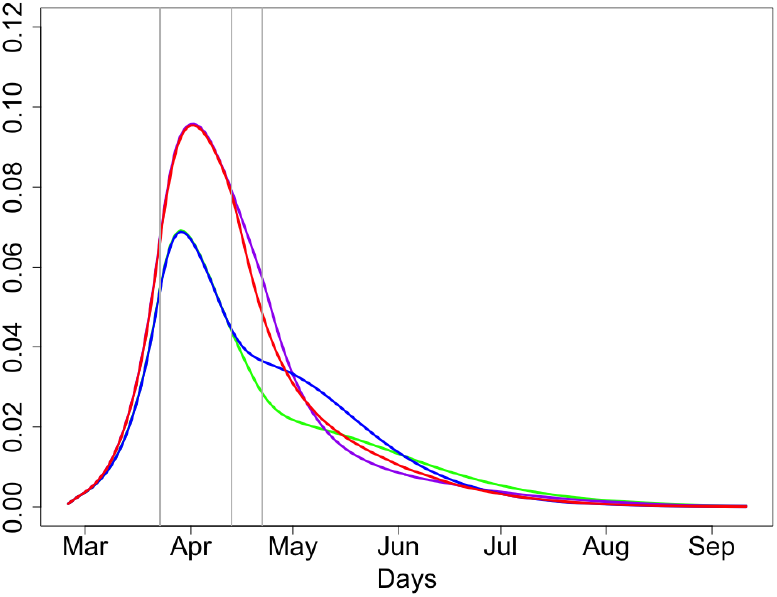
Proportions of people infected at time *t*; *t*_2_ = April 13, 22.

**Figure 54:**
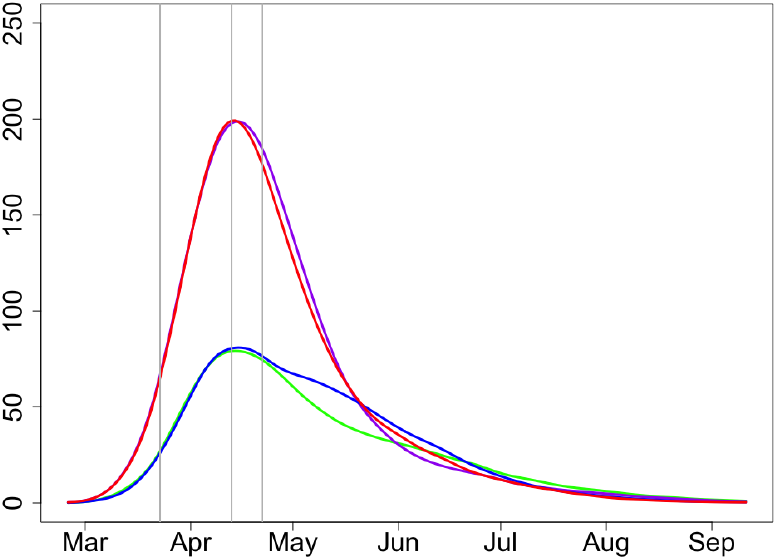
Expected deaths at time *t* at group *G* and the rest of population; *t*_2_ = April 13, 22.

Consider first Figure 53 showing proportions of infected people and reflecting the demand for hospital beds. Since in the scenario *t*_2_ = April 13 we start isolating people from *G* earlier than for *t*_2_ = April 22, the number of infected from group *G* (red solid line) is slightly lower during the end of April – beginning of May for *t*_2_ = April 13 (purple solid line). The total number of infections from the rest of population is clearly slightly higher for *t*_2_ = April 13 (blue line) than for *t*_2_ = April 22. Taking into account the fact that infected people from *G* would on average require more hospital beds than the rest of population, the expected number of hospital beds required for the whole population stays approximately the same during the whole epidemic which has to be almost over in July.

Figure 54, showing expected deaths numbers for both scenarios with *t*_2_ = April 13 and *t*_2_ = April 22, show similar patterns concluding that there is very little gain in keeping the lock-down, especially taking into account huge economic loss caused by every extra day of the lock-down [18]. Overall, the total expected death is higher but the difference can be considered as very small. Expected deaths tolls for *t*_2_ = April 22 and *t*_2_ = April 13 are 15(5.2+9.8)K and 15.3(5.6+9.7)K, respectively.

**Conclusion**. *The lock-down started on March 23 has slowed the speed of COVID-19 epidemic. However, there is very little gain, in terms of the projected hospital bed occupancy and expected numbers of death, of continuing the lock-down beyond April* 13.

#### 8.2 Sensitivity to *c*, the degree of separation of people from group *G*

The main subject of our work [1] was the argument that isolating vulnerable people makes a huge effect on expected mortality from the epidemic. In this Section, we continue the discussion started by us in [1] and illustrate this scenario using the style of figures which we use in this paper.

In Figures 55 and 56, solid and dashed line styles (for red/blue colours) correspond to *c* = 1 and *c* = 0.5 respectively. By increasing *c* we significantly increase the death toll in the group *G*. It is clear that the value of *c* measures the degree of isolation of people from *G* and has negligible effect on the rest of population; this is clearly seen in both figures. On the other hand, the value of *c* has very significant effect on the people from *G*.

**Figure 55:**
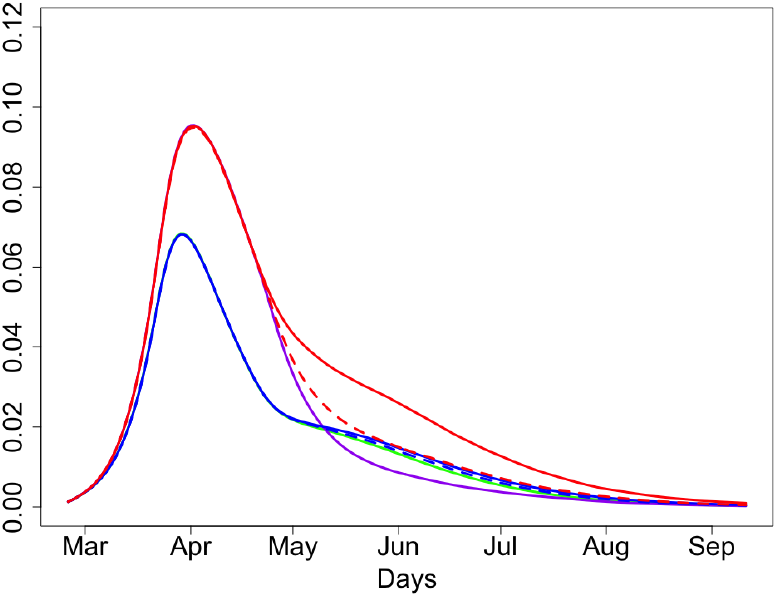
Proportions of people infected at time *t*; *c* = 0.25, 0.5, 1.

**Figure 56:**
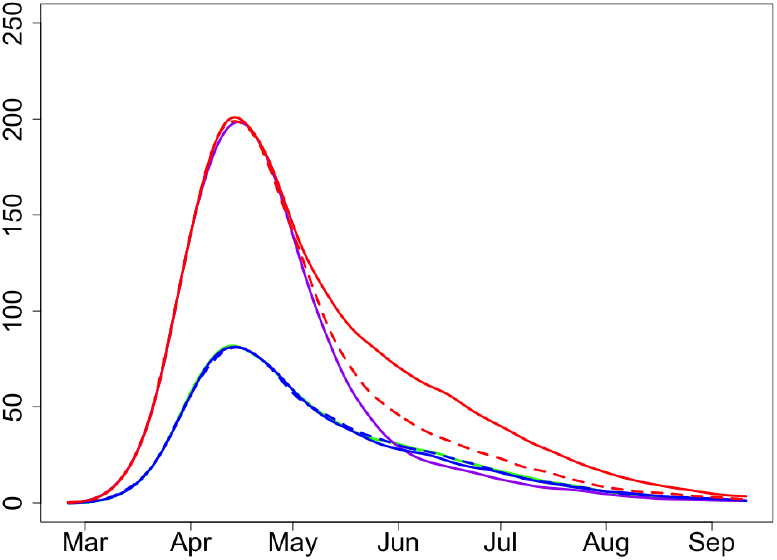
Expected deaths at time *t* at group *G* and the rest of population; *c* = 0.25, 0.5, 1 *c* = 0.25.

Consider first Figure 55 giving proportions of infected people. One can clearly see that the purple line (group *G, c* = 0.25) is much lower that the solid red line (group *G, c* = 1) for all the time until the end of the epidemic, from May until August. In June, in particular, the number of infected people from *G* with *c* = 0.25 is about 40% of the number of infected people from *G* with *c* = 1.

Curves on Figure 56, showing expected deaths at time *t* in group *G* and the rest of population, follow the same patterns (with about 2-week time shift) as the related curves of Figure 55 displaying the number of infected. In June and July we should expect only about 40% of the number of deaths in the scenario with isolation of people from *G* (*c* = 0.25) relative to the scenario with no special isolation for people from *G* (*c* = 1).

This is reflected in the expected deaths tolls for entire period of epidemic. Indeed, expected deaths tolls for *c* = 1 and 0.5 are 18.2(5.2+13)K and 16.2(5.2+11)K, respectively; compare this with 15(5.2+9.8)K for *c* = 0.25.

**Conclusion**. *The effect of the degree of isolation of people from group G in the aftermath of the lock-down is very significant*.

Note that we simply stress the importance of special measures for isolating the group of vulnerable people but not claiming it is practically possible with the current social care system.

#### 8.3 Sensitivity to *R*_0_

In view of recent data, the value of *R*_0_ is likely to be lower than 2.5, especially in small towns and rural areas. As Figures 57 and 58 demonstrate, in such places the epidemic will be significantly milder. The overall dynamics of the epidemic would not change much though.

**Figure 57:**
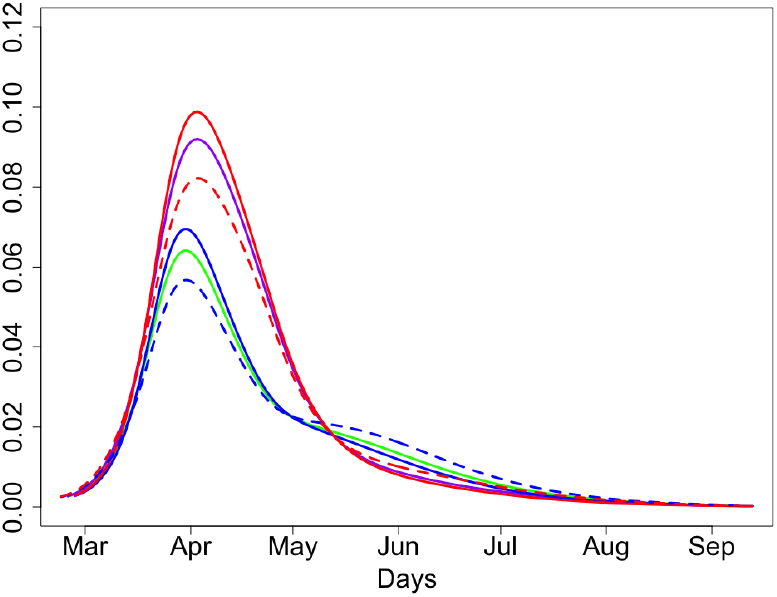
Proportions of people infected at time *t*; *R*_0_ = 2.3, 2.5, 2.7.

**Figure 58:**
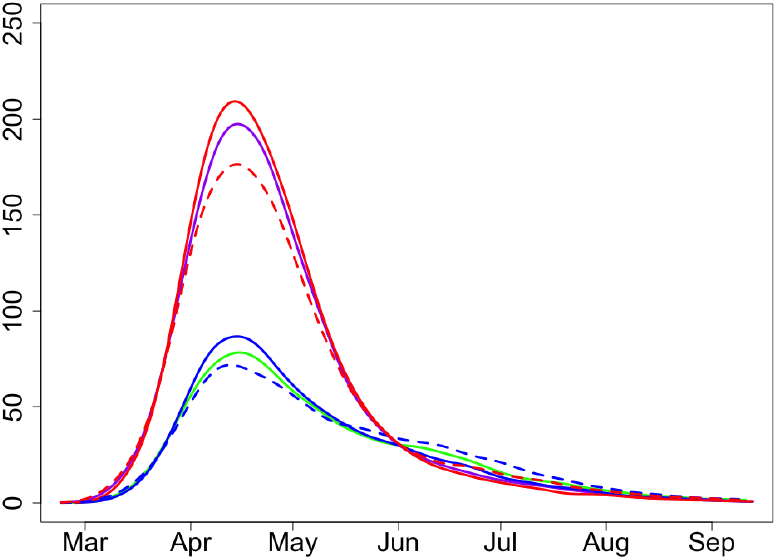
Expected deaths at time *t* at group *G* and the rest of population; *R*_0_ = 2.3, 2.5, 2.7.

In Figures 57 and 58, solid and dashed line styles (for blue/red colours) correspond to *R*_0_ = 2.3 and *R*_0_ = 2.7 respectively. Expected deaths tolls for *R*_0_ = 2.3, 2.5 and *R*_0_ = 2.7 are 14.5(5+9.5)K, 15(5.2+9.8)K and 15.5(5.3+10.2)K, respectively.

#### 8.4 Sensitivity to *x*, the proportion of susceptible at the start of the lock-down

In Figures 59 and 60, we use *x* = 0.8 and *x* = 0.9. Expected deaths toll for *x* = 0.8 is 17.3(5.3+12)K. This is higher than 15(5.2+9.8)K for *x* = 0.9. The fact that the difference is significant is related to the larger number of death for *x* = 0.8 in the initial period of the epidemic. In Figure 59, red and blue display the number of infected at time *t* for *x* = 0.8 for people from group *G* and the rest of population, respectively. Similarly, in Figure 60 the red and blue colours show the expected numbers of deaths in these two groups.

**Figure 59:**
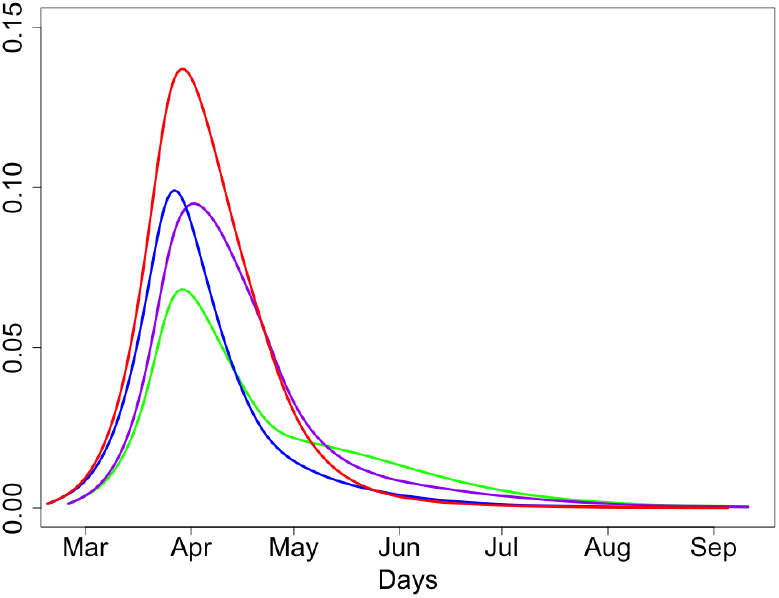
Proportions of people infected at time *t*; *x* = 0.8, 0.9.

**Figure 60:**
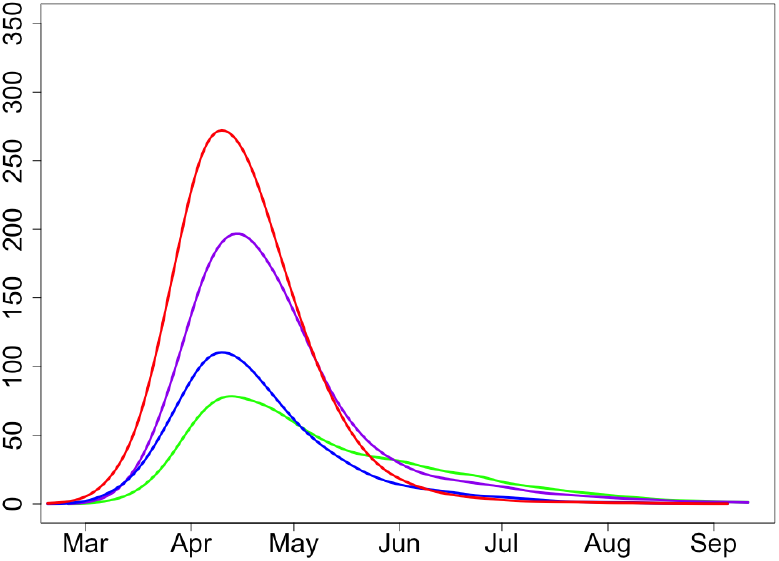
Expected deaths at time *t* at group *G* and the rest of population; *x* = 0.8, 0.9.

Figures 59 and 60 show that the timing of the lock-down has serious effect on the development of the epidemic. Late call for a lock-down (when *x* = 0.8) helps to slow down the epidemic and guarantees its fast smooth decrease but does not save as many people as, for example, the call with *x* = 0.9.

Note that in both cases, when *x* = 0.8 and *x* = 0.9, the second wave of the epidemic is not expected as by the time of lifting the lock-down, a large percentage of the population (about 25% in case *x* = 0.9 and almost 40% in case *x* = 0.8) is either infected or immune and the ‘herd immunity’ would follow shortly. In a certain sense, the decision of making a lock-down at around *x* = 0.9 seem to be a very sensible decision to make as this saves many lives and guarantees smooth dynamics of the epidemic with no second wave.

Although we need more evidence about immunity, in this paper we assume that infected people are not to be infected with COVID-19 again for at least several months as seems to be the case with other corona viruses. Also, this model only applies to the UK. All the results in this paper on the relationship between the *x* variable and the second wave only apply under the assumption of no international travel

Let us now consider more informative scenarios when *x* = 0.95 and *x* = 0.97; that is, when the lock-down is made early, see Figures 61,62 for *x* = 0.95, 0.9 and Figures 63,64 for *x* = 0.97, 0.9. With an early lock-down, there is a clear gain in hospital bed occupancy and expected number of death at the first stage of the epidemic. However, the second wave of the epidemic should be expected in both cases *x* = 0.95 and *x* = 0.97 with a peak at around 2 months after the first one, and the second peak could be higher than the first one. This can be explained by observing that, even after 2 months of an epidemic with large *x*, even with lock-down and strong isolation of the group *G* and relatively small reproductive number (recall *R*_2_ = 2), there is still a very large proportion of non-immune people available for the virus; a large part of these people is going to be infected even with smaller reproductive number. This prolongs the epidemics. Expected deaths toll for *x* = 0.95 is 13.8(5.5+8.3)K and for *x* = 0.97 it is 13.4(5.7+7.7)K. These numbers are naturally lower than 15(5.2+9.8)K for *x* = 0.9 but not by much.

**Figure 61:**
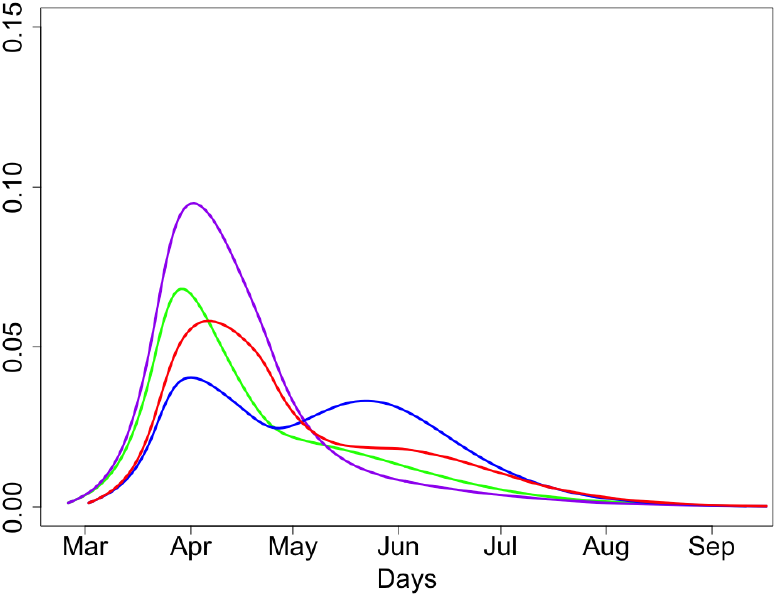
Proportions of people infected at time *t*; *x* = 0.9, 0.95.

**Figure 62:**
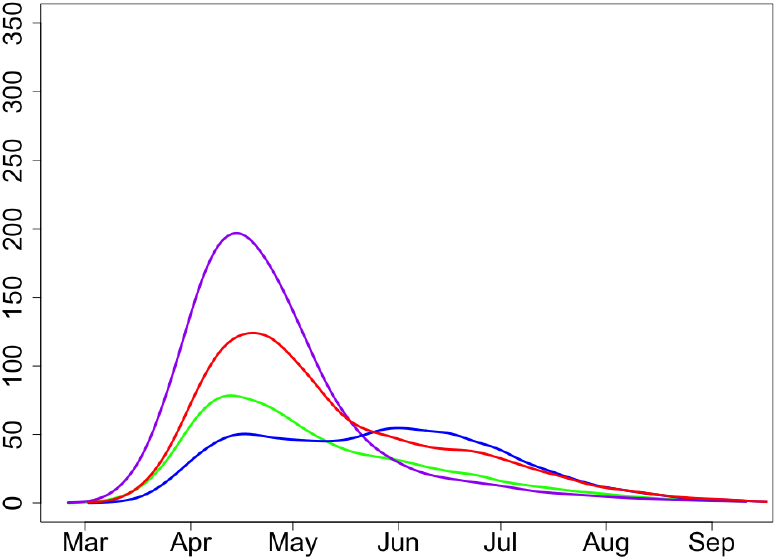
Expected deaths at time *t* at group *G* and the rest of population; *x* = 0.9, 0.95.

**Figure 63:**
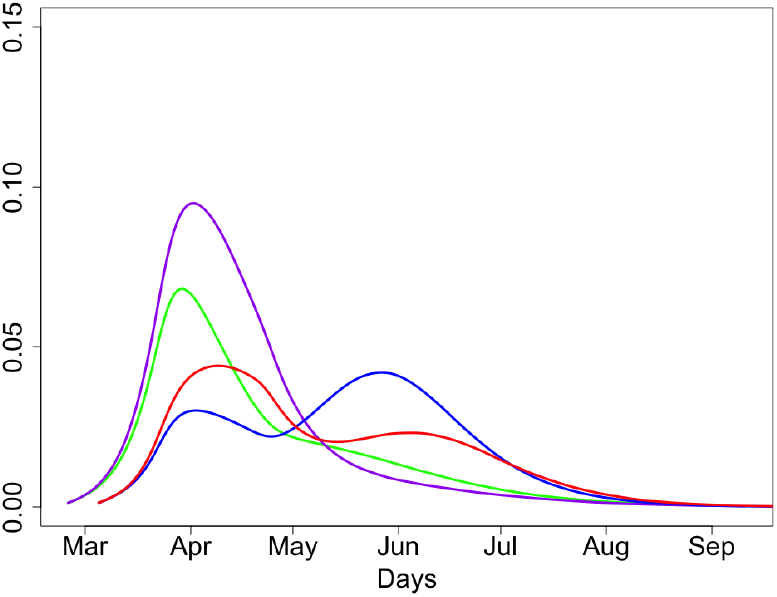
Proportions of people infected at time *t*; *x* = 0.9, 0.97.

**Figure 64:**
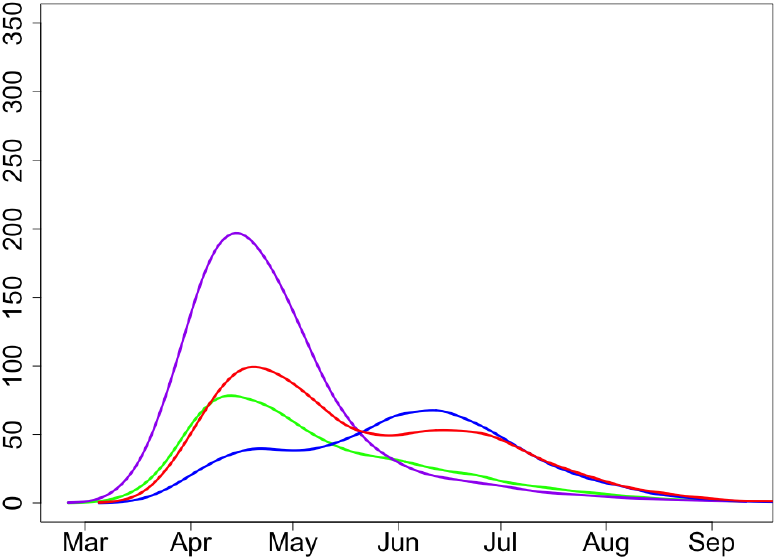
Expected deaths at time *t* at group *G* and the rest of population; *x* = 0.9, 0.97.

Figures 61–64 and the discussion above imply the following conclusion.

**Conclusion**. *A lock-down at an early stage of an epidemic is not a sensible decision in view of the economic consequences of the lock-down and the measures required for all the remaining (much longer) period of the epidemic. Moreover, in the case of an early lock-down, the second wave of the epidemic should be expected with a peak at around 2 months after the first one*.

In Figures 63 and 64, we use *x* = 0.97 and *x* = 0.9. Expected deaths toll for *x* = 0.97 is 13.8(5.7+8.3)K. This is naturally lower than 15(5.2+9.8)K for *x* = 0.9. This is related to the fact that we isolate people from group *G* much earlier.

A disadvantage of this scenario is the second wave of epidemic with a peak at around 2 months after the first one.

#### 8.5 Sensitivity to *R*_1_, the reproductive number during the lock-down

*R*_1_, the reproductive number during the lock-down, is an important factor of the model. We have assumed *R*_1_ = 1 which we believe to be a reasonable assumption overall. In Figures 65 and 66, we use *R*_1_ = 1.0 and 0.75. Decrease in *R*_1_ (that is, in the degree of isolation during the lock-down) leads to a significant decrease in the number of infections, severe cases and expected death numbers during the lock-down and about further 3 weeks but, since a larger proportion of the population is still susceptible, the epidemic is prolonged and the numbers for infected and dead at time *t* larger than *t*_2_ plus one month are visibly higher.

**Figure 65:**
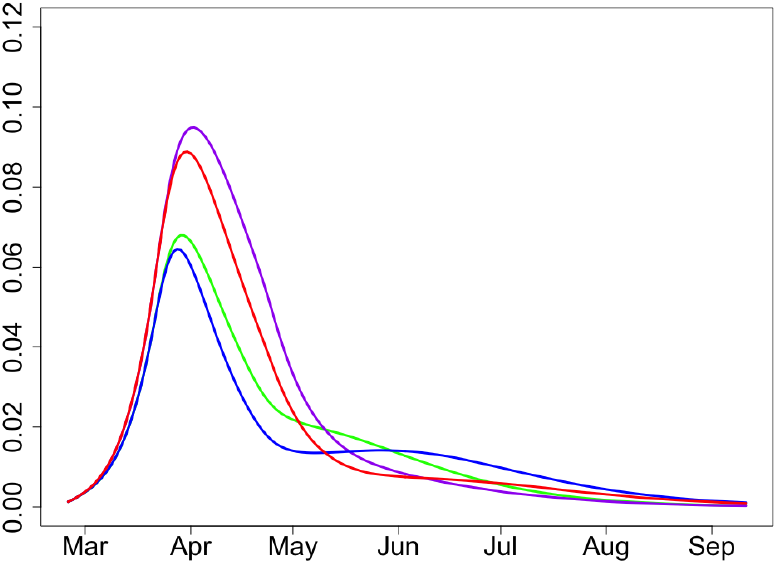
Proportions of people infected at time *t*; *R*_1_ = 1, 0.75.

**Figure 66:**
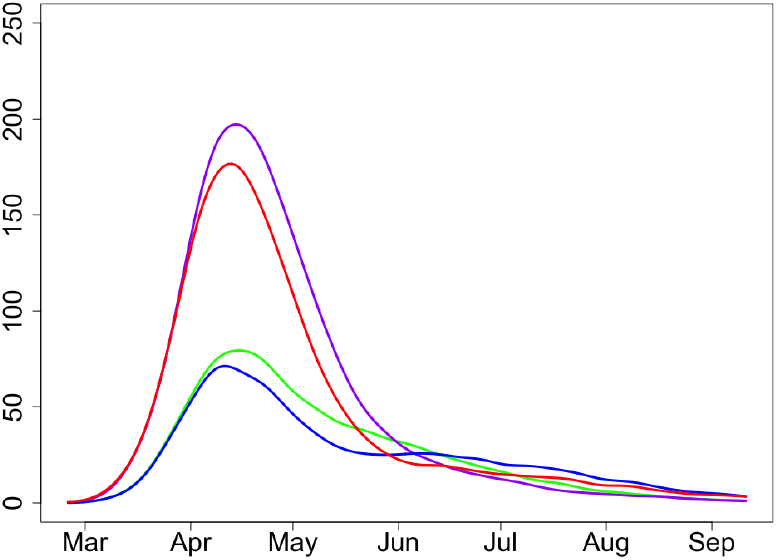
Expected deaths at time *t* at group *G* and the rest of population; *R*_1_ = 1, 0.75.

It could also be assumed that *R*_1_ may be changing in time. Indeed, *R*_1_ = *R*_1_(*t*) could be decreasing for the following reasoning. Initially, we suppose that every infected person will still infect all others in the same household, so that *R*_1_(*t*) could be larger than 1 for *t* close to March 23. Once the household has been infected, *R*_1_(*t*) drops even further to values which may be significantly lower than 1 and there will be very few new infections.

If we consider this situation realistic and hence assume *R*_1_(*t*) *>* 1 during the first week of the lock-down. As an example, consider the scenario with *R*_1_ = 1.25 during the first week of the lock-down and *R*_1_ = 0.75 thereafter; see Figures 67 and 68. Since 1.25 *>* 1, there are slightly more infected and dead in the first weeks after the lock-down but as 0.75 *<* 1, it gets compensated later. Overall, the main features of the epidemic in the new scenario are very similar to the ones we have chosen as the main ones.

**Figure 67:**
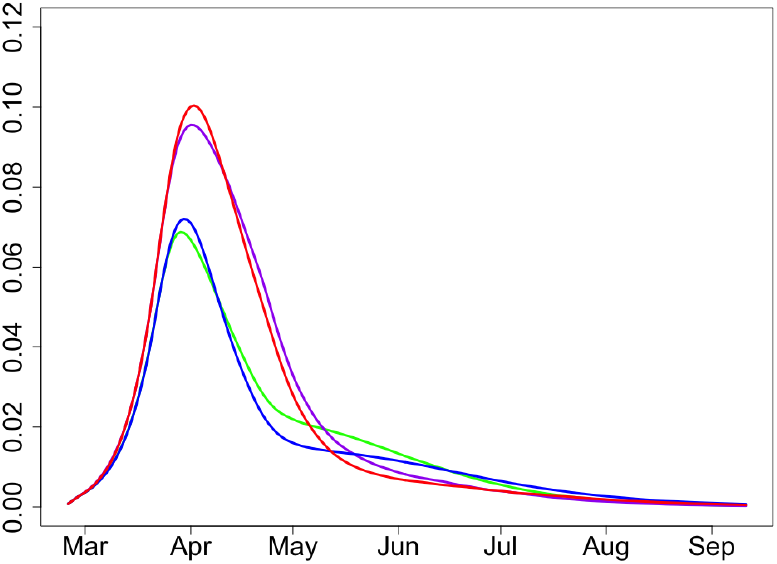
Proportions of people infected at time *t*; *R*_1_ = 1 versus *R*_1_ = 1.25 in the first week and 0.75 after.

**Figure 68:**
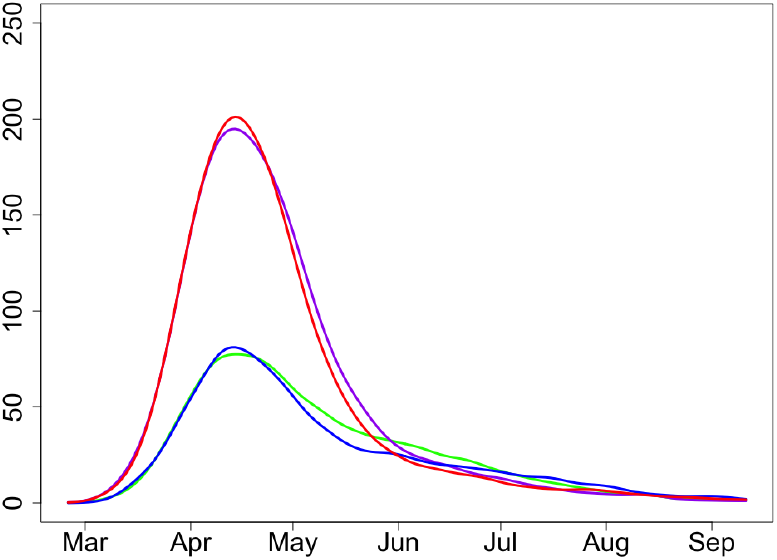
Expected deaths at time *t* at group *G* and the rest of population; *R*_1_ = 1 versus *R*_1_ = 1.25 in the first week and 0.75 after.

**Conclusion**. *Dynamics of the epidemic is rather sensitive to the value of R*_1_. *Decrease in R*_1_ *inevitably implies a visible decrease in the number of severe cases and expected death numbers in the first few weeks after the lock-down and small increase in these numbers at about 3 weeks after the lock-down is lifted. In the scenario with R*_1_ *decreasing monotonically in time, we see elevated number of infections and deaths in the first stage of the lock-down with visible decrease later on*.

#### 8.6 Sensitivity to *R*_2_, the reproductive number after lifting the lock-down

In Figures 69 and 70, we use *R*_2_ = 2.0 and 2.5. Increase in *R*_2_ would lead to a significant increase in the number of severe cases and expected death numbers.

**Figure 69:**
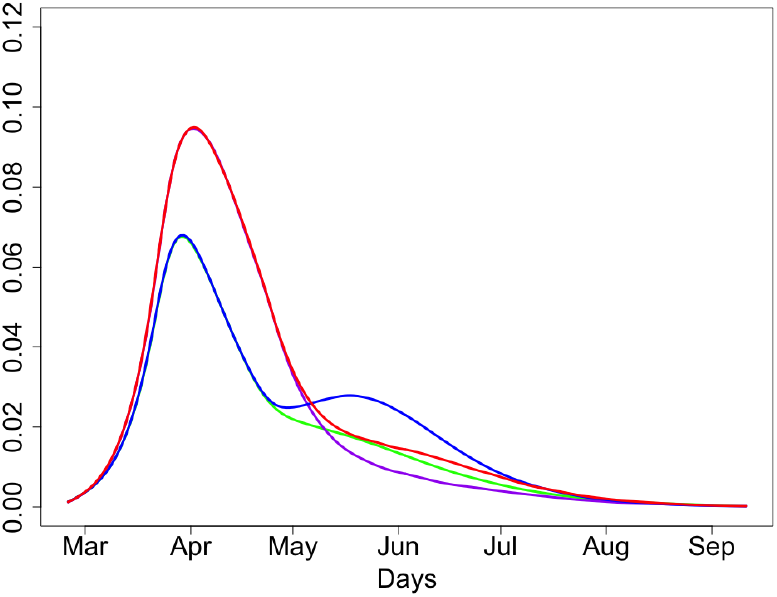
Proportions of people infected at time *t*; *R*_2_ = 2, 2.5.

**Figure 70:**
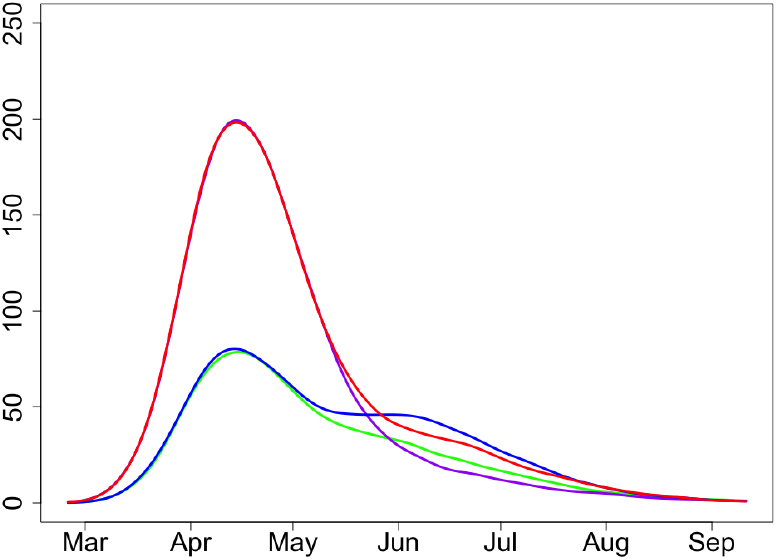
Expected deaths at time *t* at group *G* and the rest of population; *R*_2_ = 2, 2.5.

Expected deaths toll for *R*_2_ = 2.5 is 16.9(5.9+11)K. This is significantly higher than 15(5.2+9.8)K for *R*_2_ = 2. This implies that the public should carry on some level of isolation in the next 2-3 months.

**Conclusion**. *Increase in the reproductive number after lifting the lock-down would inevitably imply a significant increase in the number of severe cases and expected death numbers*.

#### 8.7 Sensitivity to *k*_*M*_ and *k*_*S*_, the shape parameter of the Erlang distributions for mild and severe cases

In Figures 71 and 72, we use *k*_*M*_ = 1, 3. The value *k*_*M*_ = 3 is default while the value *k*_*M*_ = 1 defines the exponential distribution for the period of infection in the case of mild disease and is equivalent to the corresponding assumption in SIR models. As the variance of the exponential distribution is larger than of the Erlang with *k*_*M*_ = 3 (given the same means), the epidemic with *k*_*M*_ = 1 runs longer and smoother.

**Figure 71:**
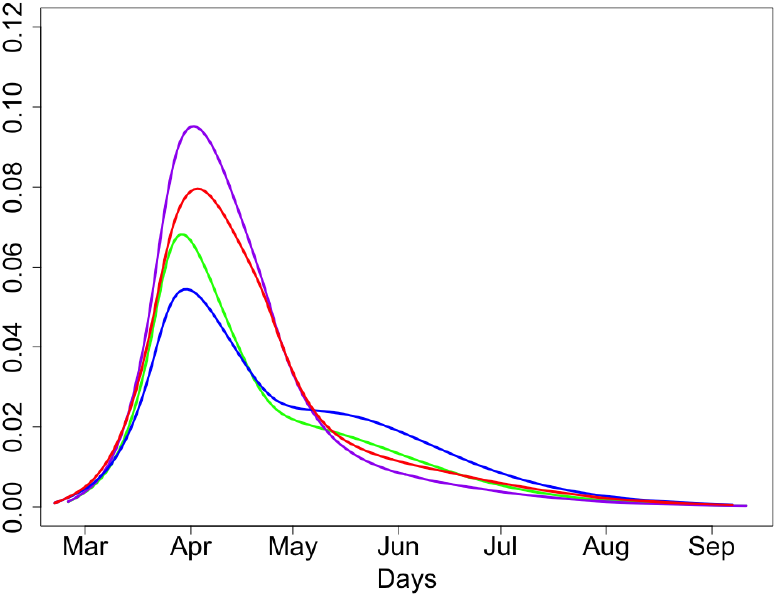
Proportions of people infected at time *t*; *k*_*M*_ = 1, 3.

**Figure 72:**
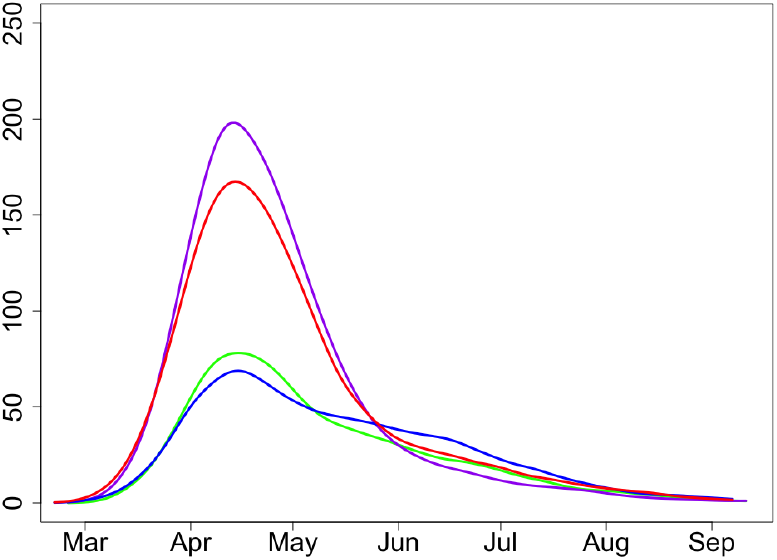
Expected deaths at time *t* at group *G* and the rest of population; *k*_*M*_ = 1, 3.

In Figures 73 and 74, we use *k*_*S*_ = 1, 3. Parameter *k*_*S*_ is less sensitive than *k*_*M*_ (as there are less severe cases than the mild ones) but it is also is.

**Figure 73:**
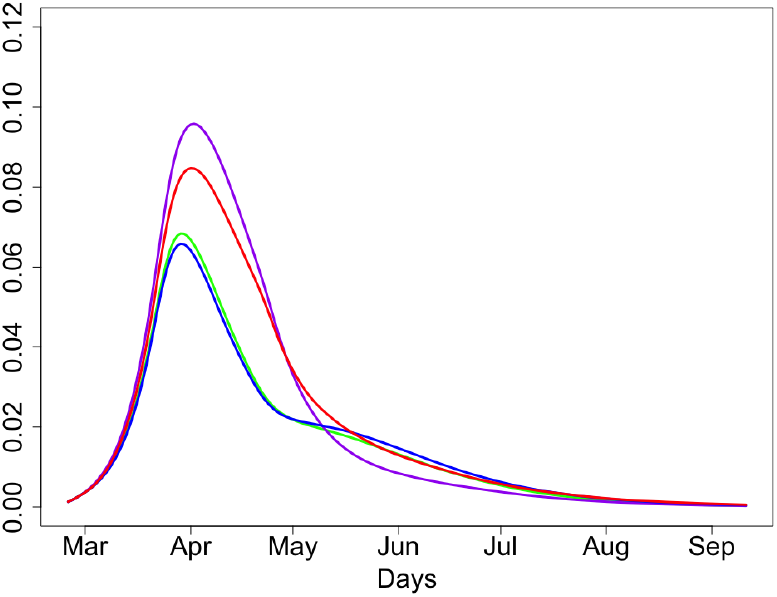
Proportions of people infected at time *t*; *k*_*S*_ = 1, 3.

**Figure 74:**
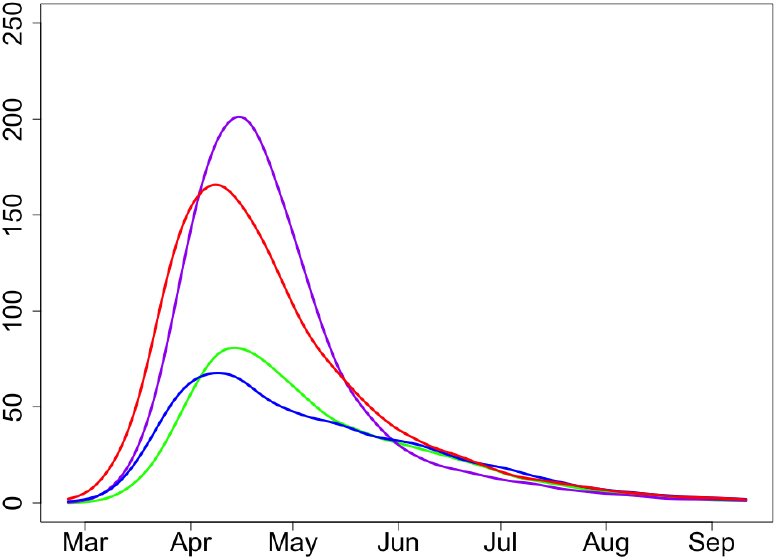
Expected deaths at time *t* at group *G* and the rest of population; *k*_*S*_ = 1, 3.

**Conclusion**. *The parameter k*_*M*_ *is rather important and more information is needed about the distribution time of infectious period. Parameter k*_*S*_ *is less sensitive than k*_*M*_ *but it is also is*.

#### 8.8 Sensitivity to *δ*, the probability of death in severe cases

In Figures 75 and 76, we use *δ* = 0.2 and 0.1. Decrease of *δ* increases the number of severe cases but does not change the expected death numbers.

**Figure 75:**
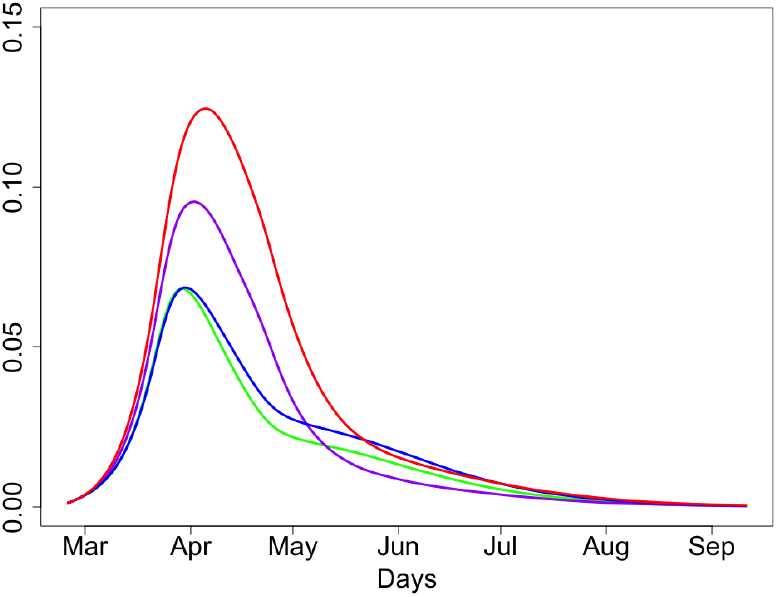
Proportions of people infected at time *t*; *δ* = 0.1, 0.2.

**Figure 76:**
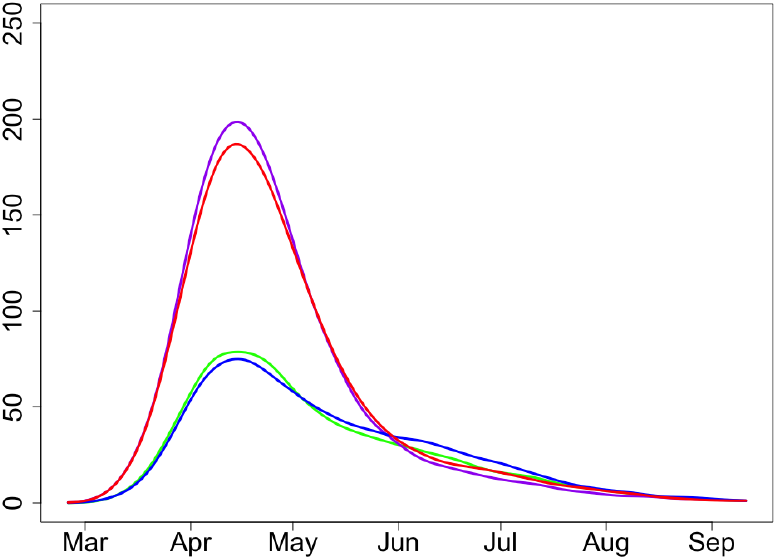
Expected deaths at time *t* at group *G* and the rest of population; *δ* = 0.1, 0.2.

### 9 Assessing benefits of the lock-down and the optimal time of lifting the lock-down

Using the example of the UK, we assess the benefit of the lock-down and discuss what is the optimal time for lifting this lock-down.

#### 9.1 A typical region

Consider a particular large city (like London or Sheffield), a small city (like Cardiff or Bath), a smaller town (like Bangor) or a particular county (like Cumberland). Call the chosen region **C** and all people of the chosen place ‘population’. Assume that this population is demographically similar to the entire UK population and that the population density and social behaviour are somewhat similar in this city. In this way, we are assuming that we can assign to this city a particular value of the reproductive number *R*_0_ *before* the lock-down.

For London, Sheffield and Manchester the reproductive number *R*_0_ could be close to 2.5 but for rural areas and small town *R*_0_ should be much smaller, even close to 1. Since the current lock-down is not very strict, the reproductive number during the lock-down (we denote this number *R*_1_) stays somewhere close to 1.

Let *x* denote the proportion of susceptible (non-immune, non-infected) people in **C** at the start of the lock-down, on March 23. This is a number between 0 and 1. The proportion of infected in **C** by March 23 is 1 − *x*.

#### 9.2 Assessing the benefits of the lock-down regionally

Consider the chosen region **C** with reproductive number *R*_0_ and the proportion of infected on March 23 1 − *x*. The numbers *x* and *R*_0_ very much determine the course of the epidemic in **C**, with and without the lock-down.

The are two reasons why 1 − *x* could be low (like 0.05 or lower):

(a) delayed start but rather large reproductive number *R*_0_ like 2.5,
(b) small reproductive number *R*_0_ like 1.5 or 2 (plus possibly late start).

The are also two reasons why 1 − *x* could be large (like 1 − *x* = 0.1 or larger):

(c) very early start but small reproductive number *R*_0_,
(d) rather large reproductive number *R*_0_ like 2.5.

The case (c) is unlikely (we would have seen this in the data) but we still will consider it.

First, let us discuss the benefits (in terms of hospital occupancy and saving lives) of the lock-down for a chosen city in these four cases.

In cases (b) and (c), with small values of *R*_0_, the lock-down is not very useful as the epidemic would have been manageable without any lock-down. Indeed, the lock-down in such places does not save any lives but just delays the course of the epidemic. Delaying a slowly running epidemic has no benefit as local hospitals would have been able to cope with demand anyway but the total number of infected and dead does not change (considering we are discussing the ‘herd immunity’ scenario). The only benefit of the lock-down is lowering *R*_0_ to *R*_2_ *< R*_0_ due to the awareness of the population. Note that other interventions might reduce the deaths further, but these are not modelled in this paper

The case (a) is considered in Section 8.4, Figures 61–64. As shown in these figures, the lock-down has definite benefit by disrupting the epidemic at the very early stage by ‘flattening the curve’ of the first wave (by this, saving lives). Unfortunately the early lock-down makes the arrival of the second wave about 2 months later very likely. Luckily, the second wave will not be anything like the projected first one (which is avoided) for the following three reasons:

i. there will be a smaller number of susceptible people in the city,
ii. *R*_2_, the reproductive number after lifting the lock-down, will be smaller than *R*_0_, and
iii. the group *G* of vulnerable people will hopefully stay well-isolated.

We believe the case (d) is the most important and London epidemic falls into the category covered by (d). In our main choice of parameters we have set 1 − *x* = 0.1, which may be an underestimation for inner London (that is, the real proportion of infected in London by March 23 might have been larger than 10%).

In case (d), the lock-down (even if terminated after 3 weeks) makes an important intervention to breaking the epidemic and saving many lives, compare Figures 47, 48 versus Figures 49, 50. In the case (d) no second wave is expected (the reasons are (i)-(iii), exactly as in the previous case) and the epidemic should gradually come to end in about 3 months time (by that time, about 2/3 of the population will be immune).

**Conclusion**

- *In regions with small R*_0_ *the lock-down is not beneficial*.
- *In cities with large R*_0_ *but late start the lock-down is beneficial but the second wave should be expected*.
- *In cities with large R*_0_ *and normal start the lock-down is beneficial and no second wave is expected*.

#### 9.3 The optimal time of lifting the lock-down, *R*_1_ = 1

Here we assume *R*_1_ = 1. Sensitivity of the epidemic towards different assumptions about *R*_1_ is discussed in Section 8.5.

We have already argued in the previous Section that in the regions with small *R*_0_ the lock-down is not beneficial. Hence, in the regions concerned, it is better to lift it as soon as possible.

Consider only the regions with large *R*_0_. Assume that during the lock-down we have reproductive number *R*_1_ = 1.

Current estimates show that in the case of death, the average delay between getting the COVID-19 virus and dying from it is about 21 days. This implies that after 21 days of the lock-down, the mortality should go down. The curve representing the proportion of newly infected people at time *t* should have started to decrease starting after about 12 days after the start of the lock-down (here 12=5+7, where 5 is the mean of the period of infection and 7 is approximately the mean of the infectious period). Therefore, under the assumption *R*_1_ = 1 all the curves (the expected number of deaths, expected hospital bed occupancy and expected number of infectious people) go down.

For any choice of time *t*_2_ of lifting the lock-down, which is April 13 or later (so that the lock-down holds for 21 days or longer), the rest of epidemic will continue to follow very similar paths, see Figures 55 and 56 and discussion in case when for 1 − *x* = 0.1 and Figures 59,60 when 1 − *x* is larger.

If 1 − *x* is smaller than 0.1 in the chosen region, then the curves representing proportion of infected and expected deaths are even less sensible to continue the lock-down beyond April 13 as the epidemic has almost stopped in such regions.

**Conclusion**. *In case R*_1_ = 1, *keeping the lock-down beyond April 13 has very little benefit in terms of the number of overall expected deaths and hospital bed occupancy in all cases (a)-(d) considered in Section 9.2*.

Note however that if we assume that *R*_1_ is monotonically decreasing during the lock-down as discussed in Section 8.5, then the optimal time of lifting the lock-down could be shifted one more week beyond April 13.

#### 9.4 Future development of the epidemic in the UK

If the proportion of susceptible in the UK is 80% or lower then the worst of the epidemic will be over very soon even if the lock-down terminates now. However, the proportion of susceptible in the UK can still be high (say, 95%). This is only possible if most of the UK regions/cities fall into the categories either (a) or (b) of Section 9.2. For regions falling into category (b), there are no benefits in keeping the lock-down (as there were very few reasons to start it). For regions falling into category (a), keeping the lock-down beyond April 13 would not alternate the future dynamics of the epidemic, and delay in lifting the lock-down would only postpone the inevitable. Unfortunately, in these regions we may expect second waves with peaks about 2 months later.

## Part III. One-stage exit scenarios starting at different dates from April 20, 2020

### Introduction

In this part we show results describing further development of the COVID-19 epidemic in the UK given the current data and assuming different dates for lifting the lock-down, which we have chosen to be April 20, April 27, May 4 and May 11. We use the model of the COVID-19 epidemic in the UK suggested and investigated in [1, 2].

We model differently the regions with high initial reproductive number chosen to be *R*_0_ = 2.5 and a lower one *R*_0_ = 2. The expected numbers of deaths are computed for a city or region with population 3 million people. The numbers for the whole of the UK can be obtained by appropriate averaging of the numbers given in the report.

For each scenario considered, we plot the expected proportion of infected at time *t* and the expected number of deaths at time *t*. We also give values of the following related quantities: *x* – the proportion of susceptible on March 23, *I*_0_ – the proportion of infected on April 16, and *I*_1_ – the proportion of infected from the start of the epidemic up to April 16.

The cities with large *R*_0_ = 2.5 are considered in Section 10. The main findings of Section 10 are:

- *The scenarios of Section 10.1.2 with large R*_0_ *and large x* = 0.97 *(and hence small I*_0_ *and I*_1_*) seem to be inadequate* as in these scenarios the peak in expected deaths should be arriving at latter dates, which seem to contradict to the current situation.
- In case *R*_0_ = 2.5, the range of most suitable values for *x* is around 0.9. *For such values of x, the epidemic in the badly hit regions of the UK is in the fast monotonic decline irrespectively of the date of the lock-down lift*.
- A rather sensitive case, considered in Section 10.2, with large *R*_0_ and rather large *x* = 0.95 seems to be less adequate than *x* = 0.9. In this case, a mild second wave may be expected in June for all dates of lifting the lock-down.

The regions with smaller *R*_0_ = 2 are considered in Section 11. The main findings of Section 11 are:

- In most natural scenarios of Section 11 with *R*_0_ = 2, the second wave should be expected for all dates of lifting the lock-down, but it is going to be mild in all scenarios.
- *For R*_0_ = 2, *the difference between the expected deaths is very small for the lifting lock-down dates* April 27, May 4 *and* May 11, *for all cases considered in Section 11*.
- Among all scenarios with *R*_0_ = 2, the scenarios of Section 11.2 look most reasonable. In these scenarios, despite rather high *x* and low *I*_0_, *I*_1_, in the very reasonable case *R*_2_ = 1.5, irrespectively of the date of lifting the lock-down, there will be no second wave in expected deaths, see Figure 100.

For the overall situation, we can observe the following:

- The expected numbers of deaths in the scenarios with large *R*_0_ of Section 10 are significantly higher than for the cases with smaller *R*_0_ = 2 considered in Section 11.
- Depending on how many regions in the UK are still almost non-affected by the epidemic and illustrated in Section 11.1, there could be a second wave of the epidemic in June (or July if the lock-down is kept until mid-May) but, *irrespectively of the date of lifting the lock-down, the second wave is going to be much milder than the first wave which is about to finish within two weeks*.

### Notation, model parameters and description of plots

- *t* - time (in days)
- *t*_1_ - the lock-down time, *t*_1_ = March 23
- *t*_2_ - time when the lock-down finishes, *t*_2_ = April 20, *t*_2_ = April 27, *t*_2_ = May 4 or *t*_2_ = May 11
- *I*(*t*) - number of infectious at time *t*
- *S*(*t*) - number of susceptible at time *t*;
- *R*_0_ - reproductive number before lock-down
- *R*_1_ - reproductive number during the lock-down (*t*_1_ ≤ *t < t*_2_)
- *R*_2_ - reproductive number after the lock-down finishes (*t* ≥ *t*_2_)
- *x* - the proportion of susceptible at the start of the lock-down
- *I*_0_ - the proportion of infected on April 16
- *I*_1_ - the proportion of infected from the start of the epidemic up to April 16

In the model, *x* is a parameter but *I*_0_ and *I*_1_ are computed; their values are uniquely determined from *x* and other parameters of the model.

As in [2], we consider a city of size 3 million people but we now use the recently updated number for the UK expected mortality rate *r* = 0.66%. We keep all other parameters of the model unchanged. In particular, we assume that after lifting the lock-down, the group of vulnerable people and people of 70+ is going to be rather well isolated. For the value of *R*_1_, we assume the following model. Initially, we suppose that every infected person will still infect all others in the same household, so that *R*_1_ could be larger than 1 for *t* close to March 23 and we assumed *R*_1_ = 1.1 during the week March 23 - March 29. Once the household has been infected, *R*_1_ drops to a value significantly lower than 0.9 (we set *R*_1_ = 0.9 for *t* ≥ March 30) as there will be very few new infections.

In all plots colours relate to the date of lifting the lock-down:

- Purple: *t*_2_ = April 20
- Green: *t*_2_ = April 27
- Red: *t*_2_ = May 4
- Black: *t*_2_ = May 11

In all figures with odd numbers, we use dashed lines and plot *I*(*t*)*/N*, the proportion of infected at time *t*. In all figures with even numbers, we use solid lines and plot expected deaths numbers at time *t*.

### 10 Regions with high initial reproductive number *R*_0_ = 2.5

#### 10.1 *x* = 0.98, *I*_0_ = 0.018, *I*_1_ = 0.138

##### 10.1.1 *R*_2_ = 2

**Figure 77:**
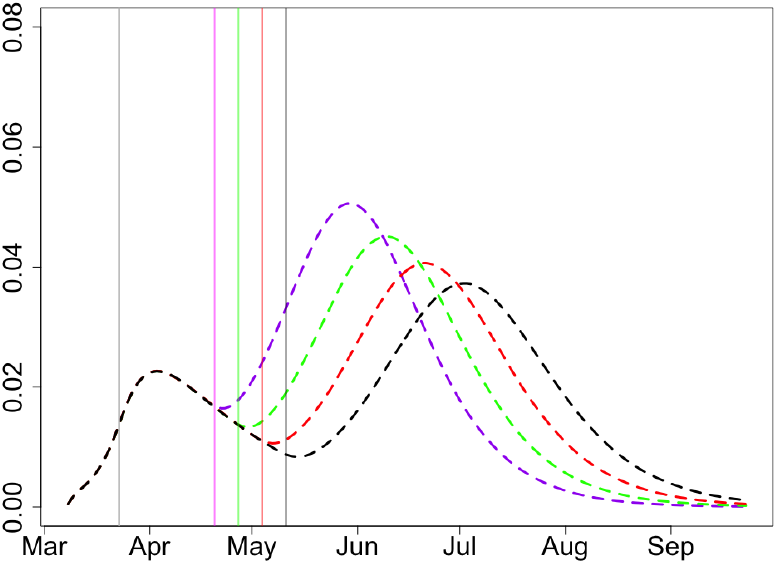
Proportion of infectious, *x* = 0.98, *R*_2_ = 2.

**Figure 78:**
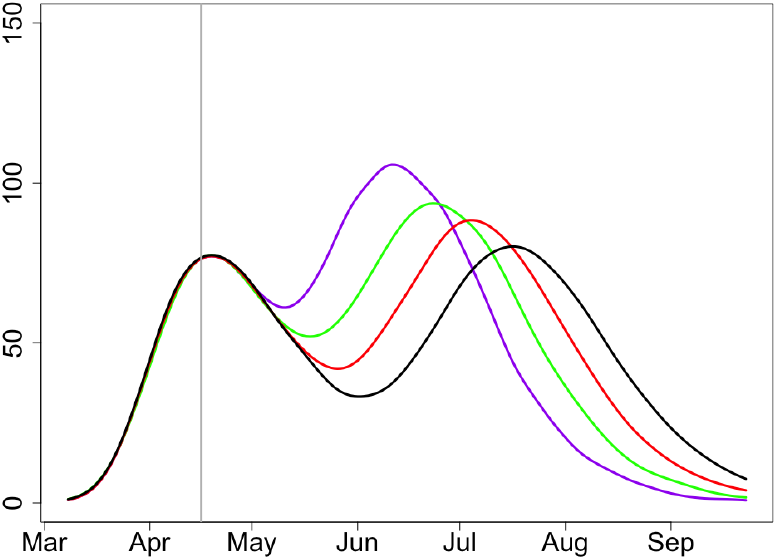
Expected deaths, *x* = 0.98, *R*_2_ = 2.

##### 10.1.2 *R*_2_ = 1.75

This scenario (with large *R*_0_ and large *x*) seems to be inadequate; in case of large *x* and hence small *I*_0_ and *I*_1_, the peak in expected deaths is shifted too far to the later dates.

**Figure 79:**
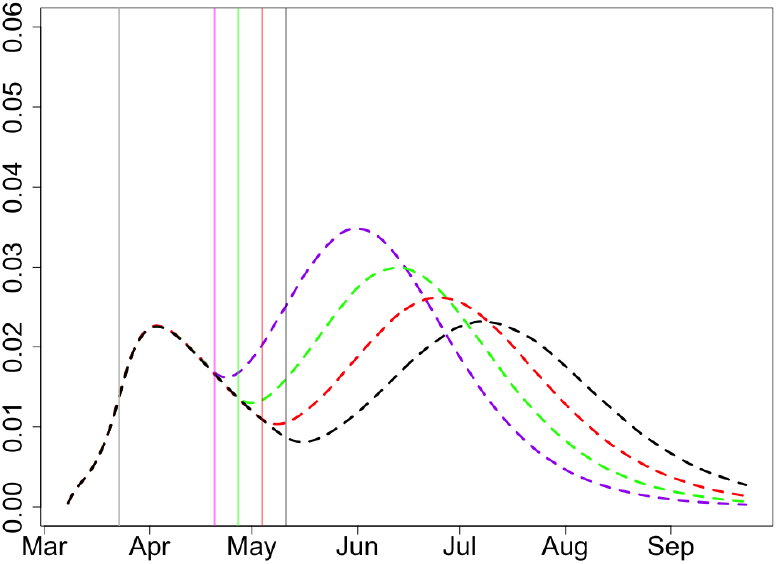
Proportion of infectious, *x* = 0.98, *R*_2_ = 1.75.

**Figure 80:**
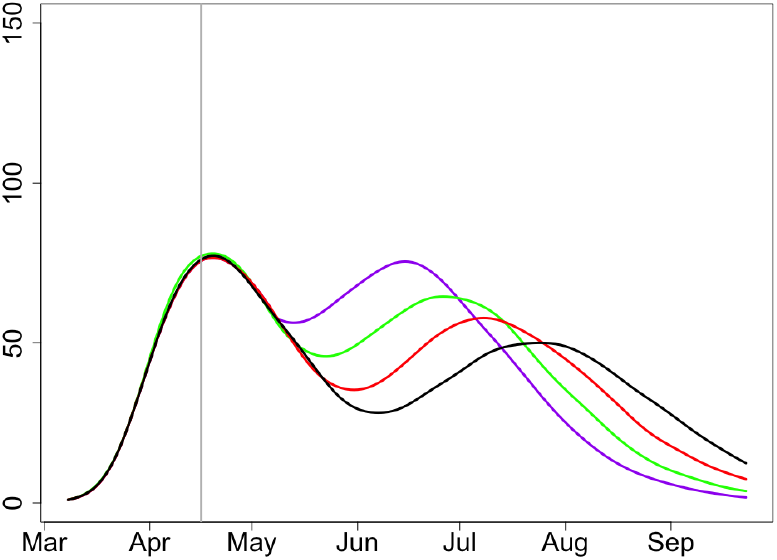
Expected deaths, *x* = 0.98, *R*_2_ = 1.75.

#### 10.2 *x* = 0.95, *I*_0_ = 0.0288, *I*_1_ = 0.255

##### 10.2.1 *R*_2_ = 2

**Figure 81:**
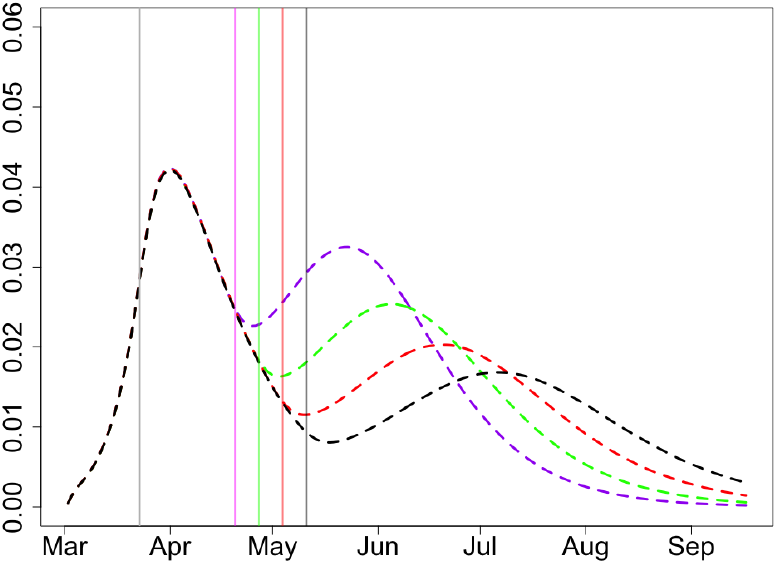
Proportion of infectious, *x* = 0.95, *R*_2_ = 2.

**Figure 82:**
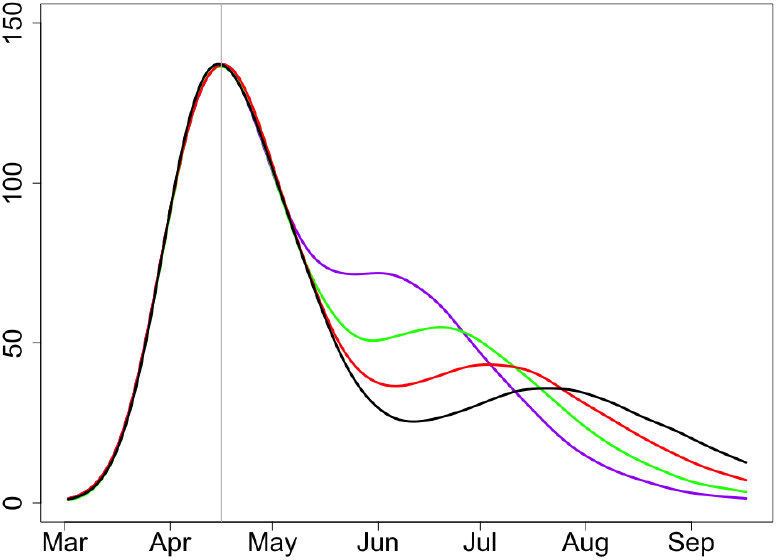
Expected deaths, *x* = 0.95, *R*_2_ = 2.

##### 10.2.2 *R*_2_ = 1.75

In view of the location of the peaks in the curve of the expected deaths, in the case *R*_0_ = 2.5, the scenarios with *x* = 0.95 seem to be more likely than with *x* = 0.98 but less likely than with *x* = 0.9. For *x* = 0.95, a visible second wave in the number of infections is only expected for reasonably high reproductive number *R*_2_ = 2 (and any date of lifting the lock-down). The second wave in the expected deaths is not as high (this is related to the assumption of a good isolation of vulnerable people).

**Figure 83:**
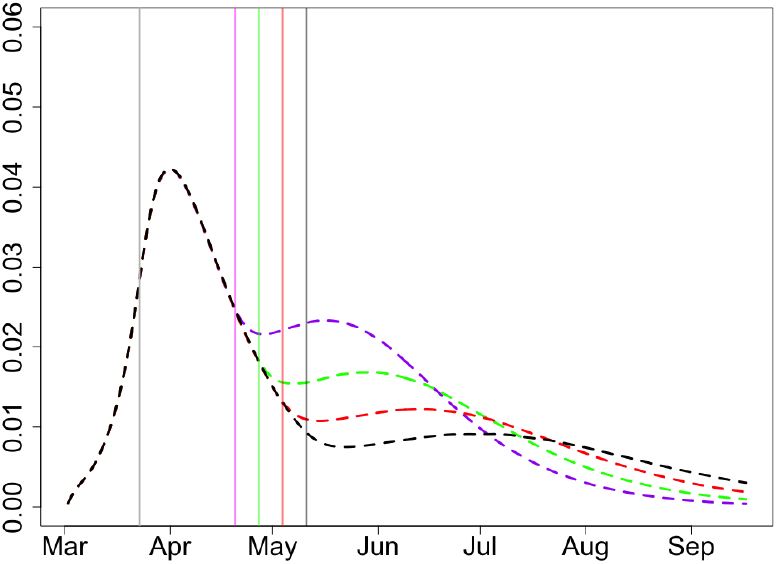
Proportion of infectious, *x* = 0.95, *R*_2_ = 1.75.

**Figure 84:**
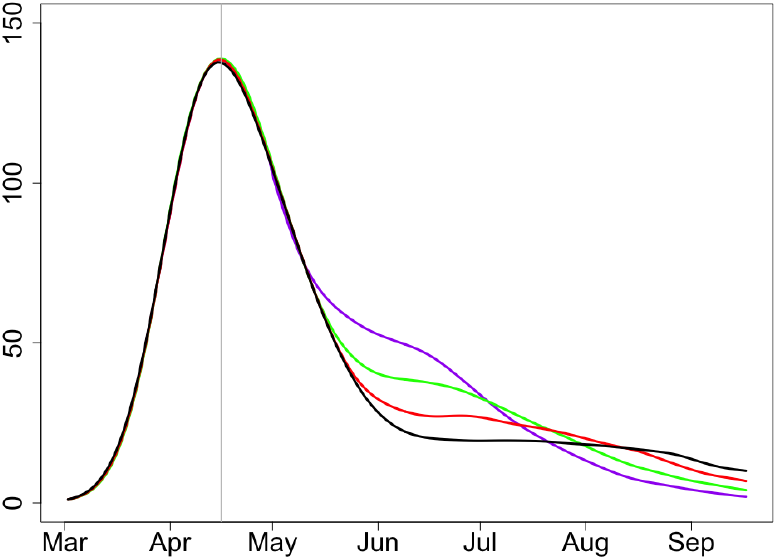
Expected deaths, *x* = 0.95, *R*_2_ = 1.75.

#### 10.3 *x* = 0.9, *I*_0_ = 0.035, *I*_1_ = 0.413

##### 10.3.1 *R*_2_ = 2

**Figure 85:**
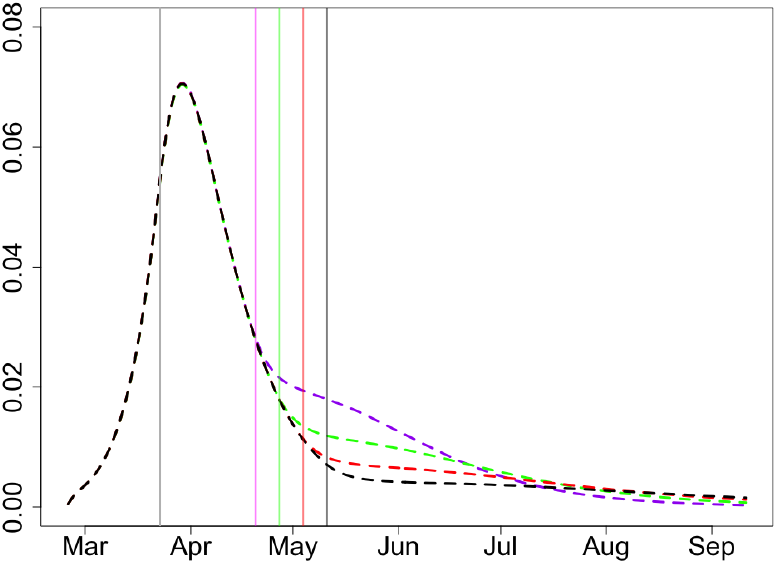
Proportion of infectious, *x* = 0.9, *R*_2_ = 2.

**Figure 86:**
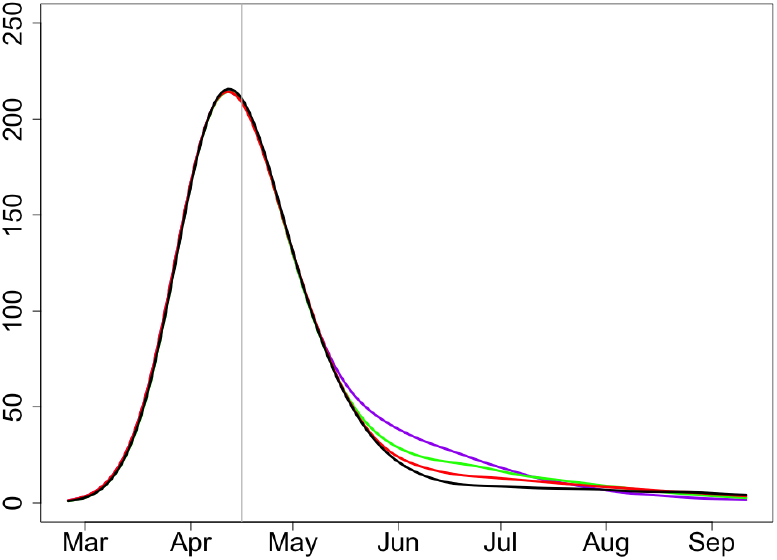
Expected deaths, *x* = 0.9, *R*_2_ = 2.

##### 10.3.2 *R*_2_ = 1.75

Among all considered scenarios with large reproductive number *R*_0_, the value *x ∼* 0.9 gives the best results matching the current data. In particular, the peak in expected deaths falls at April 10 plus/minus 1 day.

**Figure 87:**
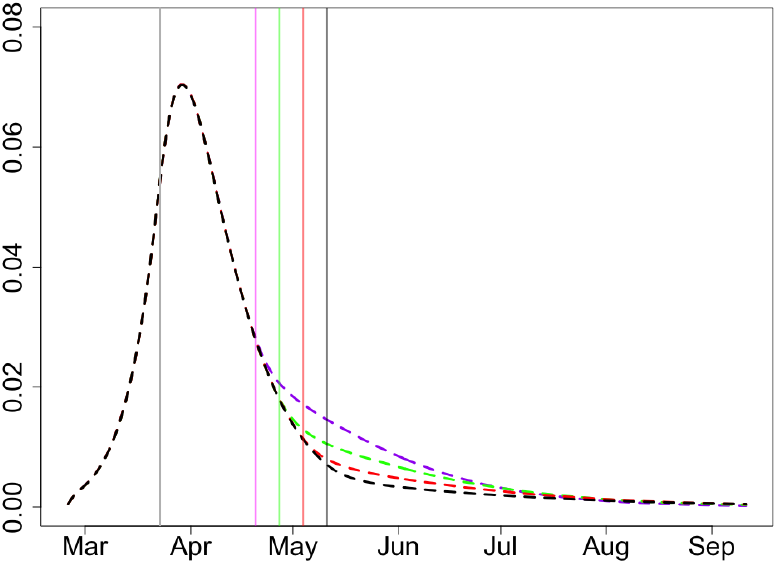
Proportion of infectious, *x* = 0.9, *R*_2_ = 1.75.

**Figure 88:**
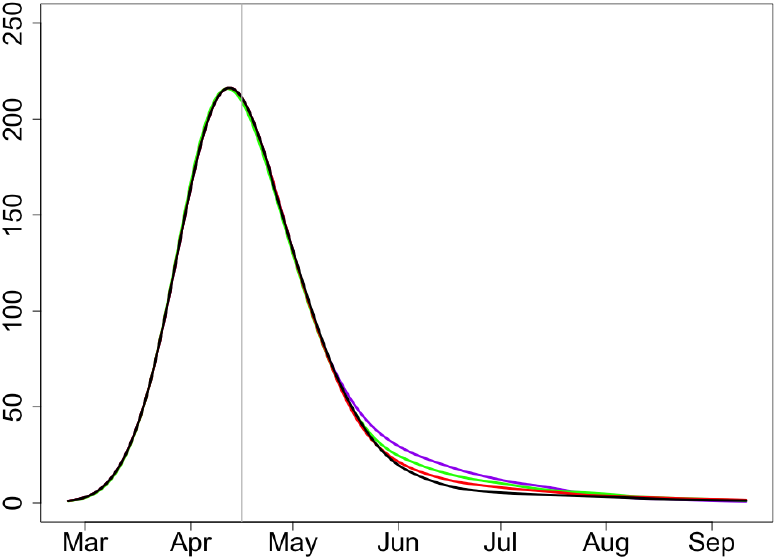
Expected deaths, *x* = 0.9, *R*_2_ = 1.75.

#### 10.4 *x* = 0.85, *I*_0_ = 0.035, *I*_1_ = 0.495

##### 10.4.1 *R*_2_ = 2

**Figure 89:**
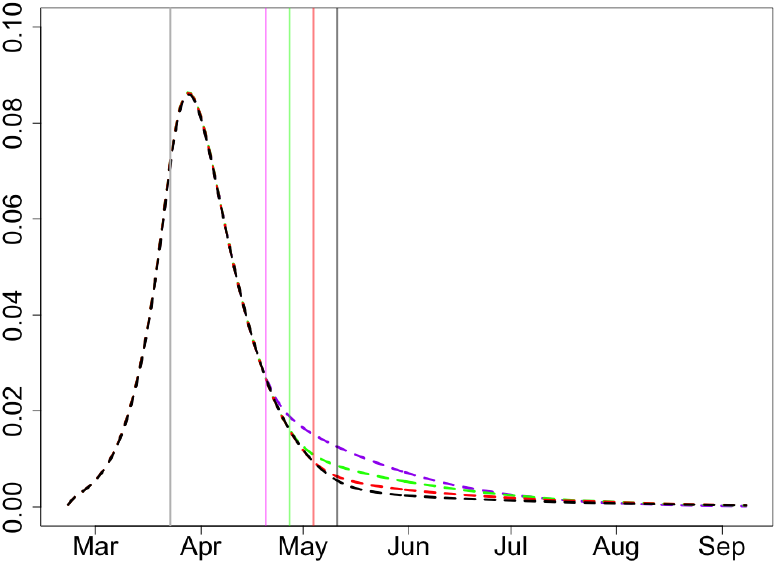
Proportion of infectious, *x* = 0.85, *R*_2_ = 2.

**Figure 90:**
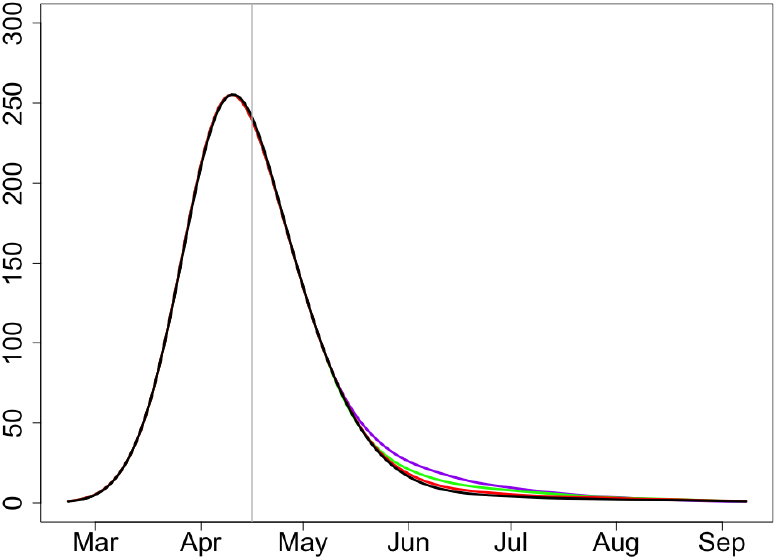
Expected deaths, *x* = 0.85, *R*_2_ = 2.

##### 10.4.2 *R*_2_ = 1.75

For *R*_0_ = 2.5, the value *x ∼* 0.85 seems to be too low as the peak in expected deaths would have arrived too early, at around April 5.

**Figure 91:**
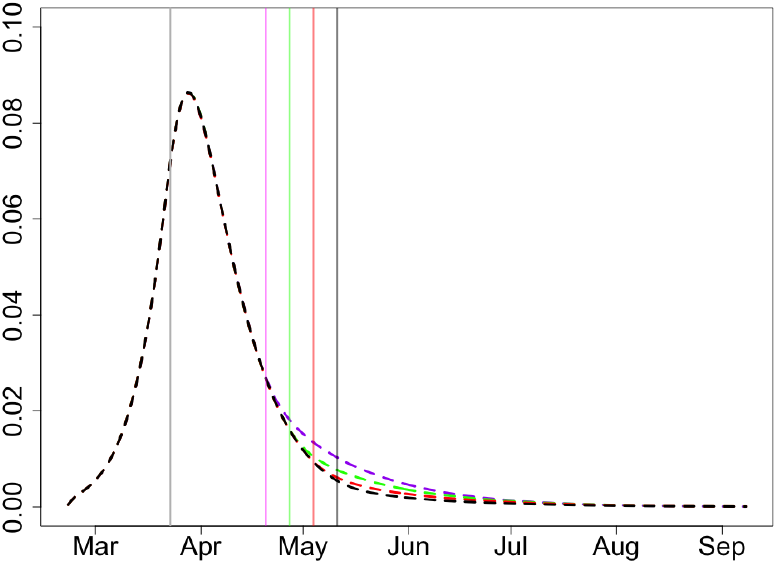
Proportion of infectious, *x* = 0.85, *R*_2_ = 1.75.

**Figure 92:**
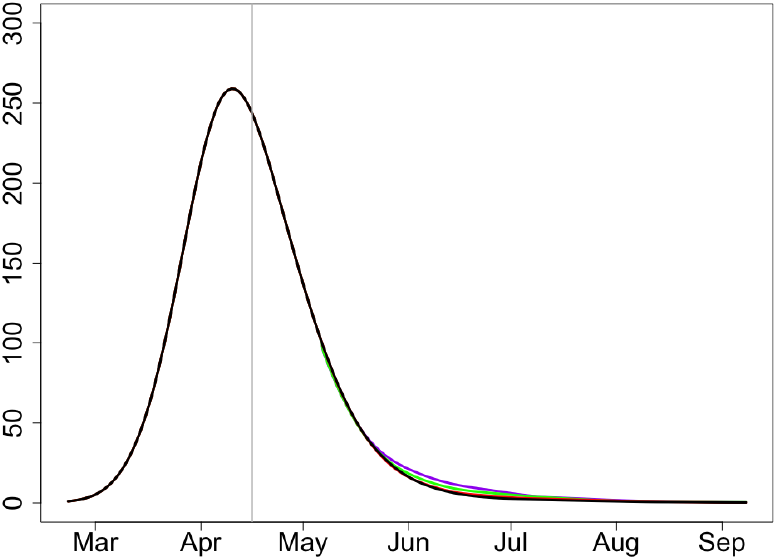
Expected deaths, *x* = 0.85, *R*_2_ = 1.75.

### 11 Regions with low initial reproductive number *R*_0_ = 2

#### 11.1 *x* = 0.98, *I*_0_ = 0.0132, *I*_1_ = 0.102

##### 11.1.1 *R*_2_ = 1.75

**Figure 93:**
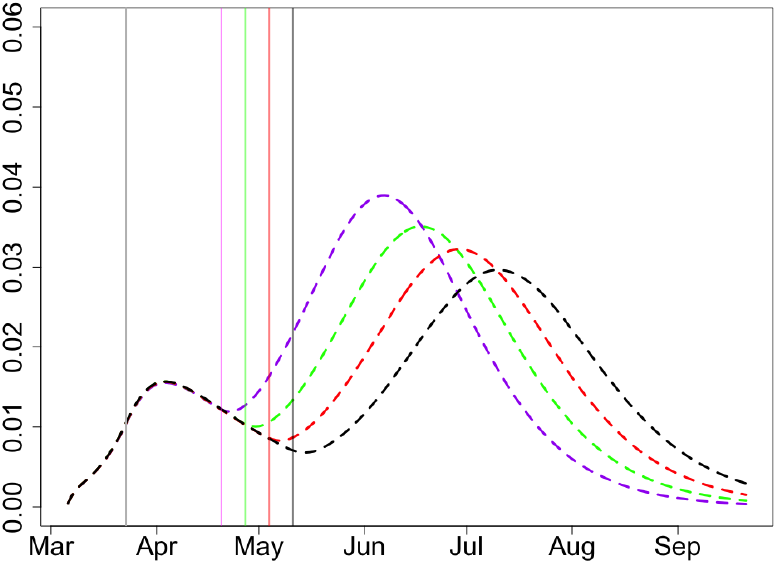
Proportion of infectious, *x* = 0.98, *R*_2_ = 1.75.

**Figure 94:**
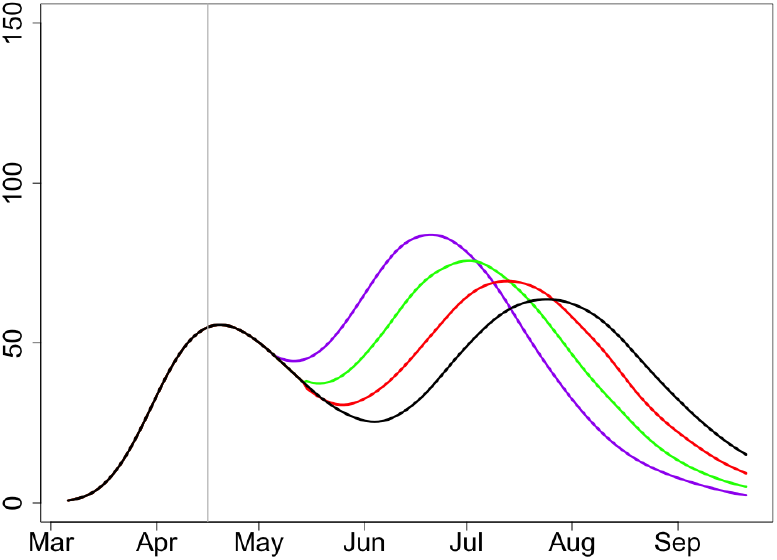
Expected deaths, *x* = 0.98, *R*_2_ = 1.75.

##### 11.1.2 *R*_2_ = 1.5

The scenario with *R*_0_ = 2 and very large *x* seems reasonable. The second wave should be expected in all cases but please note the scales for the proportions of infected and expected deaths; the numbers are much smaller than in the case *R*_0_ = 2.5.

**Figure 95:**
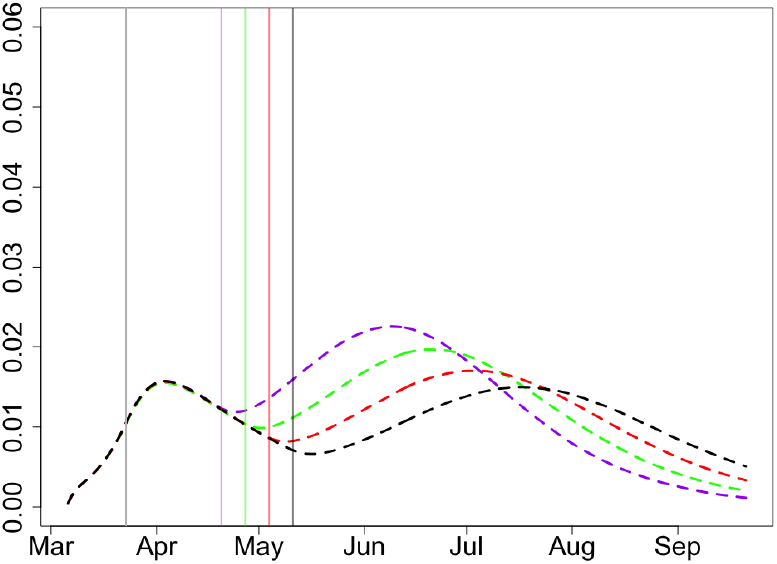
Proportion of infectious, *x* = 0.98, *R*_2_ = 1.5.

**Figure 96:**
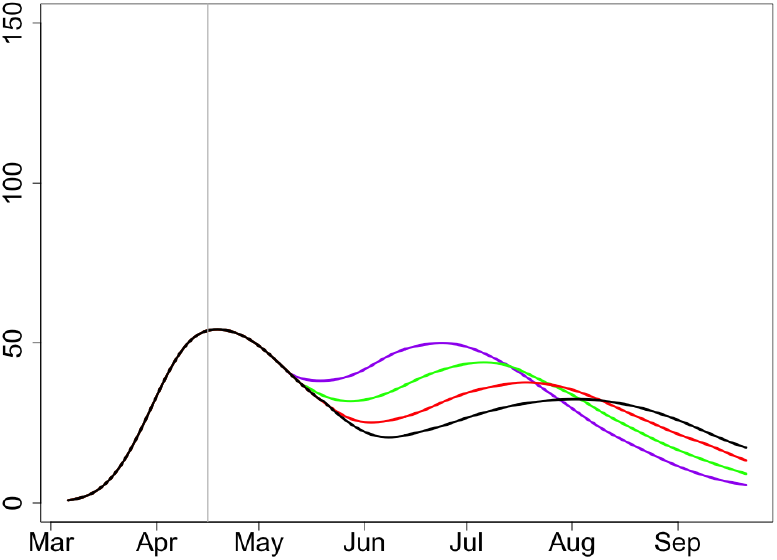
Expected deaths, *x* = 0.98, *R*_2_ = 1.5.

#### 11.2 *x* = 0.97, *I*_0_ = 0.016, *I*_1_ = 0.132

##### 11.2.1 *R*_2_ = 1.75

**Figure 97:**
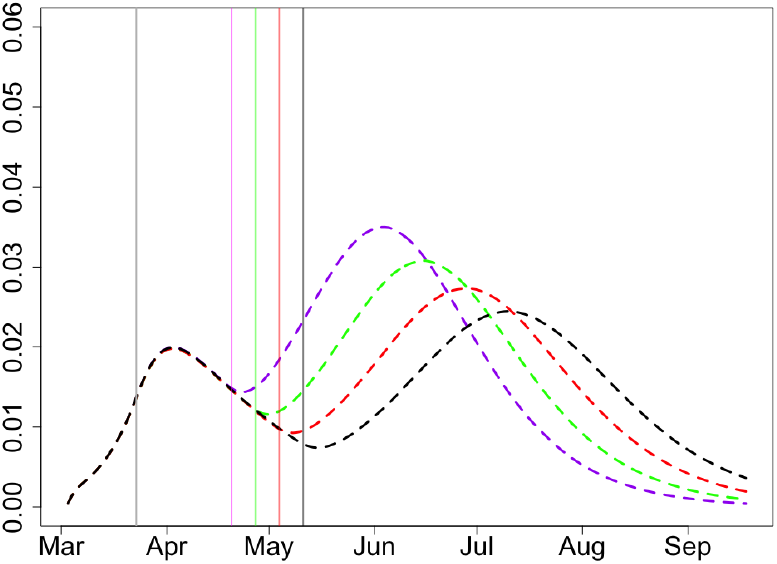
Proportion of infectious, *x* = 0.97, *R*_2_ = 1.75.

**Figure 98:**
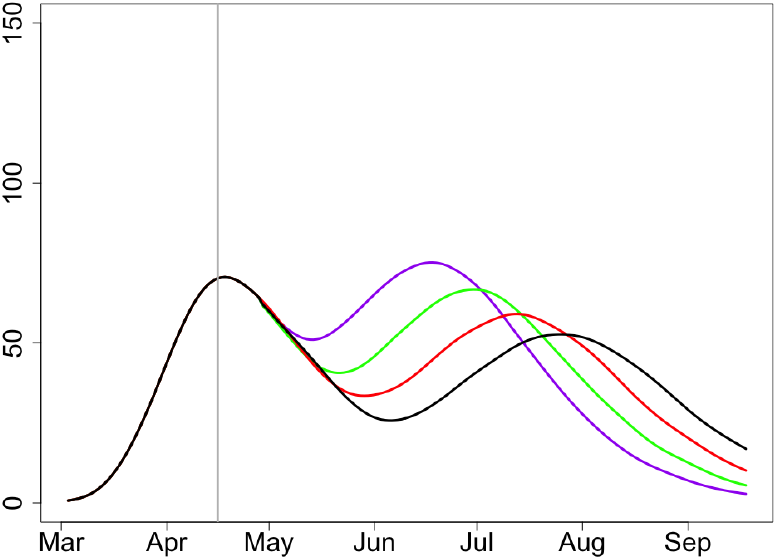
Expected deaths, *x* = 0.97, *R*_2_ = 1.75.

##### 11.2.2 *R*_2_ = 1.5

Among all scenarios with small *R*_0_, this scenario looks most reasonable. The second wave should be expected in all cases but the numbers are much smaller than in the cases with *R*_0_ = 2.5 and also smaller than with *R*_0_ = 2 and *x* = 0.98.

**Figure 99:**
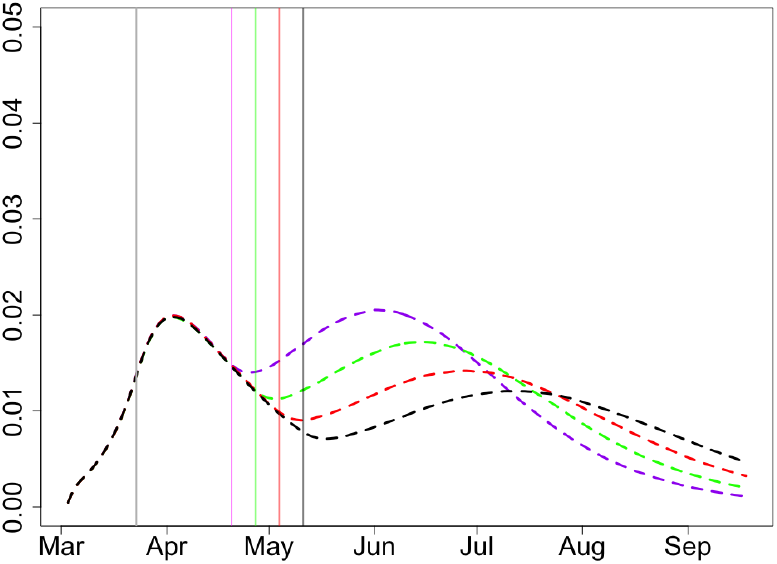
Proportion of infectious, *x* = 0.97, *R*_2_ = 1.5.

**Figure 100:**
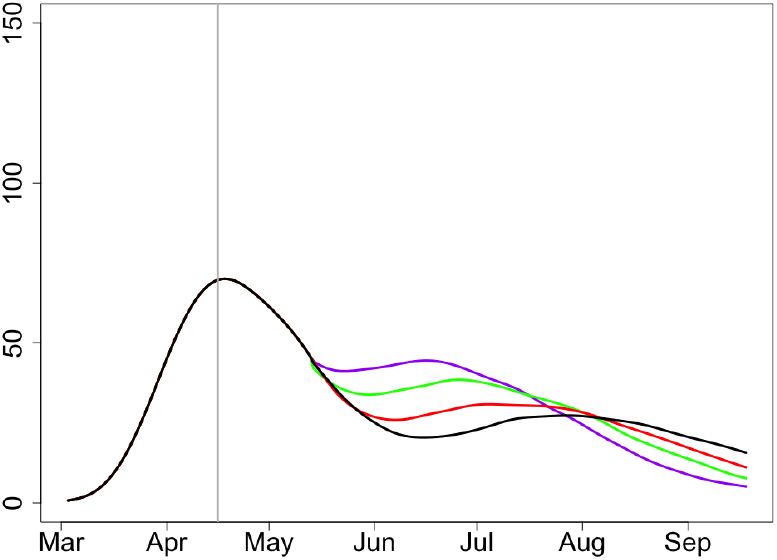
Expected deaths, *x* = 0.97, *R*_2_ = 1.5.

#### 11.3 *x* = 0.95, *I*_0_ = 0.022, *I*_1_ = 0.206

##### 11.3.1 *R*_2_ = 1.75

**Figure 101:**
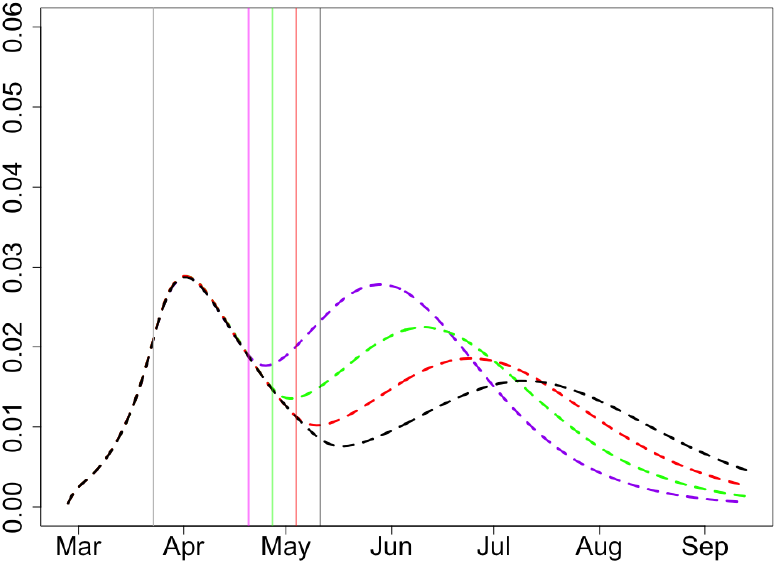
Proportion of infectious, *x* = 0.95, *R*_2_ = 1.75.

**Figure 102:**
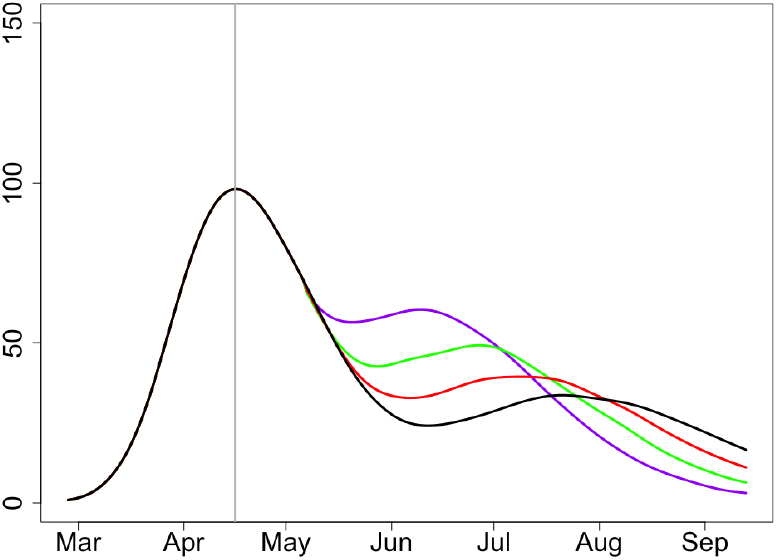
Expected deaths, *x* = 0.95, *R*_2_ = 1.75.

##### 11.3.2 *R*_2_ = 1.5

For *R*_0_ = 2, the value *x* = 0.95 is low enough to guarantee that there will be no visible second wave.

**Figure 103:**
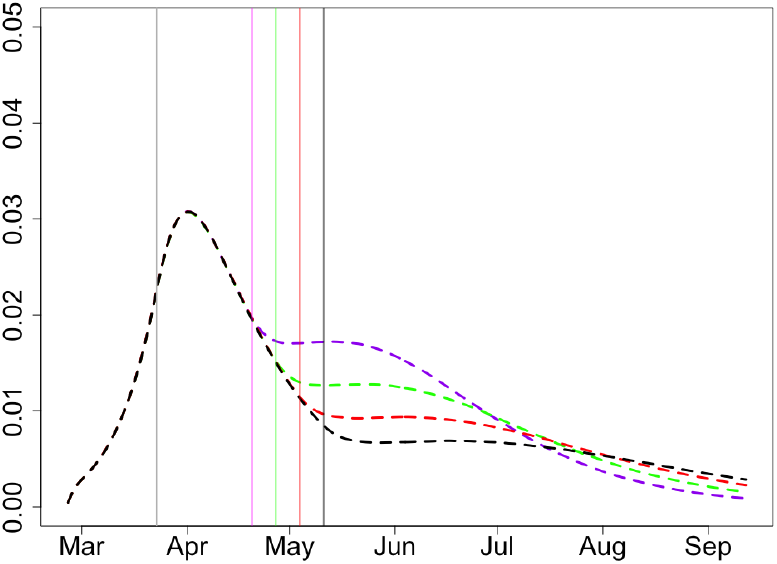
Proportion of infectious, *x* = 0.95, *R*_2_ = 1.5.

**Figure 104:**
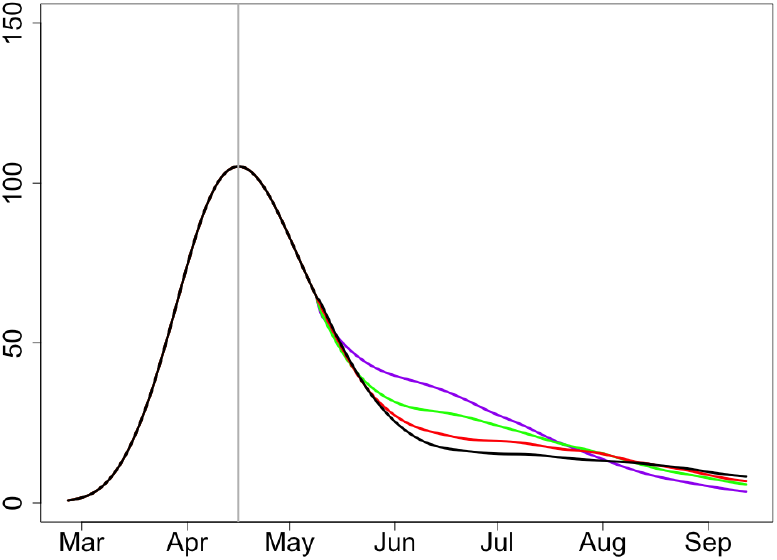
Expected deaths, *x* = 0.95, *R*_2_ = 1.5.

#### 11.4 *x* = 0.9, *I*_0_ = 0.028, *I*_1_ = 0.321

##### 11.4.1 *R*_2_ = 1.75

**Figure 105:**
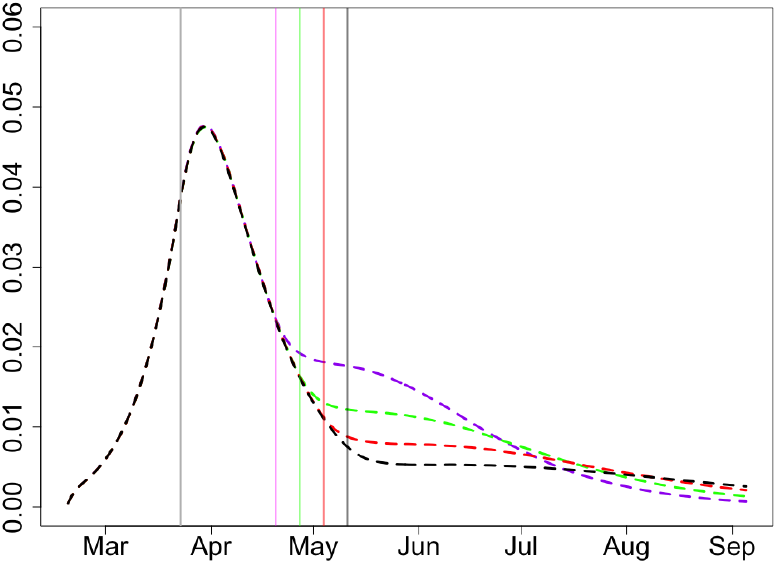
Proportion of infectious, *x* = 0.9, *R*_2_ = 1.75.

**Figure 106:**
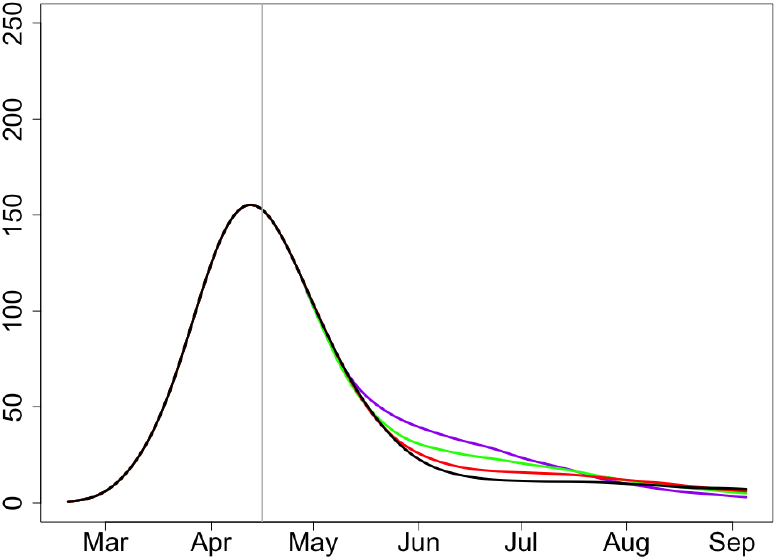
Expected deaths, *x* = 0.9, *R*_2_ = 1.75.

**Figure 107:**
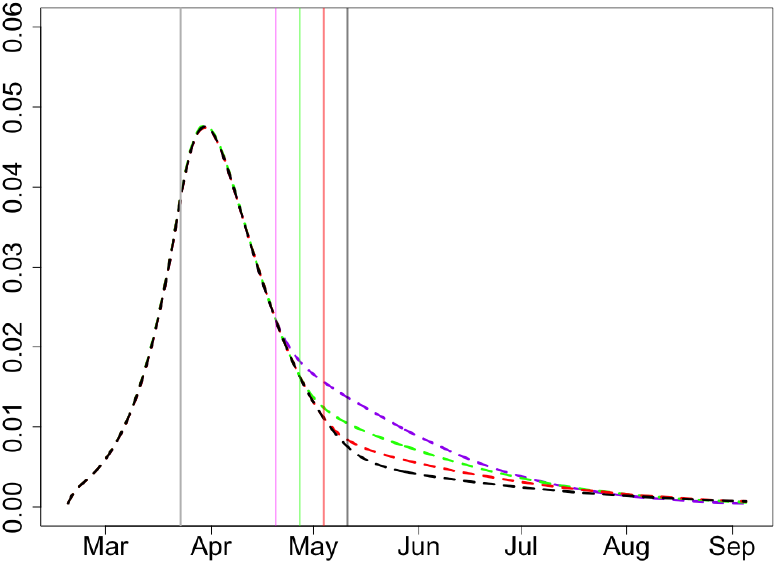
Proportion of infectious, *x* = 0.9, *R*_2_ = 1.5.

**Figure 108:**
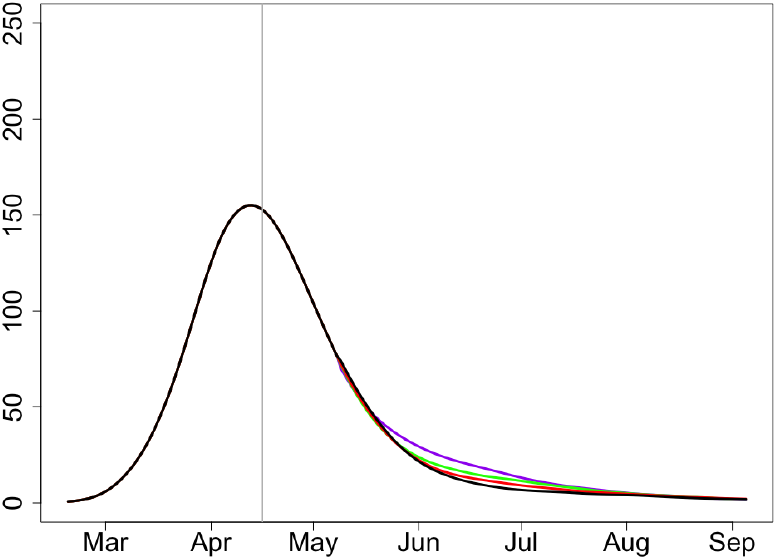
Expected deaths, *x* = 0.9, *R*_2_ = 1.5.

##### 11.4.2 *R*_2_ = 1.5

For regions with low *R*_0_, the value *x* = 0.9 guarantees that the epidemic will monotonically pass and the expected deaths will be almost unnoticeable from June.

## Data Availability

No data used. Only simulation based.

## Acknowledgement

We are grateful to Martin Hairer (ICL), Jürgen Branke (Univ. Warwick) and Jonathan Ball (Univ. Nottingham) for their valuable comments on parts of this paper and useful discussions.

## Notes

### Competing Interest Statement

The authors have declared no competing interest.

### Funding Statement

No external funding was received

